# Isoform-level transcriptome-wide association uncovers extensive novel genetic risk mechanisms for neuropsychiatric disorders in the human brain

**DOI:** 10.1101/2022.08.23.22279134

**Authors:** Arjun Bhattacharya, Daniel D. Vo, Connor Jops, Minsoo Kim, Cindy Wen, Jonatan L. Hervoso, Bogdan Pasaniuc, Michael J. Gandal

## Abstract

Integrative methods, like colocalization and transcriptome-wide association studies (TWAS), identify transcriptomic mechanisms at only a fraction of trait-associated genetic loci from genome-wide association studies (GWAS). Here, we show that a reliance on reference functional genomics panels of only total gene expression greatly contributes to this reduced discovery. This is particularly relevant for neuropsychiatric traits, as the brain expresses extensive, complex, and unique alternative splicing patterns giving rise to multiple genetically-regulated transcript-isoforms per gene. Integrating highly correlated transcript-isoform expression with GWAS requires methodological innovations.

We introduce isoTWAS, a multivariate framework to integrate genetics, isoform-level expression, and phenotypic associations in a step-wise testing framework, and evaluate it using data from the Genotype-Tissue Expression (GTEx) Project, PsychENCODE Consortium, and other sources. isoTWAS shows three main advantages. First, joint, multivariate modeling of isoform expression from *cis*-window SNPs improves prediction by ∼1.8-2.4 fold, compared to univariate modeling. Second, compared to gene-level TWAS, these improvements in prediction lead to ∼1.9-2.5-fold increase in the number of testable genes and a median of 25-70% increase in cross-validated prediction of total gene expression, with the added ability to jointly capture expression and splicing mechanisms. In external validation, isoform-centric models predicted gene expression at percent variance explained >1% for 50% more genes than gene-centric models. Third, across 15 neuropsychiatric traits, isoTWAS increased discovery of trait associations within GWAS loci over TWAS, capturing ∼60% more unique loci and 95% of loci detected by TWAS. Results from extensive simulations showed no increase in false discovery rate and reinforce isoTWAS’s advantages in prediction and trait mapping power over TWAS, especially when genetic effects on expression vary across isoforms of the same gene. We illustrate multiple biologically-relevant isoTWAS-identified trait associations undetectable by gene-level methods, including isoforms of *AKT3*, *CUL3*, and *HSPD1* with schizophrenia risk, and *PCLO* with multiple disorders.

The isoTWAS framework addresses an unmet need to consider the transcriptome on the transcript-isoform level to increase discovery of trait associations, especially for brain-relevant traits.

## INTRODUCTION

Over the past decade, the number of genetic associations with complex traits identified by genome-wide association studies (GWAS) has increased considerably^1, 2^. However, the slow translation of these genetic associations into concrete molecular mechanisms remains a major obstacle. As GWAS associations predominately localize within non-coding regions of the genome and are often tagged within large blocks of linkage disequilibrium (LD), the first major challenge is prioritizing the underlying causal variant(s) within a given locus and identifying their functional impact on nearby target genes. To address this, numerous methods have been developed to integrate transcriptomic reference panels with GWAS to prioritize genes at trait-associated loci^3–15^. TWAS and related approaches (e.g., PrediXcan) impute the *cis*-component of gene expression predicted by germline genetics into an association cohort, thereby reducing multiple comparisons and increasing interpretability by identifying a set of genes at a locus that may underlie the genetic association^3, 4^. Despite these methodological advances, a majority of GWAS loci still lack robust mechanistic interpretation^16^.

Previous integrative analyses have largely focused on aggregated measurements of total gene expression but have not systematically explored the potentially large number of distinct transcript-isoforms that can be generated from a given genetic locus through alternative splicing. Alternative splicing is a fundamental form of tissue-specific gene regulation present in more than 90% of human genes, that vastly expands the coding and regulatory potential of the genome^17–20;^ GENCODE v40 annotates an average of 4 isoforms per gene (mean 4.05, standard deviation 7.28, median 1)^21^. Genes uniquely expressed in the brain, which are longer and contain far more exons, undergo the greatest degree of splicing compared with other tissues and species—a mechanism contributing to the vast proteomic, phenotypic, and evolutionary complexity of the human brain^22–25^. Some genes are known to express up to hundreds to thousands of unique isoforms in the human brain^25^. Independent of gene expression, splicing dysregulation has been increasingly implicated as a putative disease mechanism^24, 26–30^ including for neuropsychiatric disorders^10, 24, 27, 31^. Yet, local splicing events can be computationally intensive to measure and are difficult to systematically integrate across multiple distinct large-scale datasets. Splicing is often coordinated across a gene, yielding many non-independent features that increases multiple testing burden. In contrast, transcript-isoform abundance can now be rapidly estimated across large-scale RNA-seq datasets using pseudoalignment methods^32, 33^. Furthermore, in the brain, isoform-level expression changes have shown greater enrichment for schizophrenia heritability than gene or local splicing changes^22, 31, 34–36^. However, to fully integrate transcript-isoform quantifications with GWAS, innovative computational methods are needed that jointly model the highly correlated transcripts of the same gene.

Here, we present isoform-level TWAS (isoTWAS), a flexible and scalable approach for complex trait mapping by integrating genetic effects on isoform-level expression with GWAS. In extensive simulations and real data applications in the Genotype-Tissue Expression (GTEx) Project^37^ and the PsychENCODE Consortium^22, 24^, we show that isoTWAS provides several important advantages to trait mapping. First, in the transcriptomic prediction step, the correlation between isoforms provides additional information that is unavailable when only gene-level expression is measured, which can be leveraged to improve prediction^38^ accuracy of individual isoforms in >80% of cases by a median of ∼1.8-2.4-fold improvement and of total gene expression by 25-70%. In parallel, our isoform-centric framework uncovers cross-validated predictive models for ∼2-fold more genes, doubling the number of testable features in the trait mapping step. Third, divergent patterns of genetic effects across isoforms can be leveraged to provide a more granular hypothesis for a mechanism underlying the SNP-trait relationship. Finally, the isoTWAS framework jointly captures expression and splicing disease mechanisms, while maintaining a well-controlled false discovery rate. Altogether, using GWAS data for 15 neuropsychiatric traits, isoTWAS greatly increases the power to detect gene-level trait associations uncovering associations at ∼60% more GWAS loci compared to traditional gene-level TWAS. These results stress the need to shift focus to transcript-isoforms to increase discovery of transcriptomic mechanisms underlying genetic associations with complex traits.

## RESULTS

### The isoTWAS framework

isoTWAS seeks to prioritize genes with transcript-isoforms of genes whose *cis*-genetic component of expression is significantly associated with a complex trait. We extend the traditional gene-level TWAS approach by jointly modeling the expression of distinct transcript-isoforms of a given gene as a matrix while accounting for the pair-wise correlations between these isoforms^21, 37, 39, 40^. Here, we assume that local genetic variants directly modulate expression of an isoform. In addition, we assume that the abundance of a gene is measured as the is the sum of the abundance of its isoforms, computed as transcripts per million, or TPM (**Supplemental Figure S1**)^32, 33, 41, 42^. Flexibly integrating isoform-level expression into trait mapping may lead to novel discoveries in disease mapping and prioritize specific isoforms that explain genetic associations. Accordingly, gene-level trait mapping using traditional TWAS methods may not necessarily detect a trait association on the gene-level if a gene has multiple isoforms but only one is associated with the trait (**Figure 1a**). This scenario may be particularly relevant in the human brain, where some genes may express up to hundreds to thousands of unique isoforms^25^. By modeling the genetic architectures of isoforms of a gene simultaneously, isoTWAS provides a deeper understanding of potential transcriptomic mechanisms that underlie genetic associations.

**Figure 1:**
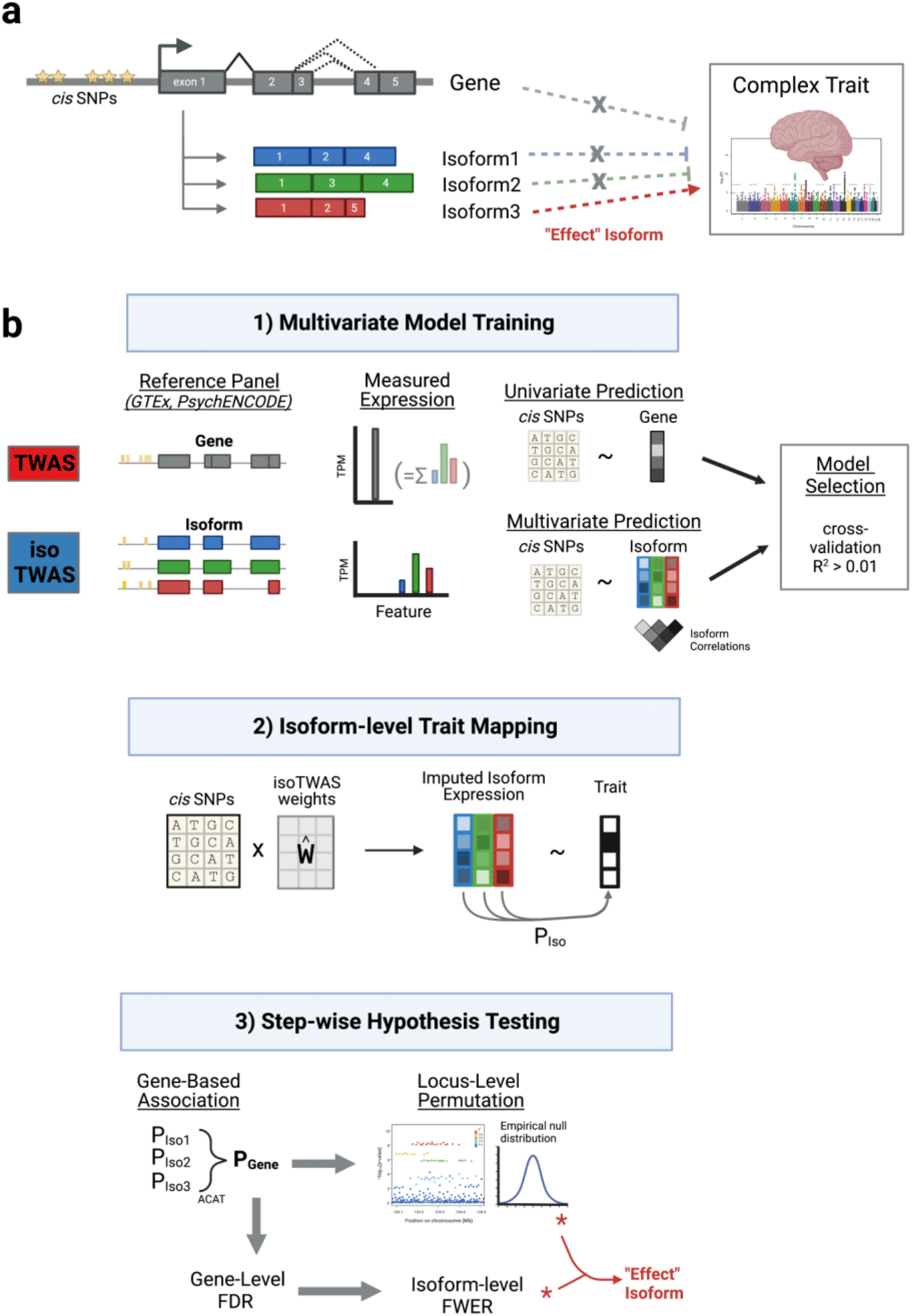
Isoform-centric approach for complex trait mapping and prioritization of disease mechanisms at a genetic locus. **(a)** Motivation for isoTWAS. Gene G has three isoforms associated with a trait but only one has an effect on the trait. Gene G itself does not show an association with the trait. Studying genetic associations with an isoform-centric perspective will prioritize Gene G, but not with a gene-centric perspective. **(b)** Schematic comparison of isoTWAS and TWAS. First, isoTWAS differs from TWAS by training a multivariate model of isoform expression, while TWAS models total gene expression, the sum of isoform expression. Second, isoTWAS maps isoform-trait associations through a step-wise hypothesis testing framework that provides gene-level false discovery control and isoform-level family-wide error rate control. TWAS only maps gene-trait association.

The isoTWAS framework contains three general steps (**Figure 1**). First, we build multivariate predictive models of isoform-level expression using well-powered functional genomics training datasets, including GTEx^37^ and PsychENCODE^24, 27^. Here, we trained and systematically compared 4 multivariate predictive frameworks: (1) multivariate elastic net penalized regression^43^, (2) multivariate LASSO penalized regression with simultaneous covariance estimation (MRCE)^44^, (3) multivariate elastic net regression with stacked generalization (joinet)^45^, and (4) sparse partial least squares (SPLS)^46^. As a baseline for comparison, we also modeled each individual isoform independently with univariate regularized regressions, as implemented in Gusev et al’s FUSION software^4, 43, 47, 48^ (see **Methods** and **Supplemental Methods**). Models were trained to predict isoform expression using the set of *cis*-SNPs within 1 Megabase (Mb) of the gene body (**Methods**, **Figure 1b**). Model performance was assessed via 5-fold cross-validation, using McNemar’s adjusted R^2^ between observed and predicted expression.

Next, we use these models to impute isoform expression into an external GWAS cohort and quantify the association with the target GWAS phenotype (**Figure 1b**). If individual-level genotypes are available, isoform expression can be directly imputed as a linear combination of the SNPs in the models, and these associations can be estimated through appropriate regression analyses. If only GWAS summary statistics are available, imputation and association testing is conducted simultaneously through a weighted burden test^4^. Finally, isoTWAS performs step-wise hypothesis testing procedure to account for multiple comparisons and control for local LD structure. Isoform-level P-value are first aggregated to the gene-level to prioritize a gene using the Aggregated Cauchy Association Test (ACAT)^49^, where false discovery rates are controlled, and then individual isoforms of prioritized genes are subjected to post-hoc family-wise error control^50^ (**Supplemental Figure S2**, **Methods**). After this step, a set of isoforms are identified whose *cis*-genetic component of expression are associated with the trait of interest^4^. For these isoforms, we apply a rigorous permutation test whereby the SNP-to-isoform effects are permuted 10,000 times to generate a null distribution; this permutation test assesses how much signal is added by isoform expression, given the GWAS architecture of the locus, and controls for large LD blocks^4^. Lastly, we can isoform-level Bayesian fine-mapping at each locus with a significant trait association to identify the minimal credible set of isoforms that contains the ‘casual’ isoform at a 90% confidence level and to assign individual posterior inclusion probabilities (**Figure 1b**, **Methods**). isoTWAS is available as an R package through Github (https://github.com/bhattacharya-a-bt/isotwas).

### isoTWAS improves prediction of isoform and gene expression in simulations and real data

Previous work has demonstrated that isoform-level quantifications, when propagated to the gene-level, can lead to more accurate gene expression estimates and differential expression results with short read RNA-seq data^41, 42^. We therefore hypothesized that our multivariate SNP-based imputation of isoform expression, when summed to the gene-level, would outperform traditional gene-level TWAS models. To systematically evaluate the performance of TWAS versus isoTWAS models on prediction of total gene expression across a variety of genetic architectures, we conducted an extensive set of simulations across 22 different gene loci using European-ancestry data from the 1000 Genomes Project^51^ (**Methods, Figure 2a**). At each gene locus, we controlled gene expression heritability and simulated 2-10 distinct isoforms, varying the proportion of causal isoQTLs (p_causal_) and their sharing between isoforms (p_shared_). We then trained cross-validated, multivariate predictive models of isoform expression (isoTWAS) or univariate models of gene expression (TWAS). For isoTWAS, of the specific multivariate prediction models tested, multivariate elastic net^43^ demonstrated the greatest CV prediction of isoform expression across most simulation settings (**Figure 2b**, **Supplemental Figure S3, Supplemental Data 1**). For gene expression prediction, the optimal isoTWAS models (in sum) outperformed the optimal TWAS model, particularly at sparser isoQTL architectures, with median absolute increase in adjusted R^2^ of 0.6-3.5% (**Figure 2c**, **Supplemental Figure S4, Supplemental Data 2**). Performance gains decreased somewhat with denser isoQTL architectures, although real data is consistent with 0.1-1% sparsity (i.e., 1-10 causal e- and isoQTLs per gene or isoform)^37^. In simulations, we found that isoTWAS prediction of gene expression also increases as the proportion of shared non-zero effect SNPs across isoforms decreases (**Figure 2b-c, Supplemental Figure S4, Supplemental Data 2**).

**Figure 2:**
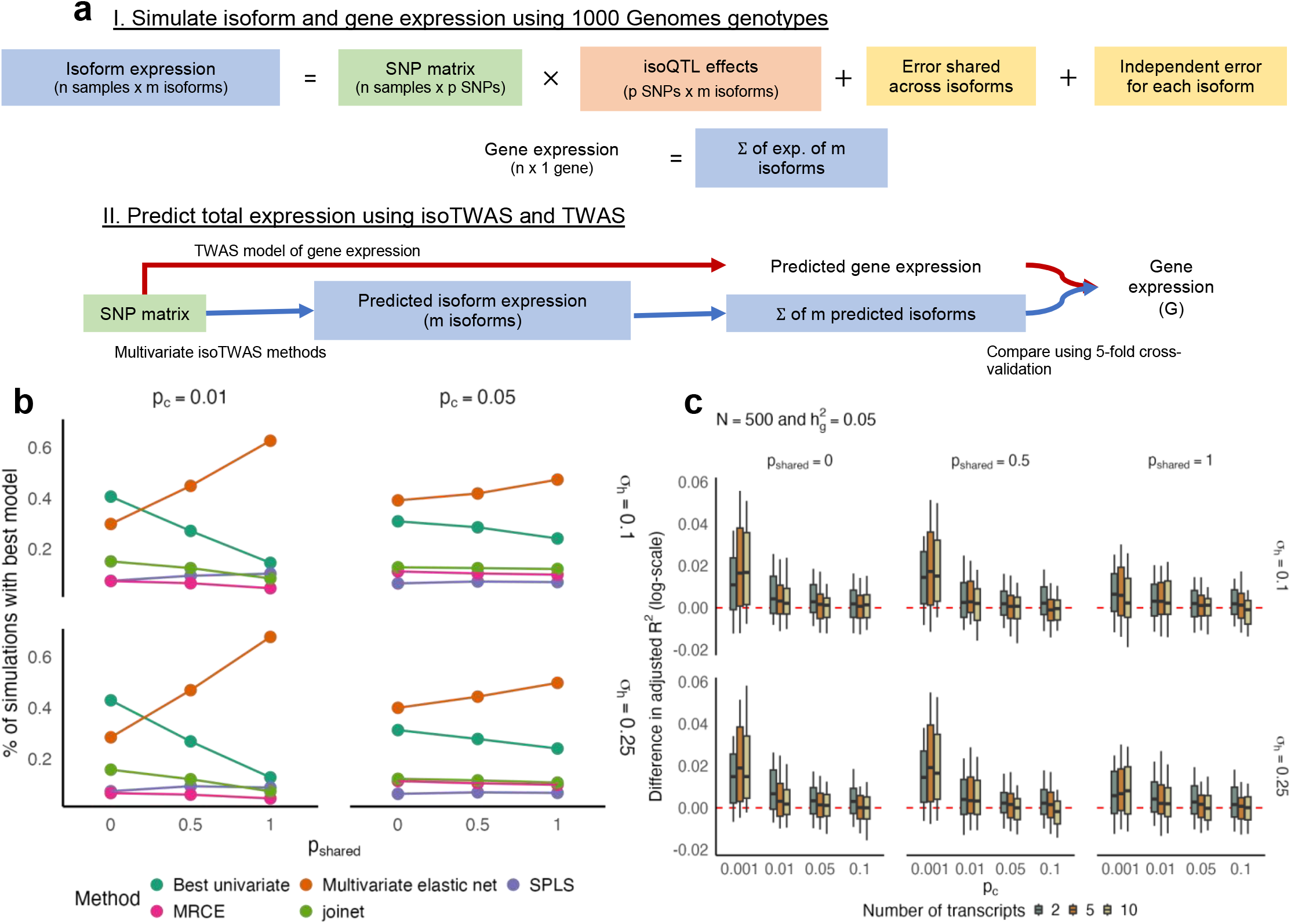
IsoTWAS models predict gene expression with more accuracy than TWAS models in simulated data. **(a)** Simulation setup to generate isoform expression with specified isoQTL architecture, controlled expression heritability, number of isoforms, and inter-isoform correlation structure. **(b)** Proportion of simulations where the isoTWAS model has the maximum adjusted R^2^ for marginal isoform prediction (Y-axis) across shared isoQTL proportion (X-axis), colored by isoTWAS method, facetted by causal isoQTL proportion (top margin) and proportion of isoform expression variance attributed to shared non cis-genetic effects (right margin). **(c)** Boxplots of difference in adjusted R^2^ in predicting gene expression between isoTWAS and TWAS models from simulations with sample size 500 where isoform and gene expression heritability are set to 0.05, across causal isoQTL proportion (X-axis) and colored by number of transcripts per gene, facetted by proportion of shared isoQTLs (top margin) and proportion of variance explained by shared non cis-genetic effects (right margin).

Next, we assessed predictive performance in real data from 48 tissues (13 brain) with sufficient sample sizes (N > 100) in GTEx for all genes with multiple expressed isoforms (**Supplemental Table S1**; **Methods**). Altogether, for 48 tissues in GTEx, we built significant predictive models for 50,000 to 80,000 isoforms across 8,000 to 12,000 unique genes per tissue (**Supplemental Figure S11, Supplemental Table S2**). We considered 3 main criteria to evaluate the performance of both the multivariate and isoform-centric approaches of isoTWAS: (1) the number of isoforms whose expression can be imputed using multivariate/univariate models with cross-validation (CV) R^2^ > 0.01; (2) the number of unique genes with at least one isoform that can be imputed at CV R^2^ > 0.01; and (3) the number of unique genes in which total gene expression can be imputed at CV R^2^ > 0.01 using isoTWAS (summed) or TWAS models.

At the isoform level (criterion 1), through joint multivariate modeling of isoform expression, we trained 2.3-2.5-fold more models at cross-validation (CV) R^2^ > 0.01 across the 48 tissues, compared to traditional univariate approaches (**Figure 3a**, **Supplemental Figure S5**). Using these multivariate models, we improve prediction for 79-82% of isoforms with a median increase of ∼1.8-2.4 fold increase in adjusted R^2^ (**Supplemental Figure S6, Supplemental Table S2**). Concordant with simulations, we found that the multivariate elastic net overwhelmingly outperformed other multivariate (and univariate) methods, indicating that leveraging the shared genetic architecture between isoforms of the same gene greatly aids in prediction of each individual isoform (**Supplemental Figure S7, Supplemental Table S2**). Notably, we observed that multivariate models were particularly important for brain tissues compared with non-brain tissues in GTEx, which showed significantly improved performance compared with univariate models (**Figure 3b**; P = 0.011 from OLS regression of median percent increase in CV R^2^ for multivariate/univariate models against tissue type, adjusted for sample size), suggesting more shared isoQTL architecture in brain tissues than others which can be leveraged by isoTWAS for improved prediction. These gains in prediction accuracy directly translate into increased power in the trait association step^52^.

**Figure 3:**
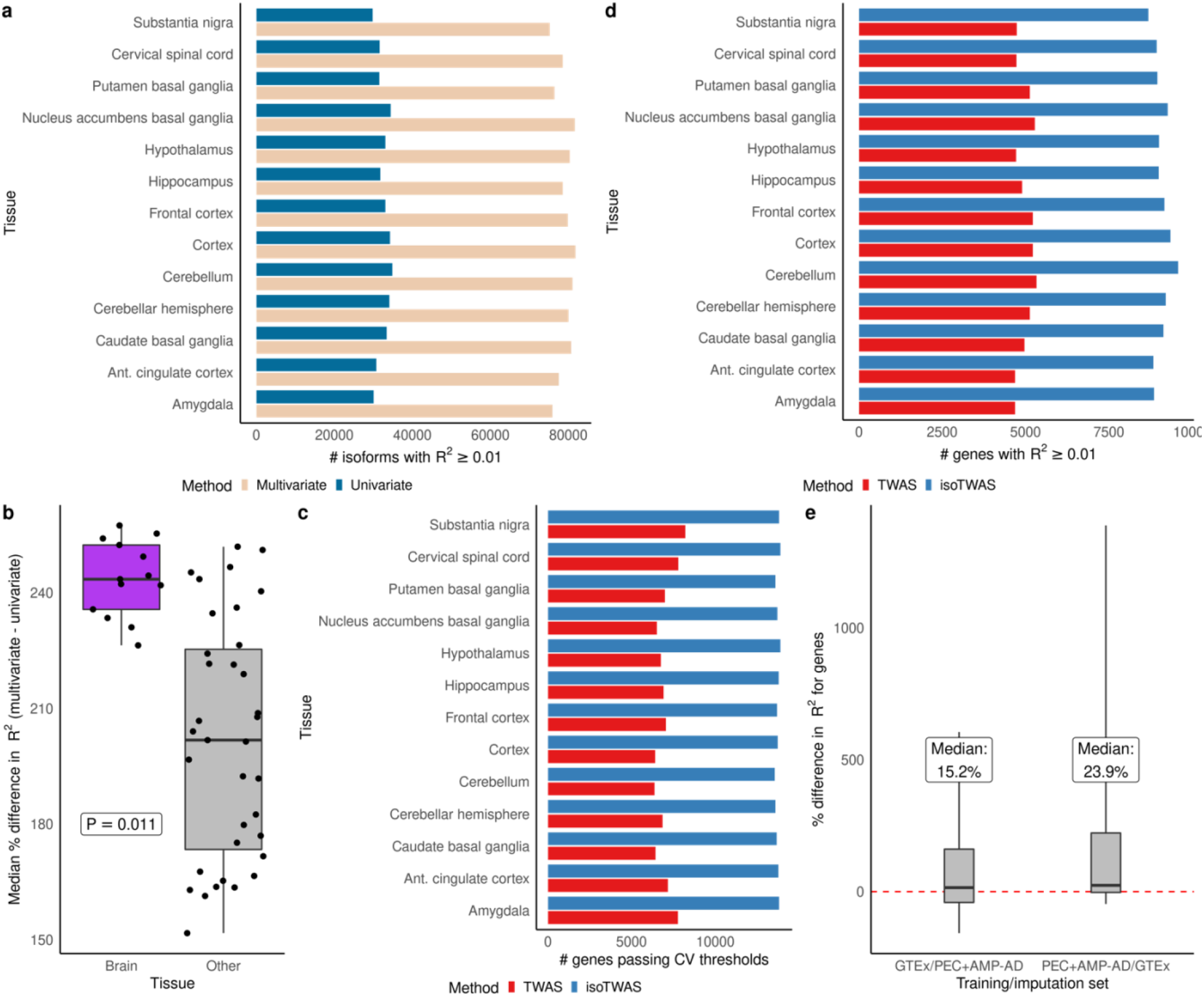
Multivariate isoform-level predictive models improve upon gene-level predictive models in predictive gene-level expression. **(a)** Barplot showing the number of isoforms with CV R^2^ > 0.01 (Y-axis) using multivariate (cream) and univariate (blue) modelling methods across brain tissues (X-axis). **(b)** Boxplot of median percent difference in predicting isoform expression (Y-axis) using multivariate compared to univariate method by brain and other tissue (X-axis). P-value corresponds to difference in median across groups, adjusted for sample size. **(c)** Barplot showing the number of genes passing CV thresholds (Y-axis) using TWAS (red) and isoTWAS (blue) across brain tissues. **(d)** Barplot showing the number of genes with CV R^2^ > 0.01 (Y-axis) using TWAS (red) and isoTWAS (blue) across brain tissues (X-axis). **(e)** Boxplot of percent difference in R^2^ (Y-axis) of predicting gene expression (isoTWAS – TWAS) in external datasets. X-axis shows the training and imputation datasets. The median percent difference in labelled.

At the gene level (criteria 2 and 3), isoTWAS also increased the number of genes with testable models in the trait mapping step and improved prediction of total gene expression. The number of unique genes with at least 1 isoTWAS model at CV R^2^ > 0.01 (inclusion criterion for isoTWAS trait mapping) was 1.9-2.5 times larger than the number of unique genes with TWAS models achieving CV R^2^ > 0.01 for gene expression prediction (**Figure 3c, Supplemental Figure S8, Supplemental Table S2**). For a given gene, isoTWAS models (summed) outperformed TWAS models in prediction of total gene expression by a median of 25-70% in cross validation (**Supplemental Figure S9**) with a 50-80% increase in the number of genes that are predicted at CV R^2^ > 0.01 (**Figure 3d, Supplemental Figure S10**). We replicated these gains in total gene expression prediction using an independent, out of sample QTL reference panel of adult cortex from PsychENCODE/AMP-AD (Methods). Multivariate isoTWAS models outperformed univariate TWAS models in predicting total gene expression with a 15.2% median percent increase in adjusted R^2^ when training in GTEx and testing in PsychENCODE/AMP-AD, and 23.9% vice versa; **Figure 3e**, **Supplemental Table S3**). As predictive performance is positively related to power to detect trait associations^51^, both the increased number and accuracy of trainable imputation models using isoTWAS’s multivariate predictive framework have strong implications on increased discovery in trait mapping^51^.

### Performance of isoTWAS across distinct genomic contexts

As genes can differ widely with respect to the number and expression patterns of their constituent isoforms, as well as by other potentially relevant features such as gene length and SNP density, we next sought to characterize the impact of these factors of isoTWAS performance using GTEx models as evaluated by the 3 criteria outlined above (**Methods, Supplemental Note, Supplemental Figures S12-19, Supplemental Data 3-4**). Overall, we observed an increase in performance of isoTWAS multivariate modeling of both isoforms and genes with increasing number of isoforms per gene, although there was less conclusive of a pattern with increasing dominant isoform fraction^52^ (**Supplemental Figures S12-13, Supplemental Note, Methods**). We also noticed trends in the performance gain using isoTWAS multivariate modeling with respect to both isoform and gene expression prediction across gene length, SNP density at the gene locus, and sample size (**Supplemental Figures S14-16**, **Supplemental Note**). Finally, as the proportion of non-zero effect SNPs in the isoTWAS model that are shared across isoforms increased (**Supplemental Note**), we found an increasing trend in the gain in prediction of gene expression using isoTWAS compared to TWAS models (**Supplemental Figure S17**), reflecting a similar observation from simulation. Interestingly, as this proportion increased, we found an increase in the gain in prediction of isoform expression, reinforcing the utility of multivariate modelling in marginal prediction of isoform expression.

Lastly, we investigated how the robustness of isoform abundance estimation from short-read RNA-seq impacted performance gains of isoTWAS, compared with TWAS. Isoform abundance was initially quantified from probabilistic point estimates using Salmon, guided by Gencode annotations. We then assessed the performance of isoTWAS across loci binned by quantification variance measured across 50 inferential replicates from Salmon^32^. In general, we found that for isoform-level prediction, multivariate modeling in isoTWAS substantially outperformed univariate approaches as quantification variance increased. However, comparing isoTWAS isoform-centric and TWAS gene-centric models, there were no discernable trends in prediction of gene expression as the mean count and/or quantification variance of genes increased (**Supplemental Figure S18-19**, **Supplemental Note**). Finally, we evaluated the impact of reference transcriptome annotation fidelity by generating a synthetic dataset quantified using a reference annotation masking the dominant isoforms for a set of genes. As expected, performance of both isoTWAS and TWAS models declined when isoforms failed to be detected in expression quantification (**Supplemental Note**, **Supplemental Figure S20**).

### Simulations support improved power and calibrated null across genetic architectures

We next introduced GWAS data for complex traits into our simulation framework to benchmark the false positive rate (FPR) and power of isoTWAS (**Methods**). First, we found that the FPR is controlled at 0.05 for isoform-level mapping using ACAT, consistent with gene-level mapping using TWAS (**Supplemental Figure S21, Supplemental Data 4**). For a simulated trait, we modeled causal effect architectures for a genomic locus with 2-10 isoforms under three main scenarios (**Methods**; **Figure 4, Supplemental Figure S22**): (1) where the true trait effect is from only total gene expression, (2) where there is only one isoform with a non-zero effect on the trait, called the “effect isoform”; and (3) where 2 isoforms are effect isoforms, with varying magnitudes and directions of association. This first scenario showed clear increases in power for TWAS over isoTWAS, but this advantage declined as the causal proportion of isoQTLs increased and as the proportion of shared isoQTLs increased (**Figure 4a, Supplemental Data 5**). For scenarios (2) and (3), as effects on the trait varied across isoforms of the same gene (**Figure 4b-c, Supplemental Data 6-7**), we observed large increases in power for isoTWAS over TWAS. Here, across most scenarios and causal effect architectures, isoTWAS demonstrated improved power compared with gene-level TWAS, particularly when only one isoform of a gene carried a trait effect or when two effect isoforms of the same gene had different directions of effects. However, when the effect sizes of two effect isoforms of the same gene converged, TWAS and isoTWAS demonstrated similar power to detect a gene-trait association (**Figure 4c**).

**Figure 4:**
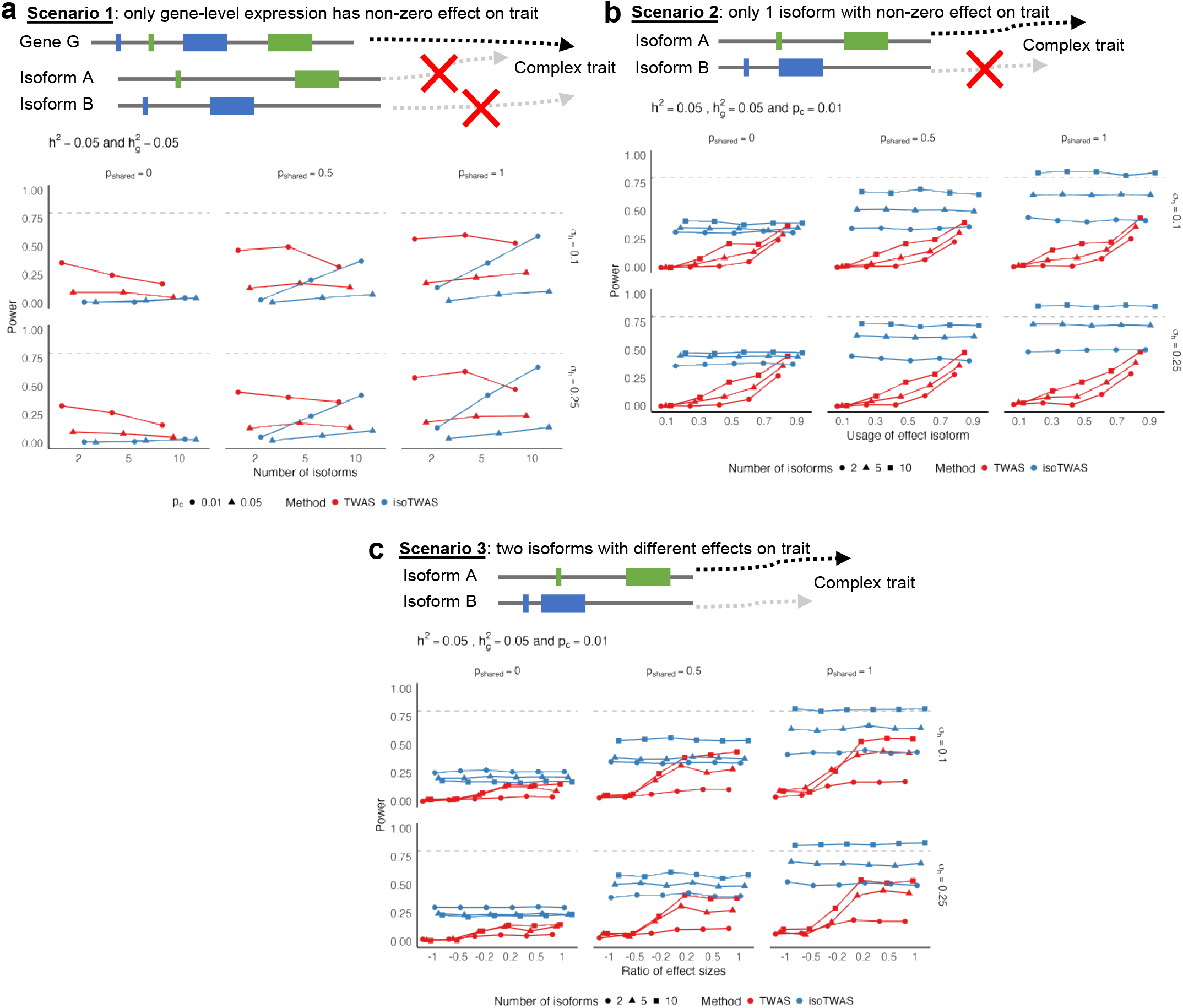
IsoTWAS improves power to detect gene-trait associations, especially when genetic effects differ across isoforms, in simulations. **(a)** Power to detect gene-trait association (proportion of tests with P < 2.5 × 10^-6^, Y-axis) across causal proportion of isoQTLs (X-axis), facetted by proportion of shared isoQTLs (top margin) and proportion of variance explained by shared non cis-genetic effects (right margin). **(b)** Power to detect gene-trait association (proportion of tests with P < 2.5 × 10^-6^, Y-axis) across proportion of gene expression explained by effect isoform (X-axis). **(c)** Power to detect gene-trait association (proportion of tests with P < 2.5 × 10^-6^, Y-axis) across ratio of effect sizes for 2 effect isoforms. Across all plots, isoform and total gene expression heritability is set to 0.05 and causal proportion of 0.01 in **(b-c)**. Points are colored by method and shaped by the number of isoforms per gene.

Finally, we assessed the performance of probabilistic fine-mapping in identifying the true effect isoform in our simulation framework of genes with 5 or 10 isoforms (**Methods, Supplemental Figure S23, Supplemental Data 9**). In general, the sensitivity of 90% credible sets (proportion of credible sets that contained the true effect isoform) was under-calibrated, likely due to difficulties in fine-mapping when QTL horizontal pleiotropy is extremely high^53^. We found that the sensitivity of 90% credible sets decreased and the mean set size increased with increasing proportion of shared isoQTLs. Our simulation results suggest that varied isoQTL architectures and isoform-trait effects for isoforms of the same gene are key features that influence power gains in isoform-centric modeling.

### isoTWAS increases discovery of trait associations across 15 neuropsychiatric disorders

To explore our central hypothesis that isoform-centric multivariate predictive modeling will improve discovery for complex trait mapping, particularly for brain relevant traits, we next sought to apply and directly compare isoTWAS and TWAS results across 15 neuropsychiatric disorders and traits. To maximize discovery, we trained both isoTWAS and TWAS models using a greatly expanded adult brain functional genomics reference panel (N = 2,115), comprised of frontal cortex samples from PsychENCODE and AMP-AD Consortia^27, 54^, as well as using a developmental^55^ pre-frontal cortex (N = 205) dataset (**Methods; Figure 5**). In the adult cortex, we trained models for 15,127 genes using isoTWAS passing the CV R^2^ > 0.01 cutoff, compared with 14,283 genes using gene-level TWAS. In the developing cortex, despite a smaller sample size, isoTWAS models were successfully trained for 16,504 genes, compared with 10,535 genes using TWAS (**Methods; Supplemental Table S1**).

**Figure 5:**
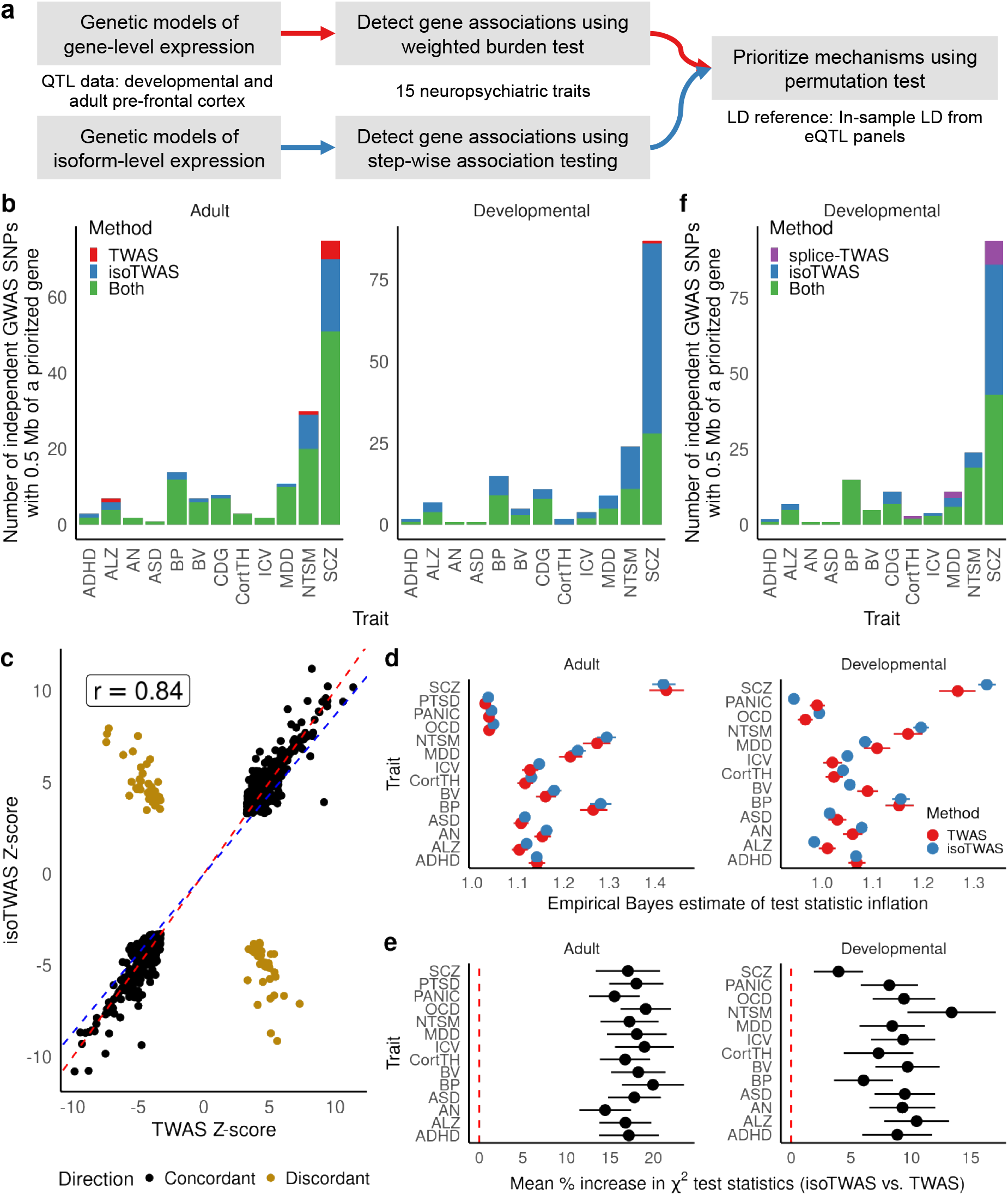
Isoform-level trait mapping increases discovery of genetic associations over gene-level trait mapping. **(a)** Schematic diagram for trait mapping using gene-level TWAS and isoTWAS using PsychENCODE data. **(b)** Number of gene-trait associations overlapping GWAS risk SNPs within 0.5 Mb using gene-level TWAS (red), isoTWAS (blue), or either (green) at conservative permutation-based significance thresholds. **(c)** Scatterplot of standardized effect sizes (Z-scores) using isoTWAS and gene-level TWAS, considered associations with nominal P < 0.05 using both TWAS and isoTWAS. Gray line shows the 45-degree line and the green line shows an ordinary least squares regression. **(d)** Empirical Bayes estimate of test statistic inflation (X-axis) for TWAS (red) and isoTWAS (red) gene-level associations across 15 traits (Y-axis). **(e)** Mean percent increase in approximate *X*^2^-test statistic (squared Z-score), which is proportional to increase in effective sample size, for isoTWAS trait associations over TWAS trait associations. **(f)** Number of gene-trait associations overlapping GWAS risk SNPs within 0.5 Mb using splicing-event-based TWAS (purple), isoTWAS (blue), or either (green) at conservative permutation-based significance thresholds.

We next applied these models to perform trait associations using summary statistics from 15 neuropsychiatric and brain-related GWAS^56–70^ (**Methods, Figure 5a, Supplemental Figure S24**), using the conservative stepwise hypothesis testing procedure (**Figure 1a**; FDR-adjusted P < 0.05 and within-locus permutation P_ACAT_\< 0.05). We detected far more trait-associated genes with isoTWAS compared with TWAS, across adult (2,595 vs 1,589 genes) and developmental (4,062 vs 890 genes) reference panels, respectively (**Supplemental Figure S25, Supplemental Data 10-13**). In total across both reference panels and all 15 traits, isoTWAS detected 3,436 unique gene and 5,377 unique isoform-trait associations (**Supplemental Figure S26**). In addition, of the 1,335 genes with multiple isoform-trait associations, 661 gene exhibited distinct isoform-level associations in different directions.

We next sought to compare the performance of isoTWAS/TWAS in prioritizing candidate mechanisms within independent, high-confidence GWAS-significant loci^73^. Across a combined 1,149 GWAS loci, isoTWAS identified significant associations within 323, compared with 201 detected by TWAS, a ∼60% increase in discovery (**Figure 5b**; **Methods, Supplemental Table S4**). For example, of the 287 GWAS loci identified for SCZ^74^, isoTWAS prioritized genes within 70 and 86 unique loci across adult and developmental reference panels, respectively, compared with 56 and 29 loci for TWAS (**Figure 5b**). Further, across the 15 traits, 96% of gene-level TWAS associations (193/201) were concordantly identified by isoTWAS. Likewise, the standardized effect sizes for significant gene- and isoform-level associations were highly correlated (r = 0.84, P < 2.2 × 10^-16^; **Figure 5c**). Finally, to explore whether these isoTWAS-specific associations were capturing true disease signal, we compared the rate at which each method prioritized constrained genes (probability of loss-of-function intolerance, pLI ≥ 0.9; **Supplemental Tables S5-S8**), which are known to be substantially enriched for disease associations^75^. Across adult and developmental panels for 15 traits, respectively, isoTWAS prioritized 724 and 385 constrained genes compared with 106 and 200 with TWAS, a significant increase (adult: P=0.048, developmental: P = 1.23 × 10^-5^, Fisher’s Exact test). Altogether, these results emphasize that isoTWAS not only recovers the vast majority of TWAS associations but also greatly increases discovery of candidate GWAS mechanisms, particularly for genes intolerant to protein-truncating variation^76^.

In our evaluation of methods in real data, we sought to compare discovery using isoTWAS to discovery using local splicing-event based trait mapping. Briefly, for the developmental brain dataset, we first calculated intron usage using Leafcutter^77^ and transformed these usage percentages to M-values^78^. Then, for all introns mapped to a given gene, we used all SNPs within 1 Mb of a splicing-event to predict its usage and mapped trait associations for these splicing events using isoTWAS’s multivariate framework (**Methods**). Overall, when aggregated to the gene-level, across 15 traits, we found that isoTWAS prioritized features at ∼40% more independent GWAS loci (167 loci) than splicing-event based trait mapping (119 loci), with 108 loci (90.7%) jointly identified (**Figure 5f**), using the same developmental brain reference panel. Taken together, isoTWAS’s specific focus on modeling isoforms of a gene provided considerable gains over considering only total gene expression or intron usage in identifying trait associations for genes and their transcript-isoforms.

To investigate whether this increase in trait mapping discovery reflected true biological signal, rather than test statistic inflation due to the increased number of tests (∼4-fold increase in number of tests), we next compared the null distributions across methods for results across the 15 traits (**Supplemental Figure S27).** As the genomic inflation factor is not a reliable measure in TWAS settings^79^, we estimated inflation in gene-level test statistics using an empirical Bayes approach (**Methods**). Collapsing across all 15 traits, there were no significant differences between TWAS and isoTWAS in the 95% credible intervals for test statistic inflation (**Figure 5d**). Using a heuristic to estimate increases in effective sample size (**Methods**), we observed an approximate increase in effective sample size of 10-20% when using isoTWAS compared to TWAS (**Figure 5e, Supplemental Table S9**). These analyses indicate that isoTWAS discovery is both well-calibrated to the null and facilitates increased discovery in real data compared to gene-level TWAS.

We also empirically compared probabilistic fine-mapping^55^ of results from isoTWAS and gene-level TWAS (**Methods**). Here, we conducted fine-mapping on significant trait-associated genes/isoforms (adjusted P < 0.05 and permutation P < 0.05) and are within 1 Mb of one another; we term a locus with overlapping genes/isoforms a risk region. Overall, the mean number of genes in a risk region using TWAS was 3.15 compared to 3.90 using isoTWAS (**Supplemental Figure 28a**); the mean number of genes in a 90% credible set using TWAS was 1.33 compared to 1.25 using isoTWAS (**Supplemental Figure 28a**). On average, there were 1.54 isoforms per gene in a risk region and 1.27 isoforms per gene in a 90% credible set (**Supplemental Figure 28b**). Isoform-centric modeling presents unique challenges for fine-mapping due to potentially high levels of horizontal pleiotropy, and remains an important and open question for the field. Nevertheless, isoTWAS identified a relatively comparable number of genes in risk regions compared with TWAS, and the combination of conservative two-step trait mapping, permutation testing, and probabilistic fine-mapping were critical for maintaining a narrow credible set size.

### isoTWAS identifies biologically-meaningful trait associations undetectable by TWAS

Overall, isoTWAS prioritized dozens of novel candidate risk genes and mechanisms in the developing and adult brain for 15 neuropsychiatric traits. These isoTWAS-prioritized genes were enriched for multiple relevant pathways consistent with the biology of the underlying trait, including cell proliferation pathways for brain volume, calcium channel activity for schizophrenia and neuroticism, and proteasome regulation in Alzheimer’s Disease, among others (**Supplemental Figure S29**). We highlight below several examples of novel trait associations in which isoTWAS prioritized a highly constrained gene within a GWAS locus (**Supplemental Table S5-8**). For example, we identified an association for SCZ with developmental expression of ENST00000519133, an isoform of *SNAP91* (adjusted P = 6.06 × 10^-7^ in isoTWAS, pLI = 0.99, chromosomal location of 6q14.2), with a GWAS-significant variant rs217291 within the gene body. *SNAP91* has been predicted to affect clathrin and phosphatidylinositol biding activity and synaptic vesicle recycling and has previously shown to impact synaptic development^80^. In addition, we found that imputed developmental cortex expression of ENST00000476671, an isoform of *KMT2E*, was associated with CDG risk (adjusted P = 7.63 × 10^-3^, pLI = 1, chromosomal location of 7q22.3); the GWAS SNP rs2385537, associated in a meta-analysis of ADHD, ASD, BP, and SCZ^81^, is within the gene body. *KMT2E* regulates post-translational histone methylation of histone 3 on lysine 4A, and *KMT2E* heterozygous variants are associated with risk of neurodevelopmental disorders^82^. Imputed adult brain expression of ENST00000492146 (adjusted P = 1.14 × 10^-6^, pLI = 0.94, chromosomal location of 3p21.1), an isoform of *SFMBT1*, showed a strong association with SCZ risk and contains a GWAS SNP within the gene body (rs2071044). In a recent gene-based analysis of GWAS data for SCZ and BD, decreased expression of *SFMBT1* was associated with increased risk of both disorders^83^, consistent with the effects for the isoform we identify in this isoTWAS analysis. Lastly, our analysis implicated ENST00000537270, an isoform of *KMT5A* (pLI = 0.99, found in 12q24.31), in an association with SCZ risk. *KMT5A*, a H4K20 methyltransferase, has been previously implicated in GWAS for SCZ but cis-eQTLs of the gene have not be colocalized with the GWAS signal previously^74, 84, 85^.

As evidenced by our simulation analyses, a main advantage of isoTWAS over TWAS is the identification of trait associations for isoforms of genes, where the gene itself is not associated with the trait. We illustrate several examples of isoforms prioritized by isoTWAS, all in adult cortex, for genes with limited or distinct expression QTLs (**Figure 6, Supplemental Figure S30, Supplemental Data 14**). First, we detected a strong SCZ association with ENST00000492957, an isoform of *AKT3* (pLI = 1) at 1q43-q44, which encodes a serine/threonine-protein kinase that regulated many processes like metabolism and cell growth, proliferation, and survival. *AKT3* has shown effects on anxiety, spatial and contextual memory, and fear extinction in mice, such that loss-of-function of *AKT3* causes learning and memory deficits^86, 87^. Within the GWAS locus, there was a strong overlapping isoQTL signal (P < 10^-50^); however, the eQTL signal includes only a single SNP that reaches P < 10^-6^, which is in low LD with the GWAS-significant SNPs in the locus (**Figure 6a**). The lead isoQTL (rs4430311) showed a significant, negative association with ENST00000492957, but not *AKT3* expression, with increasing SCZ risk alleles. However, this SNP has only a nominally significant positive association with *AKT3*, as the number of alternative alleles at the SNP increased. Interestingly, a different isoform of *AKT3* (ENST00000681794) was prioritized in an association with BV, which also has a GWAS hit at this locus (**Supplemental Figure S30**). The two distinct isoforms of *AKT3* have distinct 3’ transcript structures, particularly close to the lead isoQTL of ENST00000681794. These results suggest a complex role of *AKT3* isoforms with brain-related traits to be explored further computationally and experimentally.

**Figure 6:**
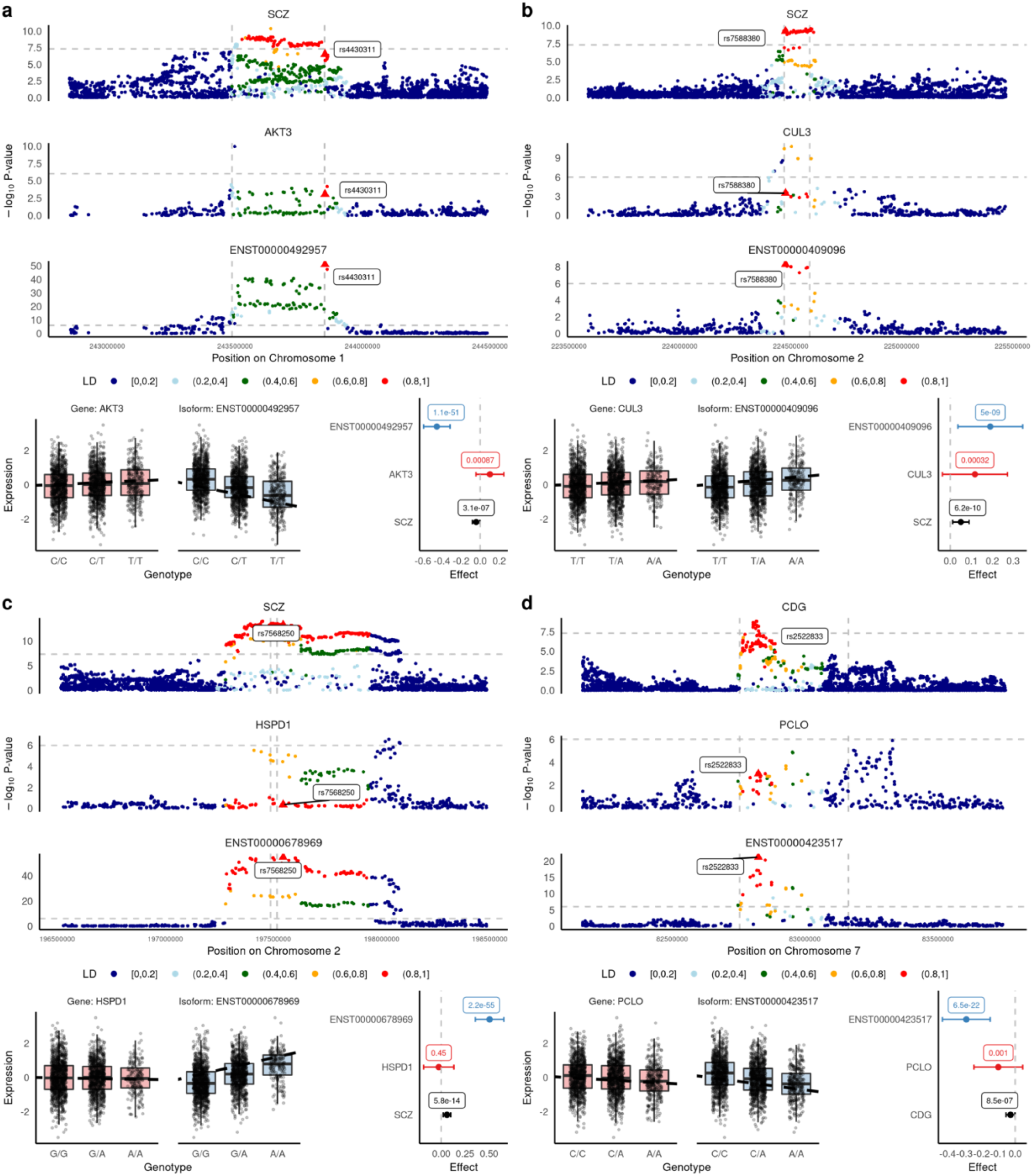
isoTWAS implicates isoforms of AKT3, CUL3, HSPD1, and PCLO in genetic associations with psychiatric traits. In each plot, (top) Manhattan plots of GWAS risk, total gene eQTLs, and isoQTLs, colored by LD to the lead isoQTL and lead isoQTL shown in a triangle and labelled. LD is based on the 1000 Genomes European reference. (bottom) Boxplots of gene and isoform expression by genotype of the lead isoQTL SNP and forest plot of the lead isoQTL’s effect and 95% confidence interval on the trait, gene, and isoform, with P-values labelled. **(a)** SCZ risk, *AKT3* gene expression, and ENST00000492957 isoform expression. **(b)** SCZ risk, *CUL3* gene expression, ENST00000409096 isoform expression. **(c)** SCZ risk, *HSPD1* gene expression, ENST00000678969 isoform expression. **(d)** CDG risk, *PCLO* gene expression, ENST00000423517 isoform expression.

Similarly, we found a strong isoQTL signal for ENST00000409096 but a weak eQTL signal of *CUL3* in the 2q36.2 locus (pLI = 0.99), in another association with SCZ (**Figure 6b**). *CUL3* is a component of Cullin-RING E3 ubiquitin ligase complexes, involved in protein ubiquitylation, cell cycle regulation, protein trafficking, signal transduction, and transcription. Previous work has implicated *CUL3* dysregulation as a pathological mechanism for both SCZ and ASD risk^88^. Next, an isoform ENST00000678969 of *HSPD1*, encoding a mitochondrial heat shock protein, was associated with SCZ risk (adjusted P = 8.79 × 10^-12^, pLI = 0.99, 2q33.1) and showed a similar pattern across GWAS, eQTL, and isoQTL signals (**Figure 6c**). *HSPD1* was identified in 3 independent secondary analyses of SCZ GWAS data, is among multiple non-MHC immune genes implicated in SCZ, and has roles in brain hypomyelination^89^. Our work adds to these previous studies by implicating a specific isoform of *HSPD1* at the locus. Lastly, ENST00000423517, an isoform of *PCLO*, was associated with multiple traits in the CDG analysis (meta-analysis of ADHD, BP, MDD, and SCZ; adjusted P = 1.83 × 10^-4^, pLI = 1). Again, we found a strong isoQTL but not eQTL signal, with the CDG risk allele negatively associated with isoform expression. *PCLO* is involved in the presynaptic cytoskeletal matrix, establishing active synaptic zones, and synaptic vesicle trafficking; rare variants of *PCLO* in diverse populations have been recently implicated in risk of SCZ and ASD^90, 91^. Altogether, these results highlight the importance of incorporating isoform-level regulation for prioritizing novel candidate GWAS risk mechanisms, as implemented in our isoTWAS framework.

## DISCUSSION

In this work, we introduce isoTWAS, a scalable framework that integrates genetic and isoform-level transcriptomic variation with GWAS to not only identify genes whose expression are associated with a trait but also prioritize a set of isoforms of the gene that best explains the association. We provide an extensive set of isoform-level predictive models (https://zenodo.org/record/679594792) and software to conduct isoform-level trait mapping with GWAS summary statistics (https://github.com/bhattacharya-a-bt/isoTWAS).

As demonstrated above, isoTWAS presents several advantages over traditional gene-level TWAS or simple univariate modeling of isoform expression. First, modeling expression at the isoform-level can detect isoQTL architectures that vary across isoforms and thus may not be captured by eQTLs of a gene. Second, joint multivariate isoform-level modeling improved predictive accuracy of isoform and total gene expression, both in simulated and real data with independent replication. Third, aggregating isoform-level associations to the gene-level through ACAT substantially increased power to detect trait associations over traditional TWAS. We attribute this increase in power to several key features: first, isoform-level modeling in isoTWAS increases the number of imputable genes by >2-fold; second, isoTWAS models improve accuracy of gene-level prediction by as much as 35%; third, isoTWAS jointly models both expression and splicing regulation, thereby capturing multiple potential underlying complex trait mechanisms. Finally, as genetic control of isoform expression and usage are often much more tissue- and cell-type-specific than eQTLs^26, 37^, we hypothesize that isoTWAS is more capable of uncovering such context-specific trait associations.

Recent work has highlighted the promise of alternative splicing as a biological mechanism underlying complex traits not fully captured through eQTLs^24, 26, 27, 93^. Splicing-QTL integration is a promising vehicle to prioritize alternative exons or splice sites that may explain the genetic association with a trait, but a single exon or splice site can correspond to multiple isoforms of the same gene. Mapping genetic regulation at the exon-rather than gene-level often leads to more detected signal^94^. However, most of these analyses have focused on local splicing events or exon-level inclusion, rather than the combined consequences of these events – namely, different isoforms of the same gene. Local splicing events can be computationally intensive to measure and are difficult to systematically integrate across multiple distinct large-scale datasets, which is necessary for achieving sample sizes sufficient for interrogation of population-level allelic effects^27, 28^. Furthermore, multiple splicing changes are often coordinated across a gene, yielding many non-independent features that increase multiple testing burden. Our results demonstrate that isoform-centric trait mapping with isoTWAS increases discovery by ∼40% compared with a matched local splicing event-based analysis, although these methods may recover some independent signal. Future work should consider integrating both reference-guided and annotation-free approaches for quantification and detection of isoform and splicing patterns to develop more nuanced mechanistic hypotheses for GWAS loci.

We conclude with some limitations of and future considerations for isoTWAS. First, we note that isoform-level expression quantifications are maximum-likelihood estimates, due to the inherent limitations of short-read RNA-seq. These estimates are generally guided by existing transcriptome annotations (e.g., GENCODE) and thus are dependent on the completeness and accuracy of these genomic annotations. Further, many dataset-specific sequencing factors may will affect the accuracy of these estimates, especially sequencing depth, read length and library preparation (mRNA vs total RNA sequencing). The continued emergence of long-read sequencing platforms, including PacBio IsoSeq and Oxford Nanopore Technologies, will be instrumental for improving reference transcriptome annotations, particularly with tissue specificity, which will in turn improve isoTWAS. Further, as these methods continue to gain scalability and cost effectiveness, they will ultimately replace short-read sequencing (and isoform estimation) for population-scale datasets. As isoTWAS is agnostic to the method of isoform expression quantification, this framework will continue to be applicable as we approach this long read sequencing era. Recent analyses have shown that larger sample sizes may outweigh sequencing coverage for eQTL mapping, but this relationship has not been investigated for isoform-level expression^95^. Thus, for optimal discovery with isoTWAS, the appropriate balance between sample size and reference panel sequencing depth remains to be determined.

Second, while inferential replicates from RNA-seq quantification can provide measures of technical variation, they are not incorporated into the predictive models. Our analyses of prediction across inferential replicates suggest a methodological opportunity: leveraging these inferential replicates as a measure of quantification error may help in estimating the robustness of isoform prediction and, potentially, of the precision of these SNP effects. A more flexible predictive model that estimates standard errors for SNP effects by model-averaging across the replicate datasets may help with trait mapping by providing a prediction interval for both isoform- and gene-level imputed expression. Third, just as TWAS can be cast as a differential gene expression analysis conducted with imputed expression, isoform-level trait mapping is akin to differential transcript expression analysis. An analogous goal of isoTWAS can be to detect trait associations with genetically-regulated transcript usage. However, it is unclear if the compositional nature of transcript usage data needs to accounted for at the prediction step or the trait mapping steps^96^. Lastly, as we show, this framework can suffer from reduced power, inflated false positives, and reduced sensitivity in fine-mapping in the presence of SNP horizontal pleiotropy, where the genetic variants in the isoform expression model affect the trait, independent of isoform expression, or when multiple SNPs affect expression of isoforms^97, 98^. For pathways that are not observed or accounted for in the reference expression panel and GWAS, accounting for horizontal pleiotropy may improve trait mapping. We motivate further methodological extensions of probabilistic fine-mapping to reconcile pleiotropy for SNPs shared across models for multiple isoforms at the same genetic locus, as summary-statistic based methods that control for horizontal pleiotropy are not yet effective^99^.

In sum, isoTWAS provides a novel framework to scalably interrogate the transcriptomic mechanisms underlying genetic associations with complex traits and generate biologically-meaningful and testable hypotheses about disease risk mechanisms. Based on our results, we emphasize a shift in focus from quantifications of the transcriptome on the gene-level to the transcript-isoform level to maximize discovery of transcriptome-centric genetic associations with complex traits.

## METHODS

isoTWAS consists of three steps: (1) training predictive models of isoform expression, (2) imputing isoform-specific expression into a separate GWAS panel, and (3) association testing between imputed expression and a phenotype (**Figure 1b**). isoTWAS contrasts with TWAS as it models correlations between the expression of isoforms of the same gene. We outline each step below, with further mathematical details in **Supplemental Methods**.

### Training predictive models of isoform expression

#### Model and assumptions

Assume a given gene *G* has *M* isoforms with expression levels across *N* samples, with each sample having *R* inferential replicates. Let 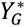 be the *N* × *M* matrix of mean isoform expression (log-scale TPM) for the *N* samples and *M* isoforms, using the point estimates from the Expectation-Maximization algorithm of a pseudo-mapping quantification algorithm, like Salmon or kallisto^32, 33^. We can jointly model isoform expression with a system of *N* × *M* × *R* equations. For sample *n* ∈ {1, …, *N*}, isoform *m* ∈ {1, …, *M*} of gene *G*, and replicate *r* ∈ {1, …, *R*}, we have:

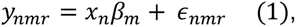

where *y*_*nmr*_ is the expression of isoform *m* for the *r*th inferential replicate of sample *n*, *x*_*n*_ is the *P*-vector (vector of length *P*) of *cis*-genotypes in a 1 Mb window around gene *G*, *β*_*m*_ is the *P*-vector of genetic effects of the *P* genotypes on isoform expression, and ϵ_*nmr*_ is normally distributed random noise with mean 0 and variance 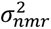. We standardize both the genotypes and the isoform expression to mean 0 and variance 1. As the SNP vector *x*_*n*_ does not differ across inferential replicates, we impose the following assumptions on the variance-covariance matrix of ϵ = (ϵ_1,1,1_, ϵ_1,1,2_, …, ϵ*_nmr_*)^*T*^, the vector of random errors: we assume that ϵ_*nmr*_ are independent and identically distributed across samples *n* ∈ {1, …, *N*} and identically distributed across replicates *r* ∈ {1, …, *R*}. Accordingly, the point estimates of the SNP effects on isoform expression are not influenced by differences in expression across replications. Therefore, in matrix form, we consider the following predictive model:

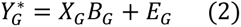

Here, *X*_*G*_ is the *N* × *P* matrix of genotype dosages, *B*_*G*_ is the *P* × *M* matrix of SNP effects on isoform expression and *E*_*G*_ is a matrix of random errors, such that *vec*(*E*_*G*_) ∼ *N*_*NM*_(0, *ɛ* = Ω^−1^ ⊗ *I*_*N*_). *ɛ* represents the variance-covariance matrix in the errors (with precision matrix *Ω* = Σ^−1^), following the aforementioned independence assumptions.

#### Estimating SNP effects on isoform expression

We apply 5 different methods to estimate 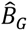, the matrix of SNP effects on isoform expression. The first four are multivariate methods that model the isoforms jointly; the last method models each isoform separately using univariate methods. The goal of this SNP effect estimation is marginal prediction, i.e., leveraging the correlation between isoforms to improve prediction of each isoform separately. The 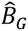 matrix that gives the largest adjusted R^2^ in 5-fold cross-validation across the 5 methods is selected as the final model to predict isoform expression for a given gene. In settings where we are interested in predicting gene-level expression from these predicted isoforms, isoTWAS trains an elastic net penalized linear regression that predicts gene-level expression from genetically-predicted isoform-level expression; this model training is conducted across the same 5 folds to prevent data leakage^100^. We provide an overview of the methods, with mathematical details in **Supplemental Methods**:

1. *Multivariate elastic net (MVEnet) regression*: This is an extension of elastic net, where the response is a matrix of correlated responses^43^. Here, the absolute penalty is imposed on each single coefficient by a group-lasso penalty on each vector of SNP effects across isoforms (rows of *B*_*G*_); accordingly, a SNP can only have a non-zero effect on an isoform if it has a non-zero effect on all isoforms.
2. *Multivariate LASSO Regression with covariance estimation (MRCE)*: We adapt Rothman et al’s proposed procedure to simultaneously and iteratively estimate both 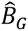, the SNP effects matrix, and 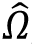, the precision matrix^44^. This procedure accounts for the correlation between isoforms but does not impose the group-lasso penalty as in MVEnet. As such, a single SNP need not have a non-zero effect on all isoforms.
3. *Multivariate elastic net with stacked generalization (joinet)*: We use Rauschenberger and Glaab’s joinet method that uses a two-step prediction^45^. In the first step, the design matrix of SNPs is used to generate a cross-validated prediction of each isoform. In the second step, the matrix of predicted isoform expression is used to predict each isoform.
4. *Sparse partial least squares (SPLS)*: This is an implementation of partial least squares with a sparsity penalty, that attempts to find an optimal latent decomposition for the linear relationship between the matrix of isoform expression and the design matrix of SNPs. We use the Chun and Keles’s implementation from the spls R package^46^.
5. *Univariate FUSION*: the simplest implemented method is the univariate predictive modelling used in FUSION^2^. We disregard the correlation structure between isoforms and train a univariate elastic net^43^, estimation of the best linear unbiased predictor (BLUP) in a linear mixed model^101^, and SuSiE^47^ predictive model for each isoform separately. The model with the largest adjusted R^2^ out of these three models is outputted. This approach serves as a baseline measurement for prediction of each isoform independently.

### Trait association and step-wise hypothesis testing

We note that the tests of association in isoTWAS are similar to tests in differential transcript expression analyses, as TWAS tests of association are analogous to tests in differential gene expression analyses. isoTWAS and TWAS are distinct, as these methods consider imputed isoform and gene expression, respectively, as predicted by the trained expression models. If individual-level genotypes are available in the external GWAS panel, isoform expression can be directly imputed by multiplying the SNP weights from the predictive model with the genotype dosages in the GWAS panel. If only summary statistics are available, we adopt the weighted burden test from Gusev et al with an ancestry-matched linkage disequilibrium panel^4, 98^. Compared to TWAS, a main distinction for isoTWAS association testing is the increased number of tests (approximately 4-fold the number of isoforms than genes)^21^ and the potential correlation in test statistics for isoforms of the same gene.

Accordingly, we perform a two-step hypothesis testing framework (**Supplemental Figure S2**)^102^. In the first step, for every isoform with a trained model, we generate a TWAS test statistic using either linear regression for GWAS with individual-level genotypes or the weighted burden test for GWAS with only summary statistics^4^. Given the *t* test statistics *T*_1_, …, *T*_*t*_ for a single gene, we used an omnibus test to aggregate the *t* test statistics into a single P-value for a gene. We benchmark different omnibus tests in simulations, but the default omnibus test in isoTWAS is aggregated Cauchy association test (ACAT)^49^. We control for false discovery across all genes via the Benjamini-Hochberg procedure, but the Bonferroni procedure can also be applied for more conservative false discovery control. In the second step, for isoforms for genes with an adjusted omnibus P < 0.05, we employ Shaffer’s modified sequentially rejective Bonferroni (MSRB) procedure to control the within-gene family-wide error rate. At the end of these two steps, we identify a set of genes and their isoforms that are associated with the trait.

### Control for false positives within GWAS loci

In TWAS and related methods, association statistics have been shown to be well calibrated under the null of no GWAS association. However, within loci harboring significant GWAS signal, false positive associations can result when eQTLs and GWAS coincide within overlapping LD blocks. To address this, we have adopted two conservative approaches to control for type 1 error within GWAS loci, namely (1) permutation testing and (2) probabilistic fine mapping. The permutation testing approach, adopted from Gusev et al^4^, is a highly conservative test of the signal added by the SNP-transcript effects from the predictive models, conditional on the GWAS architecture of the locus. Briefly, we shuffle the SNP-transcript effects in the predictive models 10,000 times and generate a null distribution for the TWAS test statistic for each isoform. We then use this null distribution to generate a permutation-based P-value for the original test statistic for each isoform. Additionally, we aggregate these null distributions using an omnibus test and compare the omnibus P-value to this distribution to generate permutation P-value for each gene. Finally, we can employ isoform-level probabilistic fine-mapping using methods from FOCUS^55^ to generate credible set of isoforms that explain the trait association at a locus. We only run isoform-level fine-mapping for significantly-associated isoforms in overlapping 1 Megabase windows.

### Simulation framework

We adopt techniques from Mancuso et al’s *twas_sim* protocol^103^ to simulate multivariate isoform expression based on randomly simulated genotypes and environmental random noise. First, for *n* samples, we generate a matrix of genotype dosages for the SNPs within 1 Megabase of 22 different genes (1 per chromosome) using an LD reference panel of European subjects from 1000 Genomes Project^51^.

Next, we generate a matrix of SNP-isoform effects across different causal SNP proportions *p*_*c*_, numbers of isoforms *t*, and *p*_*s*_ proportion of the SNP-isoform effects being shared across isoforms of the same gene. We then add two matrices of random noise *U* and ϵ. The first matrix *U* noise represents non cis-genetic effects on isoforms that are correlated between samples and isoforms; we control the proportion of variance explained in isoform expression attributed to *U* using a parameter ᓂ_*h*_. The second matrix ϵ is a matrix of random noise that is independent for each isoform, such that 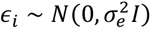 where 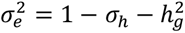. We generate 10,000 simulations for each configuration of the simulation parameters, varying 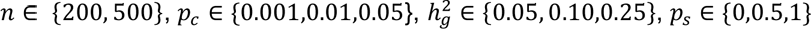, and 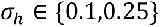. Full mathematical details are provided in **Supplemental Methods** and summarized in **Figure 2**.

We also generate traits under three distinct scenarios, with a continuous trait with heritability 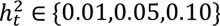 and a GWAS sample size of 50,000 (**Supplemental Methods**):

1. **Only gene-level expression has a non-zero effect on trait.** Here, we sum the isoform expression to generate a simulated gene expression. We randomly simulate the effect size and scale the error to ensure trait heritability.
2. **Only 1 isoform has a non-zero effect on the trait.** Here, we generate a multivariate isoform expression matrix with 2 isoforms and scale the total gene expression value such that one isoform (called the effect isoform) makes up *p*_*g*_ ∈ {0.10, 0.30, 0.50, 0.70, 0.90} proportion of total gene expression. We then generate effect size for one of the isoforms and scale the error to ensure trait heritability.
3. **Two isoforms with different effects on traits**. Here, we generate a multivariate isoform expression matrix with 2 isoforms that make up equal portions of the total gene expression. We then generate an effect size of *α* for one isoform and *p*_*e*_*α* for the other isoform, such that *p*_*e*_ ∈ {−1, −0.5, −0.2,0.2,0.5,1}. We then scale the error to ensure trait heritability.

Lastly, to estimate the approximate false positive rate (FPR), we followed the same simulation framework to generate eQTL data and GWAS data. In the GWAS data, however, we set the effect of gene- and isoform-level imputed expression to 0 to generate a simulated trait under the null, where the gene and isoforms are not associated with the trait. We then estimate the FPR by calculating the proportion of gene-trait associations at P < 0.05 under this null across 20 sets of 1,000 simulated GWAS panels. We also assess isoform-level fine-mapping using FOCUS in a scenario with a gene with 5 or 10 isoforms and a single effect isoform. We compute the sensitivity of 90% credible sets of isoforms (proportion of credible sets that contain the effect isoform) and the number of isoforms in the 90% credible set.

### GTEx processing and model training

We quantified GTEx v8^37^ RNA-Seq samples for 48 tissues using Salmon v1.5.2^32^ in mapping-based mode. We first built a Salmon index for a decoy-aware transcriptome consisting of GENCODE v38 transcript sequences and the full GRCh38 reference genome as decoy sequences^21^. Salmon was then run on FASTQ files with mapping validation and corrections for sequencing and GC bias. We computed 50 inferential bootstraps for isoform expression using Salmon’s Expectation-Maximization algorithm. We then imported Salmon isoform-level quantifications and aggregated to the gene-level using tximeta^41^.

Using edgeR, gene and isoform-level quantifications underwent TMM-normalization, followed by transformation into a log-space using the variance-stabilizing transformation using DESeq2^104, 105^. We then residualized isoform-level and gene-level expression (as log-transformed CPM) by all tissue-specific covariates (clinical, demographic, genotype PCs, and expression PEER factors) used in the original QTL analyses in GTEx. We calculated the quantification variance across inferential replicates using the computeInfRV() function from the fishpond package^106^. We computed the isoform fraction using the isoformToIsoformFraction() function from the IsoformSwitchAnalyzeR package^54^.

SNP genotype calls were derived from Whole Genome Sequencing data for samples from individuals of European ancestry, filtering out SNPs with minor allele frequency (MAF) less than 5% or that deviated from Hardy-Weinberg equilibrium at P < 10^-5^. We further filtered out SNPs with MAF less than 1% frequency among the European ancestry samples in 1000 Genomes Project^51^.

Details of the model training pipeline for GTEx are similar to those summarized in **Supplemental Figure S10**. Gene-level univariate models were trained using elastic net regression^43^, best linear unbiased predictor (BLUP) in a linear mixed model^48^, and SuSiE^47^, using all SNPs within 1 Mb of the gene body^4, 43, 47, 101^. For each gene, the best performing model was chosen based on McNemar’s adjusted 5-fold cross-validation (CV) R^2^. We selected only genes with CV R^2^ ≥ 0.01. We applied multivariate modeling outlined in isoTWAS to train isoform-level predictive models, selecting only those isoform models with CV R^2^ ≥ 0.01 (**Supplemental Figure S2**). All isoTWAS models generated are publicly available (see **Data Availability**).

### Developmental brain reference panel processing and model training

We quantified developmental frontal cortex^24^ (N = 205) RNA-Seq samples using Salmon v1.8.0^32^ in mapping-based mode. We used the same indexed transcriptome as in the GTEx analysis and ran Salmon with mapping validation and corrections for sequencing and GC bias. We computed 50 inferential bootstraps for isoform expression using Salmon’s Expectation-Maximization algorithm. We then imported Salmon isoform-level quantifications and aggregated to the gene-level using tximeta^41^. Using edgeR, gene and isoform-level quantifications underwent TMM-normalization, followed by transformation into a log-space using the variance-stabilizing transformation using DESeq2^104, 105^. We then residualized isoform-level and gene-level expression (as log-transformed CPM) by covariates (age, sex, 10 genotype PCs, 90 and 70 hidden covariates with prior, respectively). Typed SNPs with non-zero alternative alleles, MAF > 1%, genotyping rate > 95%, HWE P < 10^-6^ were first imputed to TOPMed Freeze 5 using minimac4 and eagle v2.4^107, 108^. We then retained biallelic SNPs with imputation accuracy R^2^ > 0.8, with rsIDs. Finally, we filtered out SNPs with MAF < 0.05 or that deviated from Hardy-Weinberg equilibrium at P < 10^-6^.

### Adult brain reference panel processing and model training

Matched genotype and RNAseq data from adult brain cortex tissue from n = 2,365 individuals were compiled and processed from the PsychENCODE Consortium^22^ and the Accelerating Medicines Partnership Program for Alzheimer’s Disease (AMP-AD)^56^, consisting of the individual studies BipSeq, BrainGVEX, CommonMind Consortium (CMC), CommonMind Consortium’s National Institute of Mental Health Human Brain Collection Core (CMC HBCC), Lieber Institute for Brain Development-szControl (LIBD_szControl), UCLA-ASD, Religious Orders Study and the Memory and Aging Project (ROSMAP), Mount Sinai Brain Bank (MSBB) and MayoRNAseq.

Typed genotypes were lifted over to the GRCh38 build using CrossMap v.0.6.3^109^ and then filtered to remove variants where the reference allele matched any of the alternate alleles. Genotype data from whole genome sequencing (BrainGVEX, UCLA-ASD, ROSMAP, MSBB, and MayoRNAseq) was further filtered to variants present on the Infinium Omni5-4 v1.2 array in order to satisfy the imputation server’s maximum limit of 20,000 typed variants per 20Mb. All genotype data was further processed with PLINK v1.90b6.21^110^, removing variants with HWE P < 10^-6^, MAF< 0.01 or missingness rate > 0.05, and removing samples with missingness rate > 0.1 across typed variants or missingness rate > 0.5 on any individual chromosome. Genotype data was prepared for imputation using the McCarthy Group’s HRC-1000G-check-bim-v4.3.0 tool against freeze 8 of the Trans-Omics for Precision Medicine (TOPMed) reference panel^111^. The tool removes A/T and G/C SNPs with MAF > 0.4, variants with alleles that differ from the reference panel, variants with an allele frequency difference > 0.2 from the reference panel and variants not in the reference panel. Additionally, the tool updates strand, position and reference/alternate allele assignment to match the reference panel.

Genotypes were then passed into the TOPMed Imputation Server by individual array batch^112^. The genotypes were phased with Eagle v2.4 and imputed with Minimac4 using the TOPMed reference panel^107, 108^. Further QC was performed on the imputed genotypes using bcftools v1.11 and PLINK. The imputed genotypes were filtered to well-imputed variants with R2 > 0.8. The arrays were merged after filtering to variants that were well-imputed in all arrays to be merged. Only arrays with at least 400,000 variants after pre-imputation QC were merged in order to prevent too many variants from dropping out. The merged genotype data was then converted to PLINK 1 binary format and further processed with PLINK, removing variants with duplicates, HWE P < 10^-6^, MAF < 0.01 or missingness rate > 0.05 and removing samples with missingness rate > 0.1. Samples from the same individual were identified by calculating the genetic relatedness matrix using SnpArrays.jl and finding sets of samples with relatedness > 0.75. From each set of replicates, only the genotyped sample from the array with the most variants after pre-imputation QC was kept. For model training, only SNPs annotated in HapMap3 were retained^113^.

RNAseq paired reads from each study were sorted by name and then converted to FASTQ format using samtools v1.14^114^. The reads were then quantified using salmon v1.8.0 in mapping-based mode using a full decoy indexed from GENCODE v38 transcriptome and GRCh38 patch 13 assembly^32^. Quantification was run using standard EM algorithm with library type automatically inferred and estimates adjusted for sequence-specific and fragment-level GC biases. Bootstrapped abundance estimates were calculated using 50 bootstrap samples. Isoform-level expression was summarized to the gene-level using tximeta^115^. Only isoforms with 0.1 TPM for more than 75% of samples were retained. The resulting expression was normalized using the variance-stabilizing transformation from DESeq2^105^. Samples with WGCNA network connectivity scores of less than -3 were removed as outliers, resulting in a total of 2,115 samples^116^. Isoform- and gene-level expression was then batch-corrected using ComBat, using study site as the batch^117^. Lastly, age, sex, squared age, 10 genotype PCs, and 200 (for gene expression) and 175 (for isoform expression) hidden covariates with prior (using sequencing metrics from PicardTools to estimate the prior) were removed from the expression^118, 119^. The number of hidden covariates with prior (HCP) were selected by optimizing the number of nominal cis-gene-level expression QTLs and cis-isoform-level expression QTLs at Bonferroni-corrected P < 0.05, respectively, on a grid from 100 to 300 HCPs, as detected by QTLtools^120^.

Details of the model training pipeline are summarized are equivalent to those used to train models in GTEx data. All isoTWAS models generated are available at https://zenodo.org/record/679594792.

### Gene- and isoform-level trait mapping

We conducted gene- and isoform-level trait mapping for 15 neuropsychiatric traits: attention-deficit hyperactivity disorder (ADHD, N_cases_ = 20,183/N_controls_ = 35,191)^59^, Alzheimer’s disease (ALZ, 90,338/1,036,225)^60^, anorexia nervosa (AN, 16,992/55,525)^72^, autism spectrum disorder (ASD, 18,381/27,969)^58^, bipolar disorder (BP, 41,917/371,549)^61^, brain volume (BV, N = 47,316)^62^, cross-disorder (CDG, 232,964/494,162)^63^, cortical thickness (CortTH, N = 51,665)^64^, intracranial volume (ICV, N = 32,438)^65^, major depressive disorder (MDD, 246,363/561,190)^66^, neuroticism (NTSM, N = 449,484)^67^, obsessive compulsive disorder (OCD, 2,688/7,037)^68^, panic and anxiety disorders (PANIC, 2,248/7,992)^69^, post-traumatic stress disorder (PTSD, 32,428/174,227)^70^, and schizophrenia (SCZ, 69,369/236,642)^71^. For gene-level trait mapping, we used the weighted burden test, followed by the permutation test, as outlined by Gusev et al^4^. For isoform-level trait mapping, we used the stage-wise testing procedure outlined in the isoTWAS method. In-sample LD from the QTL reference panels was used to calculate the standard error in the weighted burden test. For isoforms, irrespective of their corresponding genes, passing both stage-wise tests and the permutation test, we employed isoform-level probabilistic fine-mapping using FOCUS with default parameters^55^. These methods are summarized in **Supplemental Figure S24**.

We estimated the percent increase in effective sample size by employing the following heuristic. We convert gene-level association P-values into χ² test statistics with 1 degree-of-freedom. For χ² > 1, we then calculate the percent increase for isoTWAS-based associations versus TWAS-based associations. As the mean of the χ² distribution is linearly related to power and sample size^121^, we can use this percent increase in test statistic as a measure of power or effective sample size. We defined independent genome-wide significant SNPs in GWAS by LD clumping with lead GWAS SNP < 5 × 10^-8^ with P-value used for ranking and a R^2^ threshold of 0.2.

## Data Availability

GTEx genetic, transcriptomic, and covariate data were obtained through dbGAP approval at accession number phs000424.v8.p2. Linkage disequilibrium reference data from the 1000 Genomes Project were obtained at this link: https://www.internationalgenome.org/data-portal/sample. GWAS summary statistics were obtained at the following links: ADHD (https://www.med.unc.edu/pgc/download-results/), ALZ (https://ctg.cncr.nl/software/summary_statistics/), AN (http://www.med.unc.edu/pgc/results-and-downloads), ASD (https://www.med.unc.edu/pgc/download-results/), BP (https://www.med.unc.edu/pgc/download-results/), BV (https://ctg.cncr.nl/software/summary_statistics), CDG (https://www.med.unc.edu/pgc/results-and-downloads), CortTH (https://enigma.ini.usc.edu/research/download-enigma-gwas-results/), ICV (https://enigma.ini.usc.edu/research/download-enigma-gwas-results/), MDD (http://dx.doi.org/10.7488/ds/2458), NTSM (https://ctg.cncr.nl/software/summary_statistics/neuroticism_summary_statistics), OCD (https://www.med.unc.edu/pgc/download-results/), PANIC (https://www.med.unc.edu/pgc/download-results/), PTSD (https://www.med.unc.edu/pgc/results-and-downloads/), and SCZ (https://www.med.unc.edu/pgc/download-results/). The Developmental Brain RNA-seq and genotype dataset from Walker et al is available at dbGAP with accession number phs001900. The subset of Adult Brain RNA-seq and genotype data from the PsychENCODE Consortium is available at https://psychencode.synapse.org/DataAccess and from AMP-AD is available at https://adknowledgeportal.synapse.org/Data%20Access. GWAS summary statistics and accession numbers to genotype and RNA-seq data are provided in Supplemental Table S10. isoTWAS models for 48 tissues from GTEx are available at https://zenodo.org/record/8047940122, adult brain cortex from PsychENCODE and AMP-AD are available at https://zenodo.org/record/8048198123, and developmental brain cortex from Walker et al are available at https://zenodo.org/record/8048137124.

https://zenodo.org/record/8048198

https://zenodo.org/record/8047940

https://zenodo.org/record/8048137

https://www.internationalgenome.org/data-portal/sample

https://www.med.unc.edu/pgc/download-results/

https://ctg.cncr.nl/software/summary_statistics/

https://enigma.ini.usc.edu/research/download-enigma-gwas-results/

http://dx.doi.org/10.7488/ds/2458

https://psychencode.synapse.org/DataAccess

https://adknowledgeportal.synapse.org/Data%20Access

## DATA AVAILABILITY

GTEx genetic, transcriptomic, and covariate data were obtained through dbGAP approval at accession number phs000424.v8.p2. Linkage disequilibrium reference data from the 1000 Genomes Project were obtained at this link: https://www.internationalgenome.org/data-portal/sample. GWAS summary statistics were obtained at the following links: ADHD (https://www.med.unc.edu/pgc/download-results/), ALZ (https://ctg.cncr.nl/software/summary_statistics/), AN (http://www.med.unc.edu/pgc/results-and-downloads), ASD (https://www.med.unc.edu/pgc/download-results/), BP (https://www.med.unc.edu/pgc/download-results/), BV (https://ctg.cncr.nl/software/summary_statistics), CDG (https://www.med.unc.edu/pgc/results-and-downloads), CortTH (https://enigma.ini.usc.edu/research/download-enigma-gwas-results/), ICV (https://enigma.ini.usc.edu/research/download-enigma-gwas-results/), MDD (http://dx.doi.org/10.7488/ds/2458), NTSM (https://ctg.cncr.nl/software/summary_statistics/neuroticism_summary_statistics), OCD (https://www.med.unc.edu/pgc/download-results/), PANIC (https://www.med.unc.edu/pgc/download-results/), PTSD (https://www.med.unc.edu/pgc/results-and-downloads/), and SCZ (https://www.med.unc.edu/pgc/download-results/). The Developmental Brain RNA-seq and genotype dataset from Walker et al is available at dbGAP with accession number phs001900. The subset of Adult Brain RNA-seq and genotype data from the PsychENCODE Consortium is available at https://psychencode.synapse.org/DataAccess and from AMP-AD is available at https://adknowledgeportal.synapse.org/Data%20Access. GWAS summary statistics and accession numbers to genotype and RNA-seq data are provided in **Supplemental Table S10**. isoTWAS models for 48 tissues from GTEx are available at https://zenodo.org/record/8047940122, adult brain cortex from PsychENCODE and AMP-AD are available at https://zenodo.org/record/8048198123, and developmental brain cortex from Walker et al are available at https://zenodo.org/record/8048137124.

## CODE AVAILABILITY

isoTWAS is available as an R package at https://github.com/bhattacharya-a-bt/isotwas. Sample scripts for analyses are available at https://github.com/bhattacharya-a-bt/isotwas_manu_scripts.

## SUPPLEMENTAL INFORMATION

### Supplemental Methods

Supplemental Methods are provided in the attached SupplementalMethods.pdf.

### Supplemental Figure Legends

**Figure S1:** *Directed acyclic graph (DAG) illustrating causal assumptions in isoTWAS.* We assume that the local genetic variants within 1 Megabase of a gene have direct effects on the expression of a gene G and its isoforms; these genetic effects need not be shared across isoforms and the gene. Furthermore, we assume that the abundance of a gene is the sum of abundances of its isoforms. Lastly, we assume that the isoform and gene need not affect the complex trait through the same path. We also acknowledge that genetic variants may have effects on the trait through pathways independent of gene and isoform expression.

**Figure S2**: *Step-wise hypothesis testing in isoTWAS.* First, isoform-trait associations are estimated either using appropriate regressions of imputed expression in individual-level GWAS or the weighted burden test in Gusev et al in GWAS summary statistics. Then, associations for isoforms of the same gene are aggregated to the gene-level using the Aggregated Cauchy Association Test (ACAT). These aggregated gene-level associations are adjusted for multiple testing burden to control the false discovery rate (FDR) using either a Bonferroni or Benjamini-Hochberg procedure. Lastly, for isoforms of genes that pass gene-level testing, we control the family-wide error rate (FWER) using Shaffer’s modified sequentially rejective procedure.

**Figure S3**: *Comparison of predictive performance of 6 models implemented in isoTWAS.* Across 1,000 simulations of 5 isoforms with isoform heritability (*h*^2^) set to 0.05 or 0.10. Boxplots of adjusted R^2^ of prediction of isoform expression (Y-axis) across shared isoQTL proportion (X-axis)

**Figure S4**: *IsoTWAS models predict gene expression with more accuracy than TWAS models*. Boxplots of percent difference in adjusted R^2^ in predicting gene expression between isoTWAS and TWAS models from simulations with sample size 200 (compare with sample size 500 in **Figure 2**), where isoform and gene expression heritability are set to (top) 0.05 and (bottom) 0.10.

**Figure S5**: Across 48 GTEx tissues (Y-axis), the number of multivariate (cream) and univariate (blue) models predicting isoform expression at CV R^2^ > 0.01 (X-axis).

**Figure S6**: Across 48 GTEx tissues (Y-axis), percent difference in CV R^2^ (X-axis) of prediction of isoform expression models using multivariate models versus univariate models. The label shows the proportion of isoforms with improved performance using multivariate models.

**Figure S7**: Number of isoforms with CV R^2^ > 0.01 (Y-axis) using the baseline univariate model (teal, best univariate) and 4 multivariate models.

**Figure S8**: Across 48 GTEx tissues (Y-axis), number of genes that pass TWAS (blue) and isoTWAS (red) CV R^2^ cutoffs to be available for testing in the trait-mapping step (X-axis)

**Figure S9**: Across 48 GTEx tissues (Y-axis), percent difference in CV R^2^ (X-axis) of prediction of isoform expression models using multivariate models versus univariate models. The label shows the proportion of isoforms with improved performance using multivariate models.

**Figure S10**: Across 48 GTEx tissues (Y-axis), number of genes predicted at CV R^2^ > 0.01 using TWAS (blue) and isoTWAS (red)

**Figure S11**: Number of genes (left) and isoforms (right) predicted at CV R^2^ > 0.01 using isoTWAS across 48 GTEx tissues.

**Figure S12**: Performance of isoTWAS across number of isoforms per gene across 48 GTEx tissues. **(a)** Ratio of number of isoforms predicted at R^2^ > 0.01 using multivariate versus univariate prediction. **(b)** Ratio of number of genes passing CV threshold using isoTWAS versus TWAS. **(c)** Median number of isoforms predicted at CV R^2^ > 0.01 in isoTWAS models across increasing number of isoforms per gene. The red line shows the line Y = × + 1. **(d)** Ratio of number of genes with CV R^2^ > 0.01 using isoTWAS versus TWAS.

**Figure S13**: Performance of isoTWAS across increasing maximum isoform fraction across 48 GTEx tissues. **(a)** Ratio of number of isoforms predicted at R^2^ > 0.01 using multivariate versus univariate prediction. **(b)** Ratio of number of genes passing CV threshold using isoTWAS versus TWAS. **(c)** Ratio of number of genes with CV R^2^ > 0.01 using isoTWAS versus TWAS.

**Figure S14**: Performance of isoTWAS across increasing gene length across 48 GTEx tissues. **(a)** Ratio of number of isoforms predicted at R^2^ > 0.01 using multivariate versus univariate prediction. **(b)** Ratio of number of genes passing CV threshold using isoTWAS versus TWAS. **(c)** Ratio of number of genes with CV R^2^ > 0.01 using isoTWAS versus TWAS. R^2^ > 0.01 using isoTWAS versus TWAS.

**Figure S15**: Performance of isoTWAS across increasing SNP density across 48 GTEx tissues. **(a)** Ratio of number of isoforms predicted at R^2^ > 0.01 using multivariate versus univariate prediction. **(b)** Ratio of number of genes passing CV threshold using isoTWAS versus TWAS. **(c)** Ratio of number of genes with CV R^2^ > 0.01 using isoTWAS versus TWAS.

**Figure S16**: Performance of isoTWAS across increasing sample size across 48 GTEx tissues. **(a)** Ratio of number of isoforms predicted at R^2^ > 0.01 using multivariate versus univariate prediction. **(b)** Ratio of number of genes passing CV threshold using isoTWAS versus TWAS. **(c)** Ratio of number of genes with CV R^2^ > 0.01 using isoTWAS versus TWAS.

**Figure S17**: Performance of isoTWAS across proportion of shared isoTWAS model effect SNPs across 48 GTEx tissues. **(a)** Ratio of number of isoforms predicted at R^2^ > 0.01 using multivariate versus univariate prediction. **(b)** Ratio of number of genes passing CV threshold using isoTWAS versus TWAS. **(c)** Ratio of number of genes with CV R^2^ > 0.01 using isoTWAS versus TWAS.

**Figure S18**: *Performance of isoTWAS across increasing mean counts of isoforms and genes across 48 GTEx tissues.* **(a)** Ratio of number of isoforms predicted at R^2^ > 0.01 using multivariate versus univariate prediction across increasing mean counts of isoforms. **(b)** Ratio of number of genes with CV R^2^ > 0.01 using isoTWAS versus TWAS across increasing mean counts of genes.

**Figure S19**: Performance of isoTWAS across increasing quantification variance of isoforms and genes across 48 GTEx tissues. **(a)** Ratio of number of isoforms predicted at R^2^ > 0.01 using multivariate versus univariate prediction across increasing quantification variance of isoforms. **(b)** Ratio of number of genes with CV R^2^ > 0.01 using isoTWAS versus TWAS across increasing quantification variance of genes.

**Figure S20**: **(a)** Median percent difference in R^2^ of predicting original isoform expression using multivariate versus univariate models across increasing number of isoforms per gene, colored by models trained in the original dataset (pink) and the leave-one-out dataset (teal) **(b)** Median percent difference in R^2^ of predicting original gene expression using isoTWAS versus TWAS models across increasing number of isoforms per gene, colored by models trained in the original dataset (pink) and the leave-one-out dataset (teal)

**Figure S21**: *IsoTWAS and gene-level TWAS show relatively similar false positive rates*. Across 20 iterations of 1,000 simulations, boxplots of false positive rate to detect a gene-trait association using Cauchy-aggregated P-values of isoform-trait associations (red) and gene-level TWAS (blue). In these simulations, we simulate 200 and 5,000 samples in the eQTL and GWAS panels, 5 isoforms, and isoform and gene expression heritability of 0.10. We vary the causal isoQTLs proportion (p_c_, shown on X-axis), the QTL architecture (right margin), and correlation between isoforms (top margin). For each iteration, we simulate one eQTL panel and 1,000 GWAS panels where genetically-regulated gene and isoform expression have no effect on the trait. We calculate the false positive rate as the proportion of the 1,000 tests that give P > 0.05.

**Figure S22**: Power comparison between TWAS and isoTWAS in detecting gene-trait association across 3 scenarios **(Methods)**. **(a)** Power to detect gene-trait association (proportion of tests with P < 2.5 × 10^-6^, Y-axis) across number of total isoforms per gene (X-axis), facetted by proportion of shared isoQTLs (top margin) and proportion of expression heritability attributed to shared non-genetic effects across isoforms (right margin). Points are shaped by causal isoQTL proportion and colored by method. **(b)** Power to detect gene-trait association (proportion of tests with P < 2.5 × 10^-6^, Y-axis) across proportion of gene expression explained by effect isoform (X-axis), facetted by proportion of shared isoQTLs (top margin) and proportion of expression heritability attributed to shared non-genetic effects across isoforms (right margin). Points are shaped by number of isoforms per gene and colored by method. **(c)** Power to detect gene-trait association (proportion of tests with P < 2.5 × 10^-6^, Y-axis) across ratio of effect sizes of 2 effect isoforms (X-axis), facetted by proportion of shared isoQTLs (top margin) and proportion of expression heritability attributed to shared non-genetic effects across isoforms (right margin). Points are shaped by number of isoforms per gene and colored by method. Here, expression heritability is set of 0.05, trait heritability is set to 0.1, and causal proportion of **(b-c)** is set of 0.01.

**Figure S23**: Sensitivity **(a)** and mean set size **(b)** of 90% credible set using FOCUS to finemap isoform-trait associations for a single gene, across causal isoQTL proportion (X-axis). Points are colored by trait heritability and shaped by the number of isoforms per gene. Plots are facetted by proportion of shared isoQTLs (top margin) and proportion of expression heritability attributed to shared non-genetic effects across isoforms (right margin). Line-range shows a 95% jackknife confidence interval.

**Figure S24:** *Schematic diagram for analysis using adult and developmental frontal cortex data from PsychENCODE and AMP-AD.* Data sources for eQTL reference data, GWAS cohorts, and reference LD data are provided on the left (black). The full gene-level TWAS (red) and isoTWAS (blue) are summarized on the right.

**Figure S25**: Number of gene-trait associations (Y-axis) using TWAS (red) and isoTWAS (blue) across trait (X-axis), faceting by tissue (top margin) and threshold (right margin: adjusted P < 0.05 and permutation P < 0.05, top; in 90% credible set using FOCUS fine-mapping, bottom).

**Figure S26**: Number of isoform-trait associations (Y-axis) using isoTWAS across trait (X-axis), faceting by tissue (top margin) and threshold (right margin: adjusted P < 0.05 and permutation P < 0.05, top; in 90% credible set using FOCUS fine-mapping, bottom).

**Figure S27**: QQ-plots of gene-level P-values using TWAS (red) and isoTWAS (blue) across 15 traits.

**Figure S28**: **(a)** Distribution of number of genes in risk region (left) and in 90% credible set (right) using TWAS and isoTWAS. **(b)** Distribution of number of isoforms per gene in risk region (left) and in 90% credible set (right) using isoTWAS.

**Figure S29**: Lollipop plot of enrichment ratio (X-axis) of ontologies (Y-axis) for isoTWAS-prioritized genes associated at adjusted P < 0.05 and permutation P < 0.05. Points are shaped by tissue type (adult or developmental) and colored by ontology type (biological process, cell component, molecular function).

**Figure S30**: For ENST00000681794 association with BV **(a)** and ENST00000492957 with BV **(b)**, Manhattan plots of GWAS, eQTL, and isoQTL signal colored by LD (top), boxplots of gene (red) and isoform (blue) expression (Y-axis) by genotype (X-axis) (bottom left), and forest plot of lead isoQTL association with isoform (blue), gene (red), and trait (black) (bottom right). **(c)** Comparison of exon and intron structure of ENST00000681794 and ENST00000492957, based on Gencode v38 reference.

### Supplemental Tables

**Table S1**: Sample size, source, and tissues for functional genomics reference panels

**Table S2**: Number of genes/models that pass cross-validation prediction cutoffs using TWAS and isoTWAS feature selection criteria and prediction methods

**Table S3**: Distribution of predictive external R2 of observed total gene expression vs. predicted total gene expression (isoTWAS -TWAS)

**Table S4**: Number of GWAS loci with an isoTWAS-, TWAS-, and splice-TWAS-prioritized gene within 0.5 Mb

**Table S5**: TWAS and fine-mapping results for genes with adjusted P < 0.05 and permutation P < 0.05 across 15 traits using adult brain cortex models

**Table S6**: isoTWAS and fine-mapping results for genes with adjusted P < 0.05 and permutation P < 0.05 across 15 traits using adult brain cortex models

**Table S7**: TWAS and fine-mapping results for genes with adjusted P < 0.05 and permutation P < 0.05 across 15 traits using developmental brain cortex models

**Table S8:** isoTWAS and fine-mapping results for genes with adjusted P < 0.05 and permutation P < 0.05 across 15 traits using developmental brain cortex models

**Table S9**: Empirical bayes estimates of test statistic inflation and increase in χ² test statistics of gene-level associations using isoTWAS and TWAS

**Table S10:** Accession numbers and URLs for data access

## Supplemental Data

**Data 1:** Predictive performance comparison of isoTWAS multivariate methods in simulated data across a variety of genetic architecture settings

**Data 2**: Predictive performance comparison of isoTWAS and TWAS gene expression prediction in simulated data across a variety of genetic architecture settings

**Data 3**: Isoform expression prediction metrics across a variety of factors, using 48 GTEx datasets

**Data 4**: Gene expression prediction metrics across a variety of factors, using 48 GTEx datasets

**Data 5**: False positive rates using isoTWAS and TWAS to detect a gene-trait association at P < 0.05 across a variety genetic architecture parameters

**Data 6**: Power to detect trait association at P < 2.5 × 10^-6^ across 1,000 simulations each for 22 genes using TWAS and isoTWAS across various simulations. These simulations are under Scenario 1 in Figure 4a (gene has a true effect on the trait, but none of the isoforms have a true effect on the trait)

**Data 7**: Power to detect trait association at P < 2.5 × 10^-6^ across 1,000 simulations each for 22 genes using TWAS and isoTWAS (ACAT) across various simulations. These simulations are under Scenario 2 in Figure 4b (a gene has multiple isoforms, only one has an effect on the trait, and we vary the usage of this effect isoform)

**Data 8**: Power to detect trait association at P < 2.5 × 10^-6^ across 1,000 simulations each for 22 genes using TWAS and isoTWAS (ACAT) across various simulations. These simulations are under Scenario 3 in Figure 4c (a gene has two isoforms with differing effects on the trait, and we vary the effect size of one of the isoforms)

**Data 9**: Sensitivity and mean set size of 90% credible sets determined by FOCUS in simulated data across a variety of genetic architecture parameters

**Data 10**: Raw TWAS results across 15 neuropsychiatric traits using adult brain cortex expression models

**Data 11**: Raw isoTWAS results across 15 neuropsychiatric traits using adult brain cortex expression models

**Data 12**: Raw TWAS results across 15 neuropsychiatric traits using developmental brain cortex expression models

**Data 13**: Raw isoTWAS results across 5 neuropsychiatric traits using developmental brain cortex expression models

**Data 14**: GWAS and nominal eQTL and isoQTL summary statistics corresponding to isoTWAS isoform-trait association examples shown in Figure 7 and Supplemental Figure S30.

## ACKNOWLEDGEMENTS

We thank Kangcheng Hou, Tommer Schwarz, Vidhya Venkateswaran, Pan Zhang, Leanna Hernandez, Nathan LaPierre, Harold Pimentel, Mike Love, and Achal Patel for engaging discussion during the research process. We thank Kanishka Patel for her aesthetic advice for figures. We thank the Psychiatric Genomics Consortium and Complex Trait Genomics Lab for their publicly available GWAS summary statistics.

BP was partially supported by NIH awards R01 HG009120, R01 MH115676, R01 CA251555, R01 AI153827, R01 HG006399, R01 CA244670, and U01 HG011715. MJG was supported by SFARI Bridge to Independence Award, NIMH R01-MH121521, NIMH R01-MH123922, and NICHD-P50-HD103557.

## AUTHOR INFORMATION

### Contributions

Conceptualization: AB, BP, MJG; Methodology: AB, BP, MJG; Software: AB; Validation: AB; Formal analysis: AB, MK, CW, CJ; Investigation: AB, MJG; Resources: BP, MJG; Data curation: AB, MK, CW, CJ, MJG; Writing – original draft: AB, MJG; Writing – reviewing and editing: AB, MK, CW, CJ, BP, MJG; Visualization: AB; Supervision: BP, MJG; Project administration: BP, MJG; Funding acquisition: BP, MJG

### Correspondence

Please direct all correspondence to Arjun Bhattacharya.

