## Supplemental Methods for "Isoform-level transcriptome-wide association uncovers extensive novel genetic risk mechanisms for neuropsychiatric disorders in the human brain"

Supplemental Materials

### Supplemental Methods

Here, we describe mathematical details of the predictive and hypothesis testing methods in the isoTWAS pipeline.

#### Predictive modeling

For a gene $G$ with $M$ isoforms across $N$ samples, with expression measured across $R$ inferential replicates, we consider the following multivariate linear model:

$$\mathbf{Y}_G^* = \mathbf{X}_G \mathbf{B}_G + \mathbf{E}_G, \tag{1}$$

where

- $\mathbf{Y}_G^*$ is the $N \times M$ matrix of isoform expression for gene $G$,
- $\mathbf{X}_G$ is the $N \times P$ matrix of genotype dosages (coded as 0,1, or 2 alternative alleles at a SNP) for SNPs within a *cis*-window of the body $G$,
- $\mathbf{B}_G$ is the $P \times M$ matrix of SNP effects on isoform expression, and
- $\mathbf{E}_G$ is a matrix of random errors, such that vec$(\mathbf{E}_G) \sim N_{NM}\left(0, \mathbf{\Sigma} = \mathbf{\Omega}^{-1} \otimes \mathbf{I}_N\right)$. Here, $\mathbf{\Sigma}$ is the variance-covariance matrix of the random errors, with $\mathbf{\Omega} = \mathbf{\Sigma}$ representing the precision matrix. The columns of $\mathbf{X}_G$ can be standardized to mean 0 and variance 1 to remove the intercept term from the model.

We implement 4 different multivariate methods to estimate $\hat{B}_G$.

##### Multivariate elastic net

Multivariate elastic net is an extension of elastic net regression for a multivariate response variable. The optimization here, fit through coordinate descent, solves

$$\text{argmin}_{\mathbf{B}_G} \left\{ \frac{1}{2N} \sum_{i=1}^{N} \|y_i - \mathbf{B}_G^T x_{G,i}\|_F^2 + \lambda \left[ (1-\alpha)\|\mathbf{B}_G\|_F^2/2 + \alpha \sum_{j=1}^{P} \|\beta_{G,j}\|_2 \right] \right\}.$$

Here, $\beta_{G,j}$ is the $j$th row of the SNP effects matrix $\mathbf{B}_G$. There is a group-lasso penalty on each $M$-length vector of isoform effects for a single SNP. This penalty works on the whole group of coefficients for each response: either all coefficients are 0, or none are 0. All coefficients are shrunk by the $\lambda$ penalty, optimally selected through cross-validation. Intuitively, multivariate elastic net should be optimal in settings where the causal isoQTLs are the shared across different isoforms of the same gene. We fit this model using the `glmnet` package in `R`[1] for the mixing parameter $\alpha \in \{0, .5, 1\}$.

#### Multivariate Regression with LASSO with Covariance Estimation

From Equation 1, we jointly estimate $\mathbf{B}_G$ and $\mathbf{\Omega}$ by minimizing the following objective function:

$$\left(\hat{\mathbf{B}}_G, \hat{\mathbf{\Omega}}\right) = \text{argmin}_{\mathbf{B}_G,\mathbf{\Omega}} \left\{ g(\mathbf{B}_G, \mathbf{\Omega}) + \lambda_1 \sum_{j' \neq j} |\omega_{j',j}| + \lambda_2 \sum_{j=1}^{p} \sum_{k=1}^{q} |b_{jk}| \right\},$$

where

$$g(\mathbf{B}_G, \mathbf{\Omega}) = \text{tr}\left[n^-1(\mathbf{Y}_G^* - \mathbf{X}_G\mathbf{B}_G)^T(\mathbf{Y}_G^* - \mathbf{X}_G\mathbf{B}_G)\mathbf{\Omega}\right] - \log|\mathbf{\Omega}|.$$

This objective function can be iteratively minimized for both matrix parameters. In any given iteration, we first solve of $\hat{\mathbf{B}}_G$ with a fixed $\mathbf{\Omega}$ using coordinate descent. Then, we can solve for $\hat{\mathbf{\Omega}}$ with the fixed $\hat{\mathbf{B}}_G$ at the given iteration with graphical lasso. We iterate until the convergence tolerance parameter is met. Full details are outlined in Rothman et al[2].

#### Multivariate elastic net regression using stacked generalization

We employ Rauschenberger and Glaab's `joinet` R package[3] that uses a stacked generalization for multivariate elastic net regression. In general, `joinet` has two steps for prediction:

1. In the first step, or layer, each column of $\mathbf{Y}_G^*$ is predicted from $\mathbf{X}_G$ using elastic net regression via cross-validation to prevent data leakage. This gives us a predicted vector $Y_{g,m}^{(cv)}$ of isoform expression for each isoform $m$, and taken together, a predicted matrix of isoform expressions $\mathbf{Y}_G^{(cv)}$.
2. In the second layer, each column of $\mathbf{Y}_G^*$ is predicted from $\mathbf{Y}_G^{(cv)}$ with LASSO regression.

Through this two-step prediction, we can estimate a matrix of predicted SNP-isoform effects $\hat{\mathbf{B}}_G$. For each SNP, this stacking process exchanges information among the estimated effects on the isoforms, such that the final estimated effect one a single isoform combines the initial SNP effect estimates on all isoforms linearly.

#### Sparse partial least squares

Lastly, we use sparse partial least squares[4], as implemented in the `spls` R package. First, it is important to note that partial least squares is a alternative to ordinary least squares for linear regression models without proper conditions. Partial least squares hinges on a dimension reduction technique that assumes that there is a latent decomposition of the response matrix (matrix of isoform expression in the case of isoTWAS) and the predictor matrix (the design matrix of SNPs). This latent decomposition is represented with a $K$-dimensional matrix $\mathbf{T}$. Partial least squares estimates $\mathbf{T} = \mathbf{X}_G\mathbf{W}$ through successive optimization steps to find the columns of $\mathbf{W}$ using an objective function that depends on the columns of $\mathbf{W}$ and the covariance between the response and predictor matrices.

Sparse partial least squares identifies this latent decomposition with added parameters to induce sparsity. In short, let $\mathbf{M} = \mathbf{X}'_G\mathbf{Y}^*_G\mathbf{Y}'^*_G\mathbf{X}_G$. Sparse partial least squares attempts to minimize the following objective function for $\omega$ and $c$, subject to $\omega'\omega = 1$:

$$-\kappa\omega'\mathbf{M}\omega + (1-\kappa)(c-\omega)'\mathbf{M}(c-\omega) + \lambda_1\|c\|_1 + \lambda_2\|c\|_2^2.$$

There are four parameters that require tuning through cross-validation $(\kappa, \lambda_1, \lambda_2, K)$. In isoTWAS, we find the optimal $\kappa \in \{0.1, 0.2, 0.3, \dots, 0.9\}$, $K \in \{1, 2, \dots, \lfloor M/2 \rfloor\}$, $\lambda_1$ and $\lambda_2$ through 5-fold cross-validation.

The isoTWAS package also allows the user to fit a univariate model for each isoform. We use this as a baseline to compare the advantages of multivariate modelling to this univariate approach.

#### Univariate modeling

The simplest method implemented is univariate predictive modelling, as implemented in Gusev et al's FUSION software[5]. We ignore the correlation structure between isoforms and train a univariate model. For the $m$th isoform, we fit:

$$y^*_{G,m} = \mathbf{X}_G\beta_{G,m} + \epsilon_{G,m} \tag{2}$$

We include three univariate methods:

1. **Elastic net regression with elastic net mixing parameter** $\alpha = 0.5$[1]. This procedure finds the $\hat{\beta}_{G,m}$ that minimizes

   $$L(\beta_{G,m}) = \frac{1}{2N}\sum_{i=1}^{N}(y_{G,m,i} - x^T_{G,i}\beta_{G,m})^2 + \lambda[(1-\alpha)\|\beta_{G,m}\|_2^2/2 + \alpha\|\beta_{G,m}\|_1].$$

   We use the `glmnet` package in `R` for implementation with cross-validation.

2. **Best linear unbiased predictor (BLUP) using a linear mixed model**[6]. Here, we assume, in Equation 2, that $\beta_{G,m}$ are random SNP effects on the isoform $m$, such that $\beta_{G,m} \sim \mathbf{N}\left(\mathbf{0}, \frac{\sigma^2_m}{P}I_N\right)$. Here, $\sigma^2_m$ is a variance parameter for the SNP effects. We can calculate the BLUP of $\beta_{G,m}$ with the following solution of the Henderson mixed-model[6]:

   $$\hat{\beta}_{G,m} = \frac{\hat{\sigma}^2_m}{M}\mathbf{X}^T_G\hat{\mathbf{V}}^{-1}y^*_{G,m},$$

   where $\hat{\sigma}^2_m$ and $\mathbf{V} = \sigma^2_m\mathbf{X}_G\mathbf{X}^T_G/P + \sigma^2_\epsilon\mathbf{I}_N$ are estimated with restricted maximum likelihood estimation and subsequent matrix multiplication. We implement an estimation to this model using ridge regression with the `rrBLUP`package in `R`.

3. **Sum of Single Effects (SuSiE) regression.** Here, we assume that, in Equation 2, $\beta_{G,m} = \sum_{i=1}^{L}\beta_{l,G,m}$, where $\beta_{l,G,m}$ has exactly one non-zero element. SuSiE estimates the variance components using maximum likelihood prior to the estimating $\beta_{G,m}$ using an empirical Bayes approach. We implement this procedure using the `susieR` package in `R`[7].

### Association testing procedure

We employ a stage-wise testing procedure, similar to the `stageR` method[8].

1. We impute genetically-regulated expression of each isoform and estimate associations between each isoform using (1) the appropriate linear regression if we have access to individual-level genotypes in the GWAS and (2) the weighted burden test if we only have access to GWAS summary statistics[5]. We use an LD reference panel that appropriately matches the ancestry of the GWAS sample and the eQTL sample the predictive models were trained with.
2. Given the Wald-type test statistics $Z_1, \ldots, Z_m$ for a given gene, we run an omnibus test to aggregate the test statistics of isoforms of the same gene. We employ either (1) minimum P-value aggregation (i.e. set the gene-level omnibus P-value to the minimum isoform-level P-value), (2) an aggregated Cauchy association test (ACAT)[9], or (3) Chi-square aggregation, where we define the gene-level test statistic $T_G = \sum_{i=1}^{m} Z_i^2$ and compare to the Chi-square distribution with $m$ degrees of freedom. We correct for multiple comparisons using the Benjamini-Hochberg procedure.
3. We then run an isoform-level multiple testing procedure using the Shaffer MSRB method to assess all isoform-level associations[8]. This procedure controls the family-wide error rate when hypotheses are correlated within the family (i.e. isoforms of the same gene).

Given any overlapping isoforms (i.e. isoforms within 0.5 Megabases of one another), we use transcript-level probabilistic fine-mapping[10] to generate a 90% credible set of associated isoforms.

### Simulation framework and parameters

Here, we adopt techniques from Mancuso et al's `twas_sim` package[11] to simulate multivariate isoform expression. We consider the following model

$$\mathbf{Y} = \mathbf{X}\mathbf{B} + \mathbf{U} + \epsilon,$$

where, for $n$ total samples, $\mathbf{Y}$ is an $n \times m$ matrix of expression values for $m$ isoforms, $\mathbf{X}$ is an $n \times p$ matrix of $p$ SNPs within 1 Megabase of the isoforms in $\mathbf{Y}$, $\mathbf{B}$ is an $p \times m$ matrix of SNP-isoform effects, $\mathbf{U}$ is the non-cis genetic effects on isoforms that are correlated between both isoforms and samples, and $\epsilon$ represents the independent noise added to each isoform separately. We first simulate the SNPs in $\mathbf{X}$ by selecting all the SNPs within 1 Megabase of 22 randomly selected genes (1 per chromosome), by using the linkage disequilibrium matrix from European samples of the 1000 Genomes Project and the framework outlined in `twas_sim`. We then simulate $\mathbf{B}$ by selecting $p_c$ proportion of the SNPs in $\mathbf{X}$ as "causal" and generating a non-zero effect size for these SNPs. We allow for a proportion, $p_s$, of these "causal" SNPs to be shared across different isoforms. For example, if we set $p_s = 0.50$, we select $0.5p_c$ of the SNPs to be shared across all isoforms and assign, for each isoform, a non-zero effect for these selected shared SNPs. For each isoform, an additional $0.5p_c$ proportion of the SNPs will be randomly selected as non-zero effect SNPs. We then scale each column of $\mathbf{B}$ to ensure that the genetically-determined portion of each column of $\mathbf{Y}$ equals the isoform expression heritability parameter $h_g^2$.

Next, we simulate $U \sim MVN(\mathbf{0}, \sigma_h V, \sigma_h W)$. $\sigma_h$ is a tunable parameter for controlling the proportion of variance in isoform expression explained, and $V$ and $W$ are correlation matrices between isoforms and samples, respectively. As simulating positive-semidefinite matrices, especially of large dimension, is difficult, we employ a heuristic that roughly generates dense correlation matrices (off-diagonals are far from 0) for $V$ and sparser correlation matrices (off-diagonals are closer to 0) for $W$. For $V$ and $W$, we first generate $V_1$ , an $m \times m$ matrix, and $W_1$, an $n \times n$ matrix, where each off-diagonal element is drawn from Unif$(-.5, .5)$ or Unif$(-0.02, 0.02)$, respectively, and the diagonal is set to 1. We then set $V = V_1' V_1 / max(V_1)$ and $W = W_1' W_1 / max(W_1)$. Lastly, we draw $\epsilon_i \sim N(\mathbf{0}, \sigma_e^2 I)$, where $\sigma_e^2 = 1 - \sigma_h - h_g^2$.

We conduct these simulations 10,000 times across the following set of parameters:

- $n \in \{200, 500, 1000\}$
- $p_c \in \{0.001, 0.01, 0.05\}$
- $h_g^2 \in \{0.05, 0.10, 0.25\}$
- $p_s \in \{0, 0.5, 1\}$
- $\sigma_h \in \{0.1, 0.25\}$

For the GWAS dataset, we first generate genotypes and genetically-regulated isoform expression using the same framework as the QTL dataset and the same causal $\mathbf{B}$ matrix. We then estimate traits in 3 scenarios with a GWAS sample size of 50,000:

1. **Only gene-level expression has a non-zero effect on trait.** Here, we sum the isoform expression to generate a simulated gene expression. We randomly simulate the effect size and scale the error to ensure trait heritability $h_t^2 \in \{0.01, 0.05, 0.10\}$.
2. **Only 1 isoform has a non-zero effect on the trait.** Here, we generate a multivariate isoform expression matrix with 2 isoforms and scale the total gene expression value such that one isoform (called the effect isoform) makes up $p_g \in \{0.10, 0.30, 0.50, 0.70, 0.90\}$ proportion of total gene expression. We then generate effect size for one of the isoforms and scale the error to ensure trait heritability $h_t^2 \in \{0.01, 0.05, 0.10\}$.
3. **Two isoforms with different effects on traits**. Here, we generate a multivariate isoform expression matrix with 2 isoforms that make up equal portions of the total gene expression. We then generate an effect size of $\alpha$ for one isoform and $p_e\alpha$ for the other isoform, such that $p_e \in \{-1, -0.5, -0.2, 0.2, 0.5, 1\}$. We then scale the error to ensure trait heritability $h_t^2 \in \{0.01, 0.05, 0.10\}$.

We also benchmark transcript-level fine-mapping using FOCUS[10]. Here, we use a similar framework, as above. We simulate a gene with 5 or 10 isoforms with the same QTL architecture parameters. We randomly selected one of the isoforms to be the "causal" effect isoform on the trait in Scenario 2 above. Then, we run transcript-level fine-mapping using FOCUS and record the size of the 90% credible set of isoforms and the sensitivity of the 90% credible set (i.e., the proportion of credible sets that contain the "causal" isoform).
