## Supplemental Note for "Isoform-level transcriptome-wide association uncovers extensive novel genetic risk mechanisms for neuropsychiatric disorders in the human brain"

Here, we discuss added evaluations of isoTWAS and TWAS: (1) across different factors that may influence prediction and (2) when dominant isoforms are failed to be detected.

**Gene and isoform expression prediction across different factors**

We evaluated the prediction of multivariate isoform-centric prediction models across a variety of factors that may influence the genetic architecture of isoform regulation at a locus or the inference of the regulation. In general, we computed three ratios: (1) the isoform prediction ratio, or the ratio of number of isoforms that are predicted at CV R^2^ > 0.01 using multivariate and univariate models, (2) the inclusion criterion ratio, or the ratio of the number of genes that meet inclusion criteria for isoTWAS (gene with 1+ isoforms predicted at CV R^2^ > 0.01) compared to TWAS (gene is predicted at CV R^2^ > 0.01), and (3) the gene prediction ratio, or the ratio of the number of genes that are predicted at CV R^2^ > 0.01 using isoform-centric isoTWAS models compared to gene-centric TWAS models. We plot boxplots of these ratios across the 48 GTEx tissues, and vary these factors across bins. In general, we note that, despite trends in these ratios across these factors, the ratios are always above 1, reinforcing the gains the prediction afforded by the multivariate isoform-centric prediction in isoTWAS.

1. *Number of expressed isoforms per gene*. **Supplemental Figure S12a** shows that the isoform prediction ratio stays relatively even as the number of isoforms per gene increases. The inclusion criterion ratio increases as the number of isoforms per gene increases but only until approximately 10 isoforms per gene. For genes with >10 isoforms per gene, the inclusion criterion ratio remains relatively even (**Supplemental Figure S12b**). There is a clear increasing trend in the median number of well-predicted isoforms per gene (CV R^2^ > 0.01) as the number of expression isoforms per gene increases, suggesting that the increased number of isoforms per gene provides more information about shared genetic architecture between isoforms that can be leveraged for improved prediction (**Supplemental Figure S12c**). Lastly, in **Supplemental Figure S12d** we see a similar increasing trend in the gene prediction ratio as the number of isoforms per gene increases (increase in the ratio until approximately 10 isoforms per gene and a leveling off after).
2. *Maximum isoform fraction per gene.* We computed, using the raw counts of isoforms, the isoform fraction of each isoform of a gene using the isoformtoIsoformFraction() function in the Bioconductor package IsoformSwitchAnalyzeR^1^. This function computes the fraction of each isoform’s expression to the total gene expression. We then found the isoform with the maximum isoform fraction for each gene and termed this fraction the maximum isoform fraction for the gene. In general, genes with large maximum isoform fraction are dominated by a single isoform, whereas genes with a small maximum isoform fraction have multiple isoforms with similar levels of expression. **Supplemental Figure S13** shows that, as maximum isoform fraction increases, there is a slight increase in the isoform prediction ratio, a larger increase in the inclusion criterion ratio, and no general trend in the gene prediction ratio.
3. *Gene length*. We computed the length of each gene as the difference in the end and start positions of the gene, as annotated in Ensembl v109. **Supplemental Figure S14** shows that, as gene length increases, there is a slight increase in both the isoform prediction ratio and the inclusion criterion ratio, and no general trend in the gene prediction ratio.
4. *SNP density.* We computed the number of SNPs that are within 1 Mb of the gene body, calling this value the SNP density of the gene locus. The SNP density represents the number of SNPs that comprise the design matrix in both the isoform-centric isoTWAS and gene-centric TWAS prediction models. **Supplemental Figure S15** shows that, as SNP density increases, there is a slight decrease in the isoform prediction ratio but no general trend in the inclusion criterion ratio and gene prediction ratio.
5. *Sample size.* **Supplemental Figure S16** shows that, as sample size increases, there is a decrease in the gene prediction ratio. This may reflect that, with larger sample sizes, gene-level expression QTLs may start to reflect the more subtle isoform-level expression QTLs, leading to a decrease in this ratio. We do note that, even at the largest sample sizes in GTEx, this gene prediction ratio is greater than 1. In datasets of small sample size (<175 samples), the isoform prediction ration and inclusion criterion ratio are largest. These ratios decrease in datasets of larger sample size, but the ratio does not decrease consistently (e.g., isoform prediction ratio is higher in datasets of 250-500 samples compared to 175-200 and inclusion criterion ratio remain relatively similar as sample size increases beyond 175).
6. *Proportion of shared isoTWAS model effect SNPs.* For each isoform’s predictive model, we determined which SNPs have large effects. First, we standardized the effect sizes in the model to mean 0 and unit variance. We found the SNPs whose effect sizes deviated significantly (Benjamini-Hochberg adjusted P < 0.05) and called them the isoform’s effect SNPs. For each gene, we then computed the proportion of effect SNPs that were shared across all isoforms of the same gene. **Supplemental Figure S17** shows a clear increasing trend in the isoform prediction ratios, indicating that multivariate modelling can leverage shared isoform QTL architecture to improve marginal prediction of each isoform’s expression. The inclusion criterion ratio remains relatively even as this proportion increases. The expression prediction ratio shows a decreasing trend as the proportion of shared isoTWAS effect SNPs increases, reflecting results from simulation (**Figure 2**).
7. *Mean normalized counts.* Here, using the isoform and gene counts normalized to library size and gene length, we compute each isoform and gene’s mean normalized count across samples, using the countsFromAbundance = ‘lengthScaledTPM’ option from tximport. Since the bins of mean normalized counts for gene and isoform expression do not map one-to-one as for the previous factors, we only compute and plot the isoform prediction and gene prediction ratios. As the mean normalized counts for isoform expression increases, we see a decreasing trend in the isoform prediction ratio, with a slight increase in the bin of isoforms with large mean normalized counts. We see no clear trend in the gene expression prediction as the mean normalized gene counts increase, though we see a similar increase in the largest bin (**Supplemental Figure S18**).
8. *Quantification variance across inferential replicates of genes and isoforms.* Here, using the raw isoform and gene counts, we compute the quantification variance across inferential replicates from Salmon^2^. We obtain the 50 inferential replicated from Salmon and import this using the Bioconductor package tximport^3^. We then computed the quantification variance for each isoform and gene using the computeInfRV() function from the Bioconductor package fishpond^4^. Briefly, this function first computes a matrix of variance (samples by features) across the 50 inferential replicates. Then, it computes the inferential quantification variance as the difference of the variance matrix and the mean counts matrix (Salmon Expectation-Maximization point estimates), standardized by the mean counts matrix. We collapse this inferential quantification variance to a per-feature measure by taking the mean of each row in the inferential quantification variance matrix. Again, since the bins of quantification variance for gene and isoform expression do not map one-to-one as for the previous factors, we only compute and plot the isoform prediction and gene prediction ratios. We find, for isoforms with low quantification variance (variance < 1.5), the isoform prediction ratio stays relatively even but increases as isoform quantification variance exceeds 1.5. However, there is no general trend with gene prediction ratio as gene quantification variance increases (**Supplemental Figure S19**). Leveraging this quantification variance to improve prediction is an interesting and worthwhile methodological opportunity that is discussed in the Discussion section.

**Synthetic leave-one-isoform-out models**

We consider a corollary experiment to assess how well isoTWAS prediction models impute gene and isoform expression when isoforms are failed to be detected. Across the 13 brain tissues in GTEx, we selected a random set of ~9000 genes in the following manner:

- We stratified our gene sets by the number of isoforms per gene and subset to genes with between 3 to 16 isoforms.
- We then randomly selected 50 genes from each group of genes with a certain number of isoforms per gene.

Then, we generated a synthetic leave-one-isoform-out (LOO) dataset, where the dominant isoform of each gene is missing. Using the raw transcript-level salmon quantifications, we removed each gene’s dominant isoform and summarized gene expression using tximeta’s summarizeToGene() function. Then, we trained isoTWAS and TWAS models in the synthetic datasets and imputed isoform (using isoTWAS) and gene (using isoTWAS and TWAS) expression in the original GTEx datasets. We compared these leave-one-out predictions to predictions from models trained in the original GTEx datasets.

**Supplemental Figure S20a** plots the distribution of the percent difference in isoform expression (using multivariate compared to univariate models), when using the original and synthetic leave-one-out training datasets. We find that the advantage of the multivariate models over univariate models greatly decreases with using this leave-one-out dataset compared to the true original, true gene expression measures. In addition, this drop in performance is consistent as we increase the number of isoforms per gene in the original dataset. This result is unsurprising as, in the leave-one-out dataset, though each isoform’s expression remains equal to its expression in the original dataset, the correlation structure is altered. The multivariate models depend on shared genetic architecture to best predict each isoform’s expression marginally. **Supplemental Figure 20b** plots the distribution of the percent difference in gene expression (using isoTWAS compared to TWAS models), when using the original and synthetic leave-one-out training datasets. We find that the advantage of the isoTWAS models over TWAS models greatly decreases with using this leave-one-out dataset compared to the true original, but only for genes with smaller numbers of isoforms per gene. As the number of isoforms per gene in the original dataset increases, this drop in performance decreases. In general, these results suggest that, when a large portion of the total gene’s expression is removed from the dataset, both isoform- and gene-centric expression prediction is negatively affected. Taken together, these results about isoforms underscores that proper quantification of isoforms is important, especially when building a genetic predictor, and motivates a need to develop more accurate and updated transcriptome annotations that are tissue-specific to aid isoform expression quantification. We discuss this further in the Discussion section.
