## Supplemental Figures S1-S30 for "Isoform-level transcriptome-wide association uncovers extensive novel genetic risk mechanisms for neuropsychiatric disorders in the human brain"

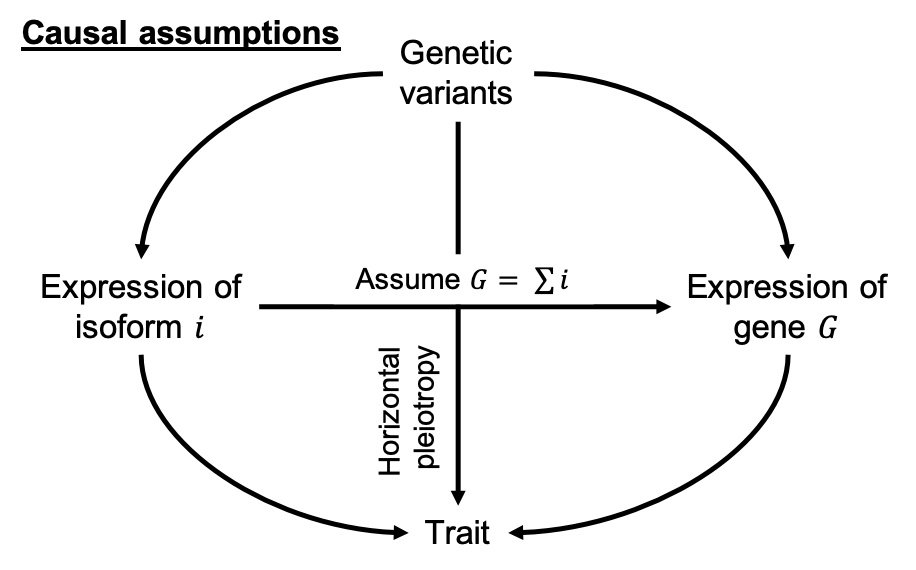

### **Figure S1**: *Directed acyclic graph (DAG) illustrating causal assumptions in isoTWAS.* We assume that the local genetic variants within 1 Megabase of a gene have direct effects on the expression of a gene $G$ and its isoforms; these genetic effects need not be shared across isoforms and the gene. Furthermore, we assume that the abundance of a gene is the sum of abundances of its isoforms. Lastly, we assume that the isoform and gene need not affect the complex trait through the same path. We also acknowledge that genetic variants may have effects on the trait through pathways independent of gene and isoform expression.

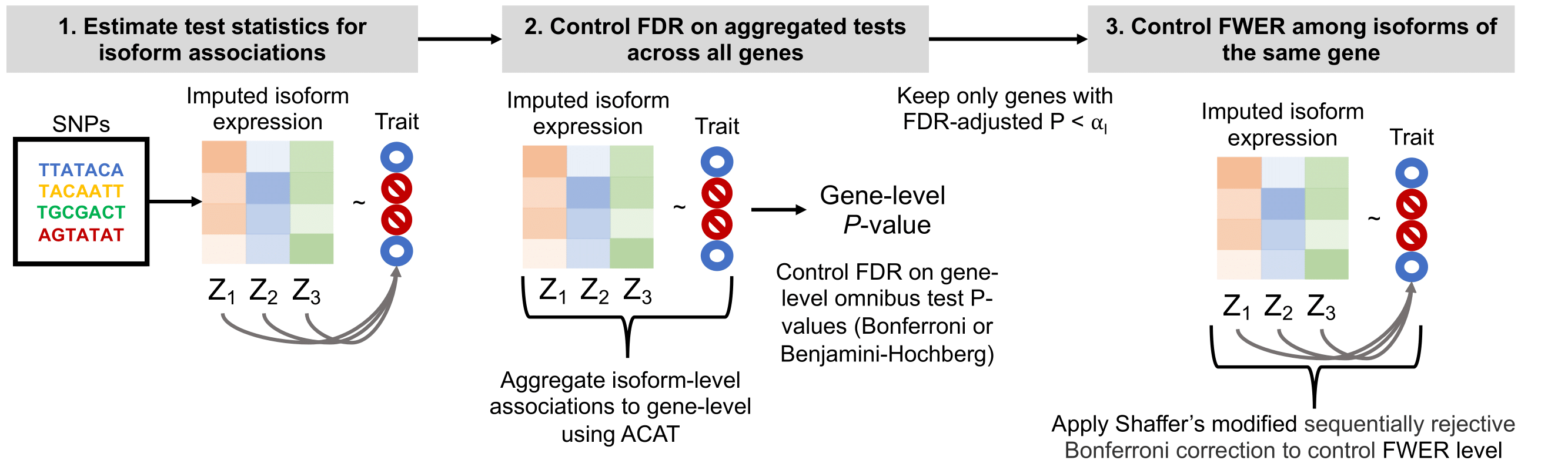

### **Figure S2**: *Step-wise hypothesis testing in isoTWAS.* First, isoform-trait associations are estimated either using linear regressions of imputed expression in individual-level GWAS or the weighted burden test using GWAS summary statistics. Then, associations for isoforms of the same gene are aggregated to the gene-level using the Aggregated Cauchy Association Test (ACAT). These aggregated gene-level associations are adjusted for multiple testing burden to control the false discovery rate (FDR) using either a Bonferroni or Benjamini-Hochberg procedure. Lastly, for isoforms of genes that pass gene-level testing, we control the family-wide error rate (FWER) using Shaffer’s modified sequentially rejective procedure.

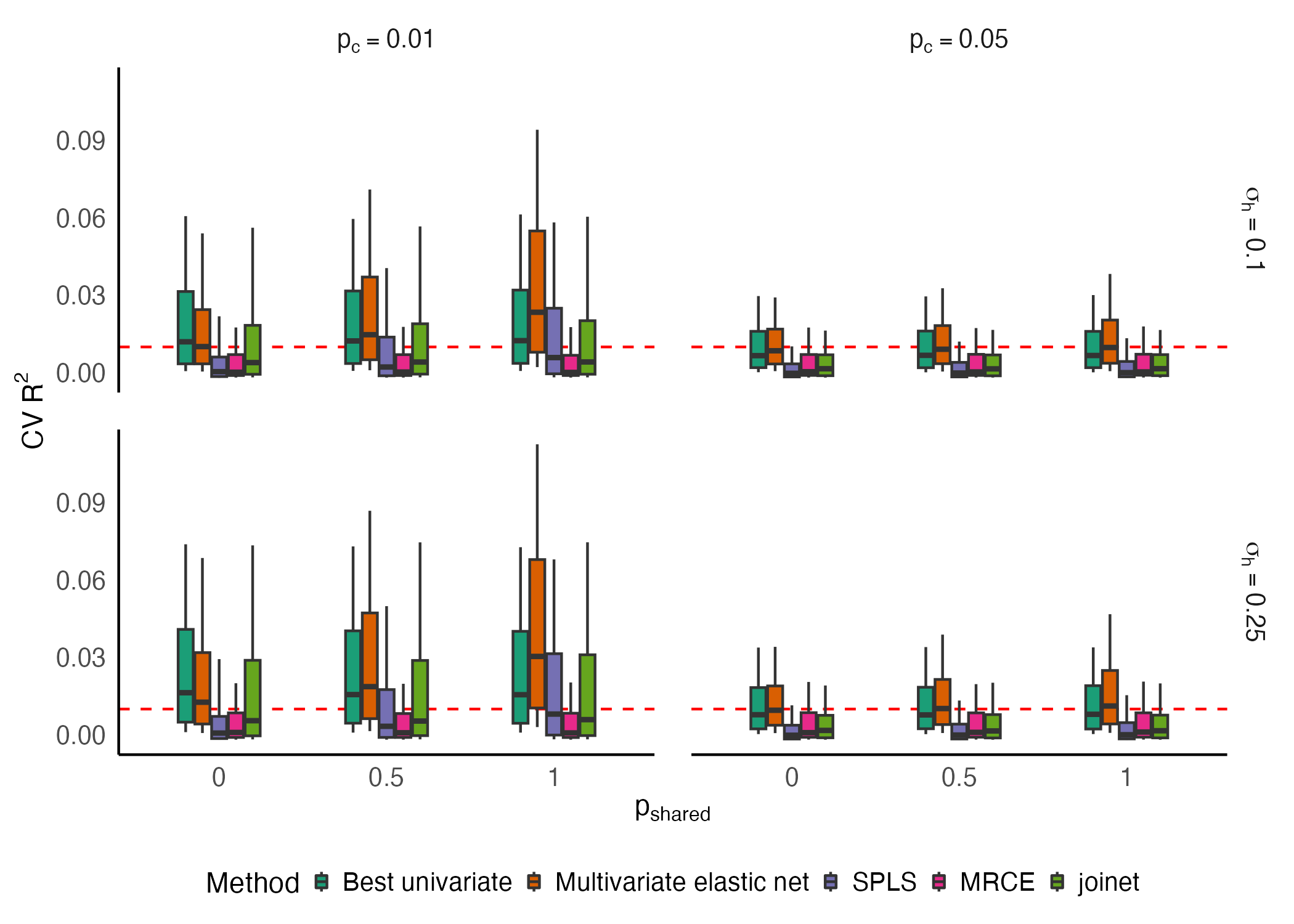

**Figure S3**: *Predictive performance across 5 models implemented in isoTWAS.* Across 1,000 simulations of 5 isoforms with isoform heritability ($h_{i}^{2}$) set to 0.05 or 0.10. Boxplots of adjusted R^2^ of prediction of isoform expression (Y-axis) across shared isoQTL proportion (X-axis).

**Figure S4**: *Improved prediction of total gene expression with isoTWAS versus TWAS models*. Boxplots of percent difference in adjusted R^2^ in predicting gene expression between isoTWAS and TWAS models from simulations with sample size 200 (compare with sample size 500 in **Figure 2**), where isoform and gene expression heritability are set to (top) 0.05 and (bottom) 0.10.

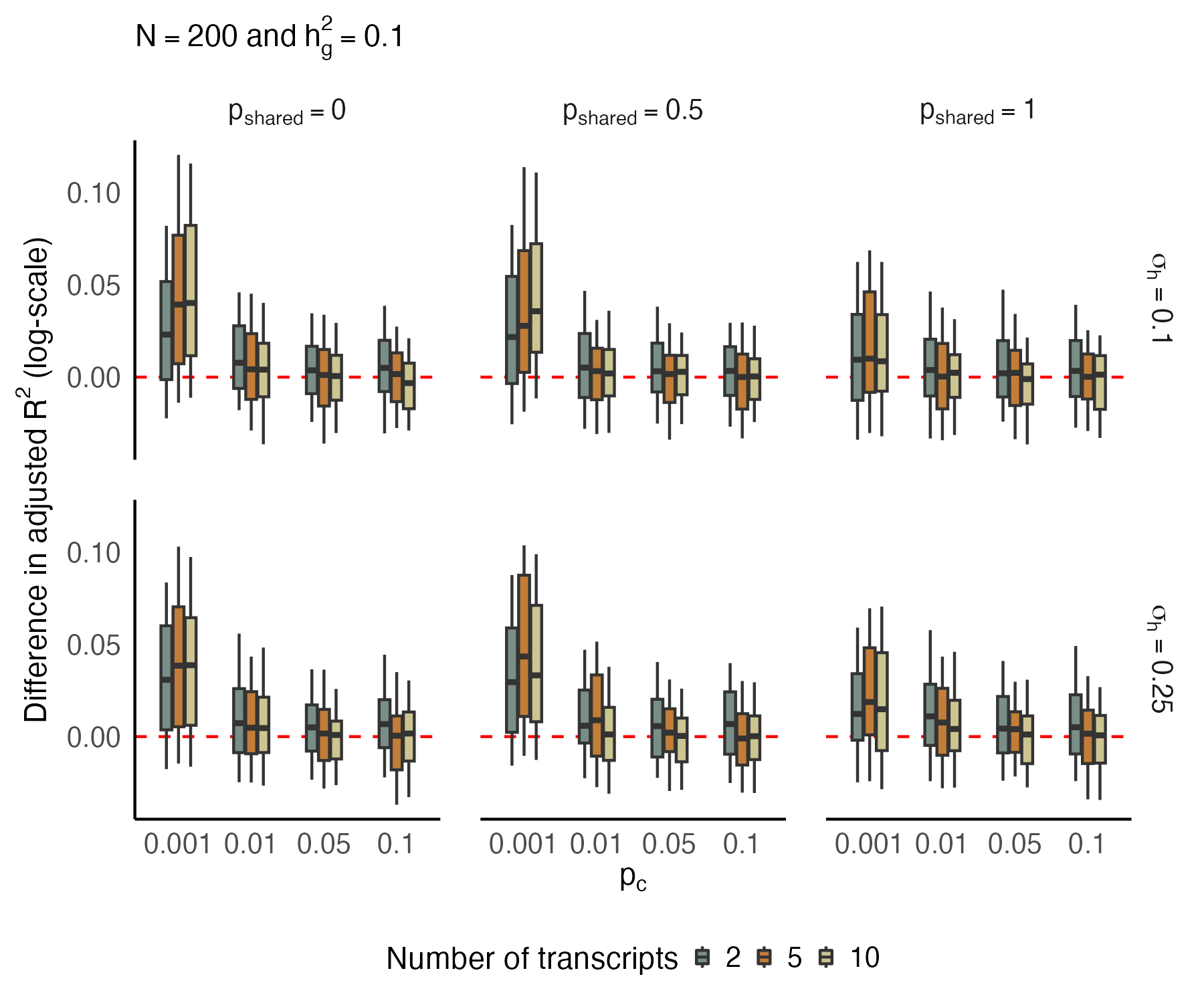

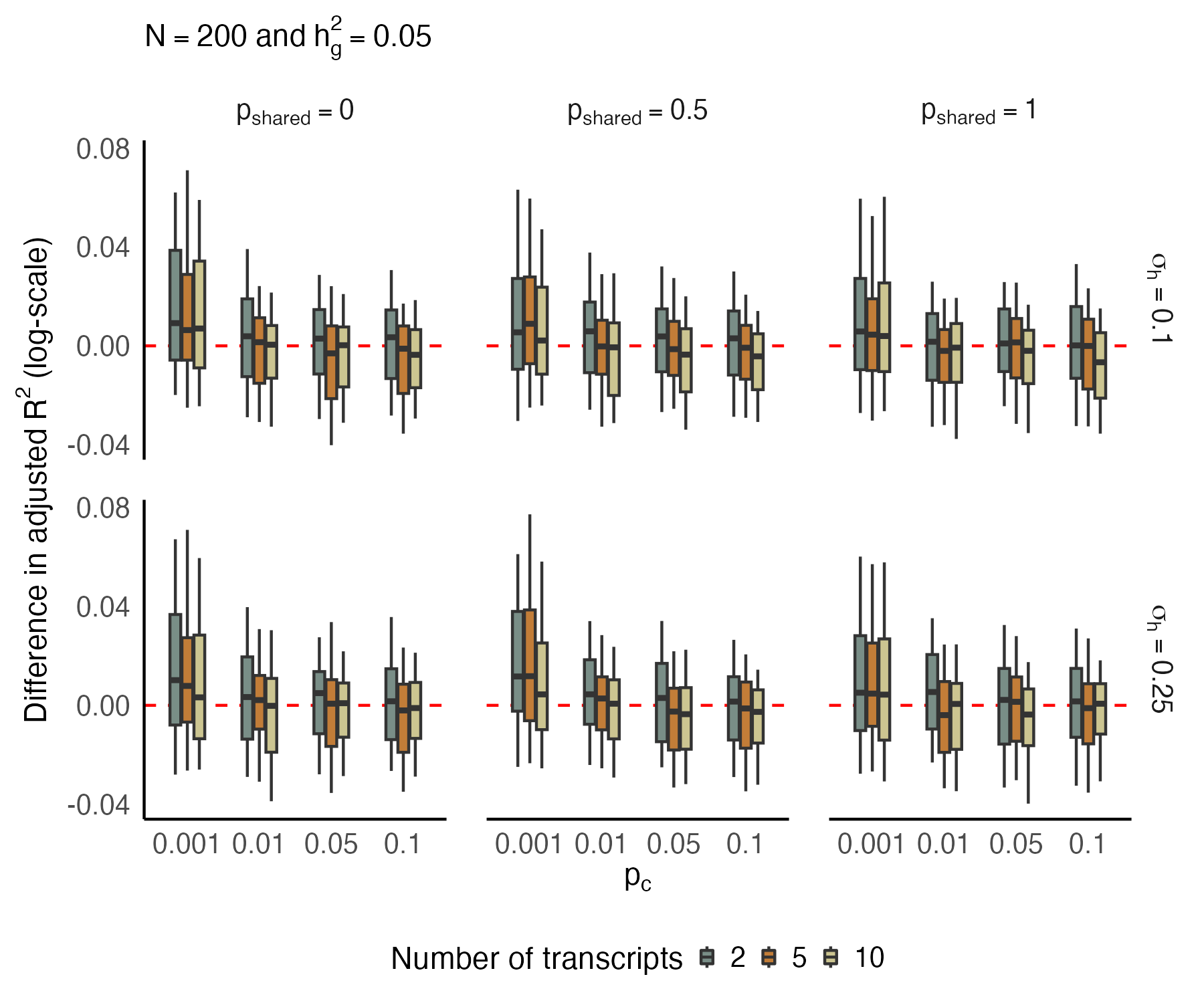

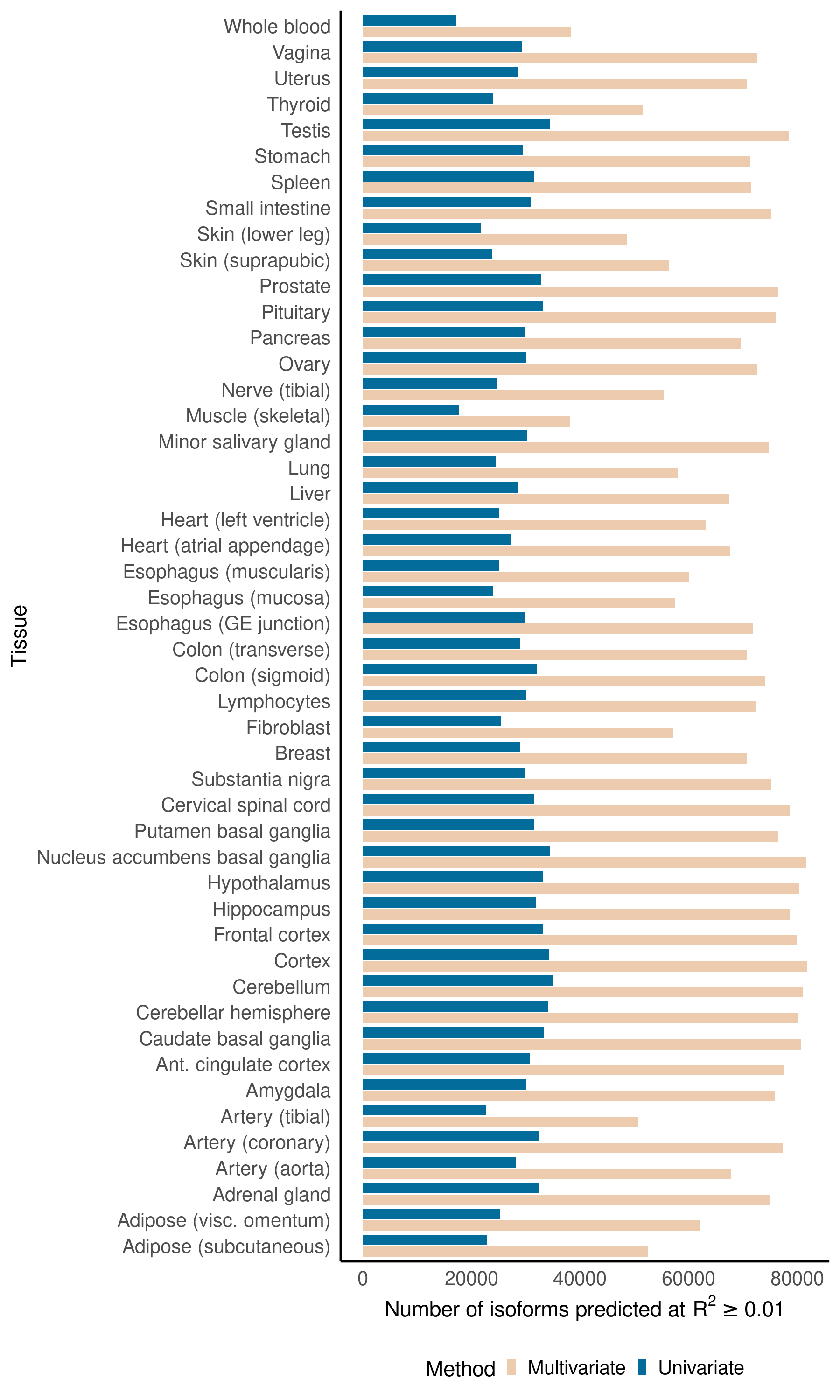

### **Figure S5**: Across 48 GTEx tissues (Y-axis), the number of multivariate (cream) and univariate (blue) models predicting isoform expression at CV R^2^ > 0.01 (X-axis).

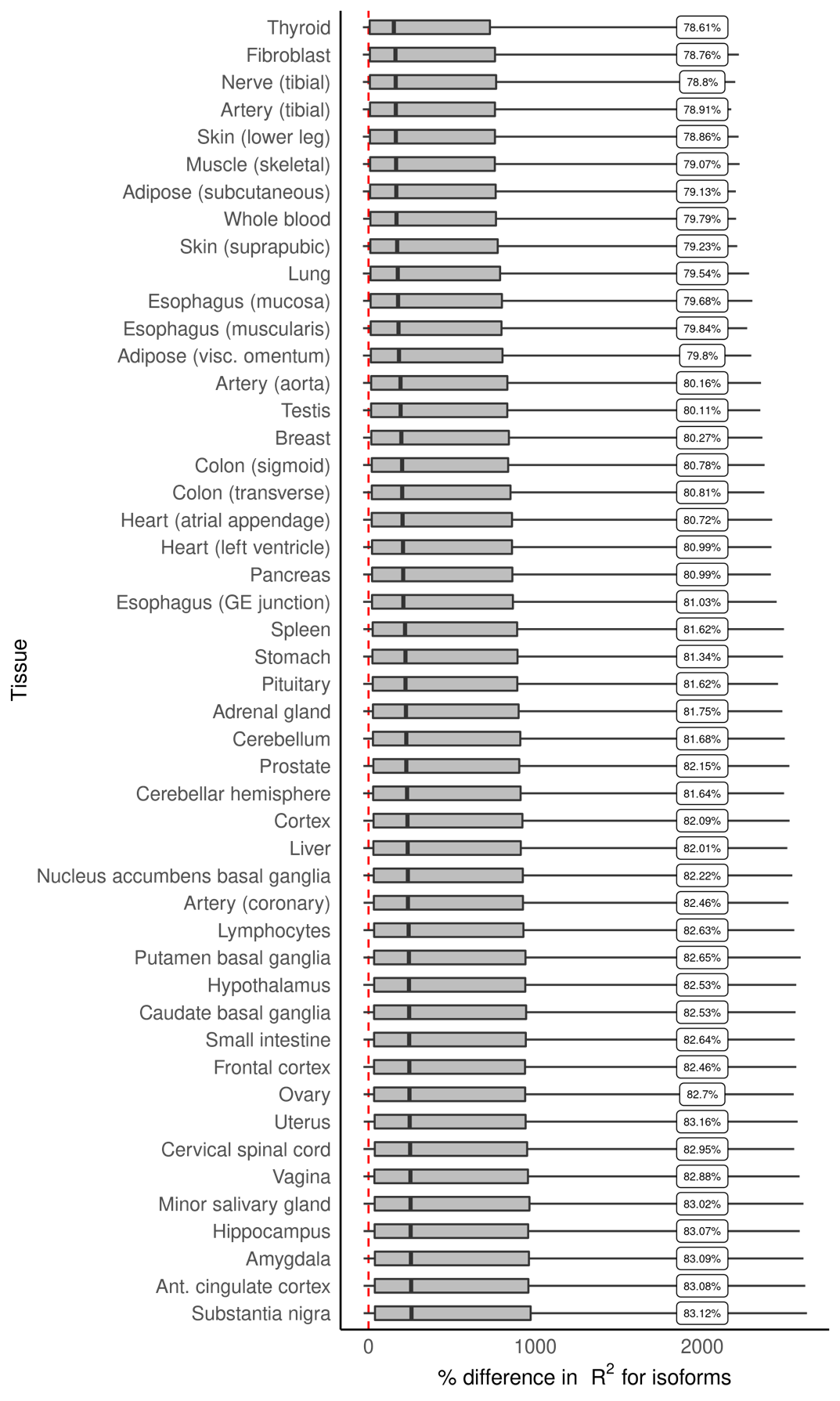

### **Figure S6**: Across 48 GTEx tissues (Y-axis), percent difference in CV R^2^ (X-axis) of prediction of isoform expression using multivariate models versus univariate models. The label shows the proportion of isoforms with improved performance using multivariate models.

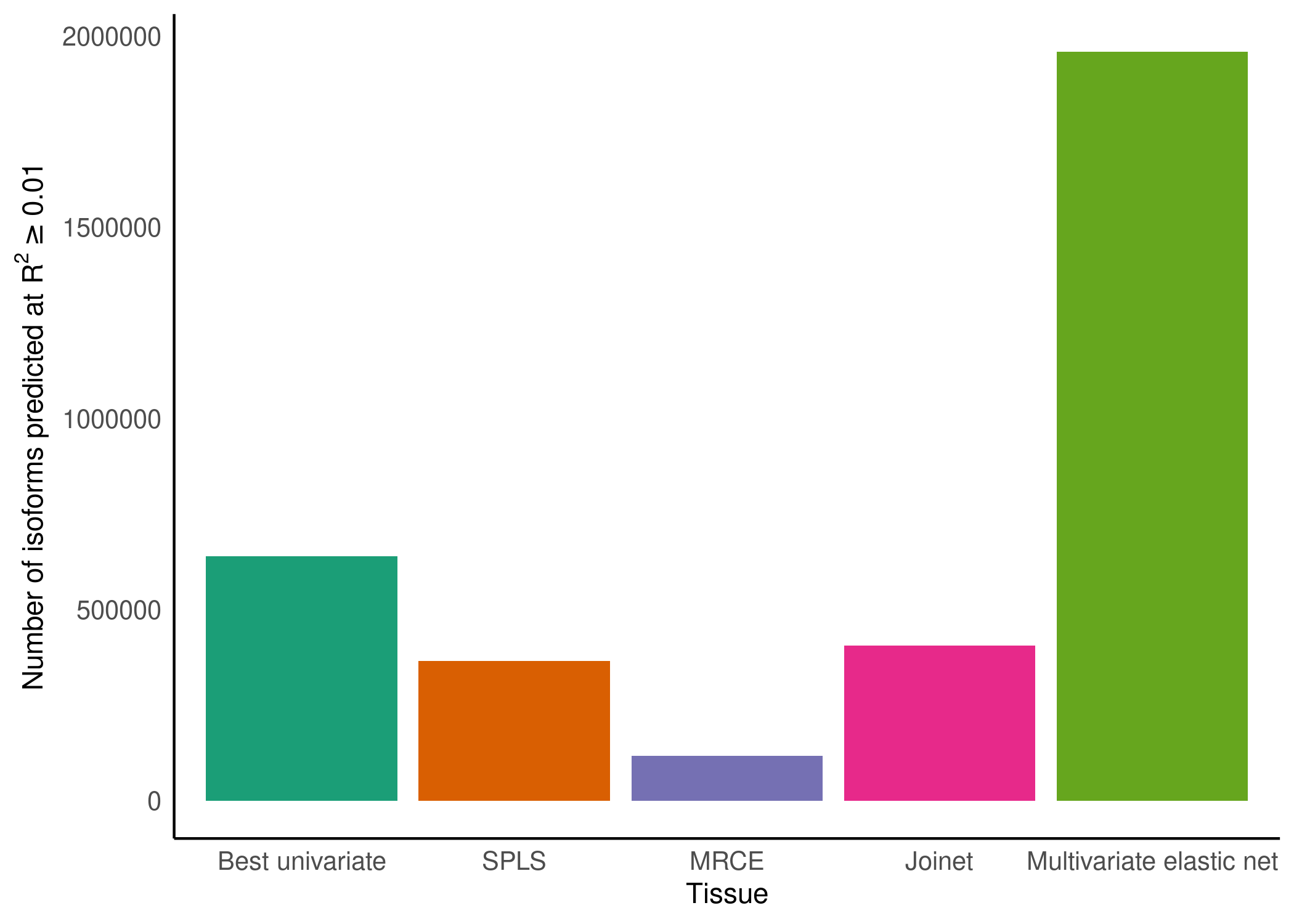

### **Figure S7**: **Best performing predictive model of isoform expression across 48 GTEx tissues**. Y-axis shows the number of isoforms that can be predicted with cross-validated (CV) R^2^ > 0.01. Several different models were compared, including using a univariate framework in which isoforms were modeled independently using the TWAS FUSION framework. Several multivariate models were tested, including sparse partial least squares (SPLS), multivariate LASSO penalized regression with simultaneous covariance estimation (MRCE), elastic net with stacked generalization (Joinet), and multivariate elastic net. As demonstrated, multivariate elastic net substantially outperformed the other multivariate and univariate models.

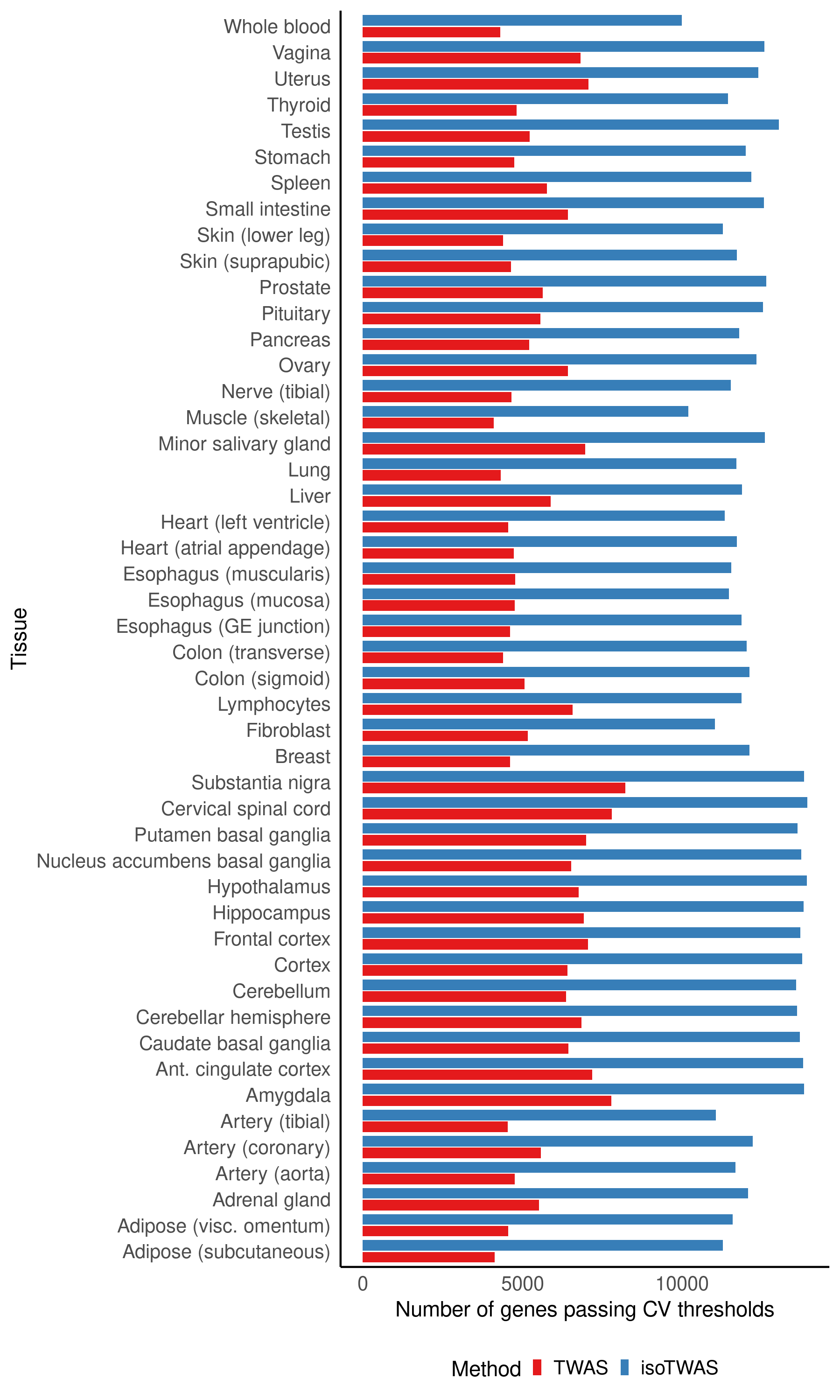

### **Figure S8**: Across 48 GTEx tissues (Y-axis), number of genes that pass TWAS (blue) and isoTWAS (red) CV R^2^ cutoffs to be available for testing in the trait-mapping step (X-axis)

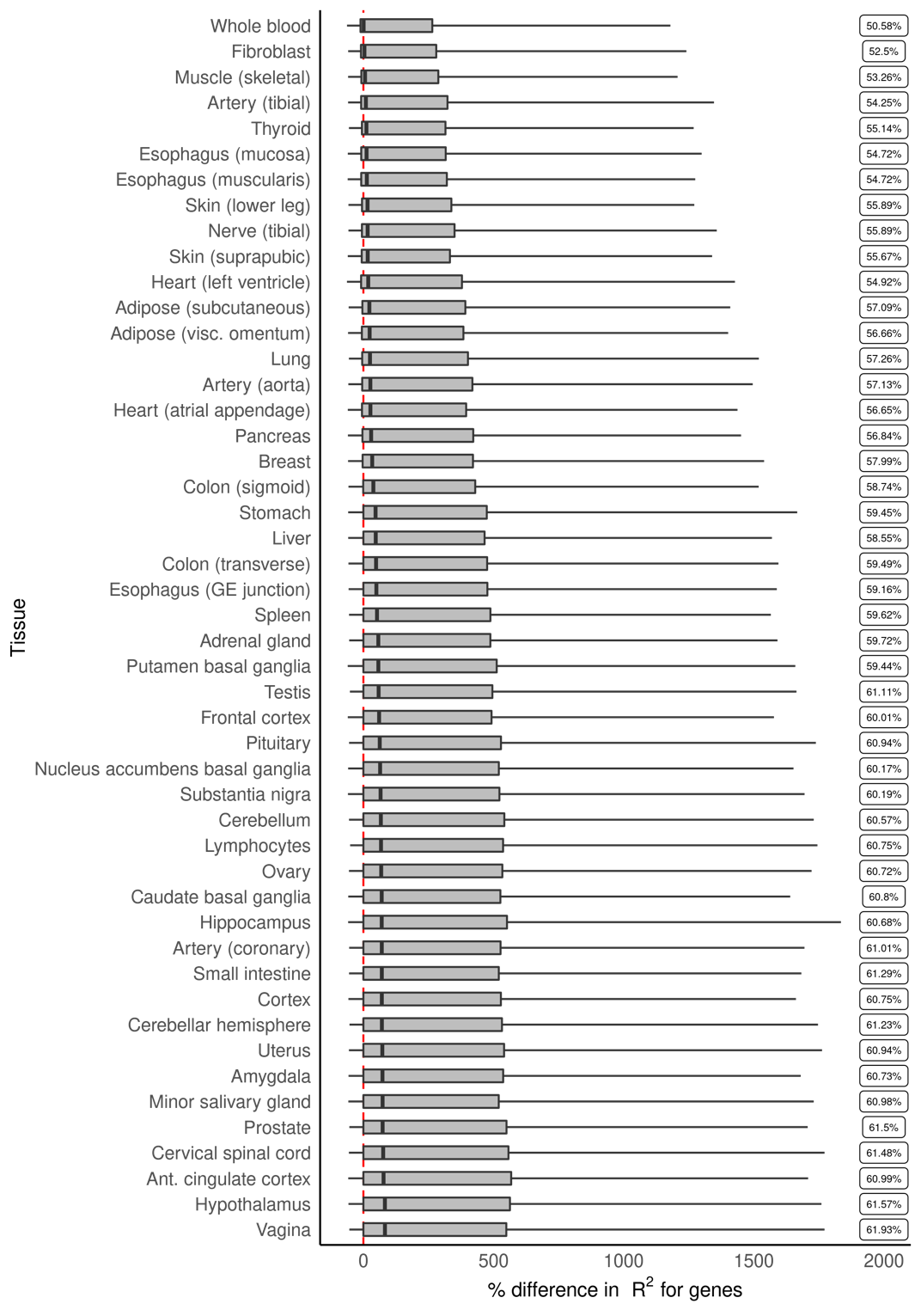

### **Figure S9**: Across 48 GTEx tissues (Y-axis), percent difference in CV R^2^ (X-axis) of prediction of gene expression using isoTWAS compared with TWAS models. The label shows the proportion of isoforms with improved performance using multivariate models.

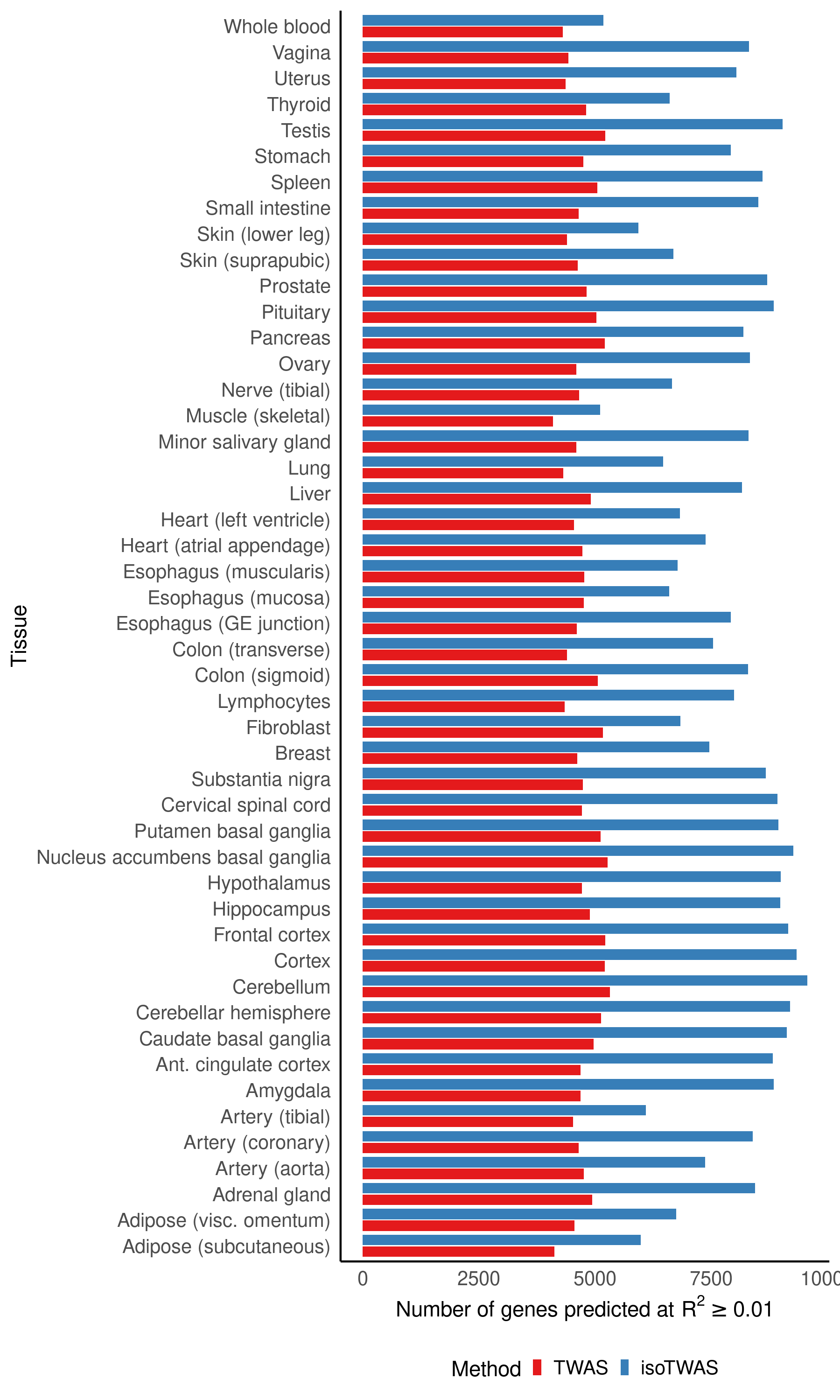

### **Figure S10**: Across 48 GTEx tissues (Y-axis), number of genes predicted at CV R^2^ > 0.01 using TWAS (blue) and isoTWAS (red)

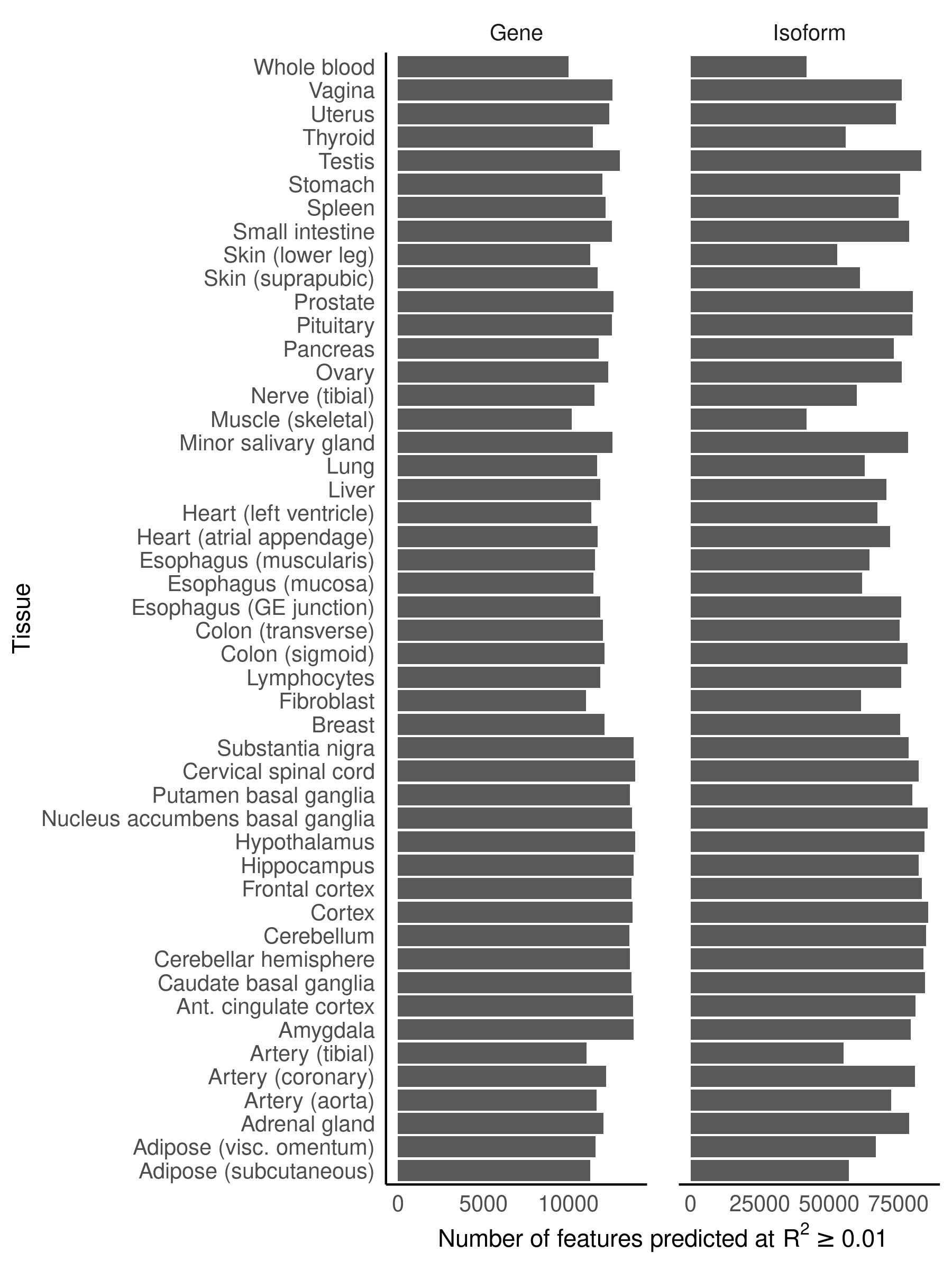

### **Figure S11**: Number of genes (left) and isoforms (right) predicted at CV R^2^ > 0.01 using isoTWAS across 48 GTEx tissues.

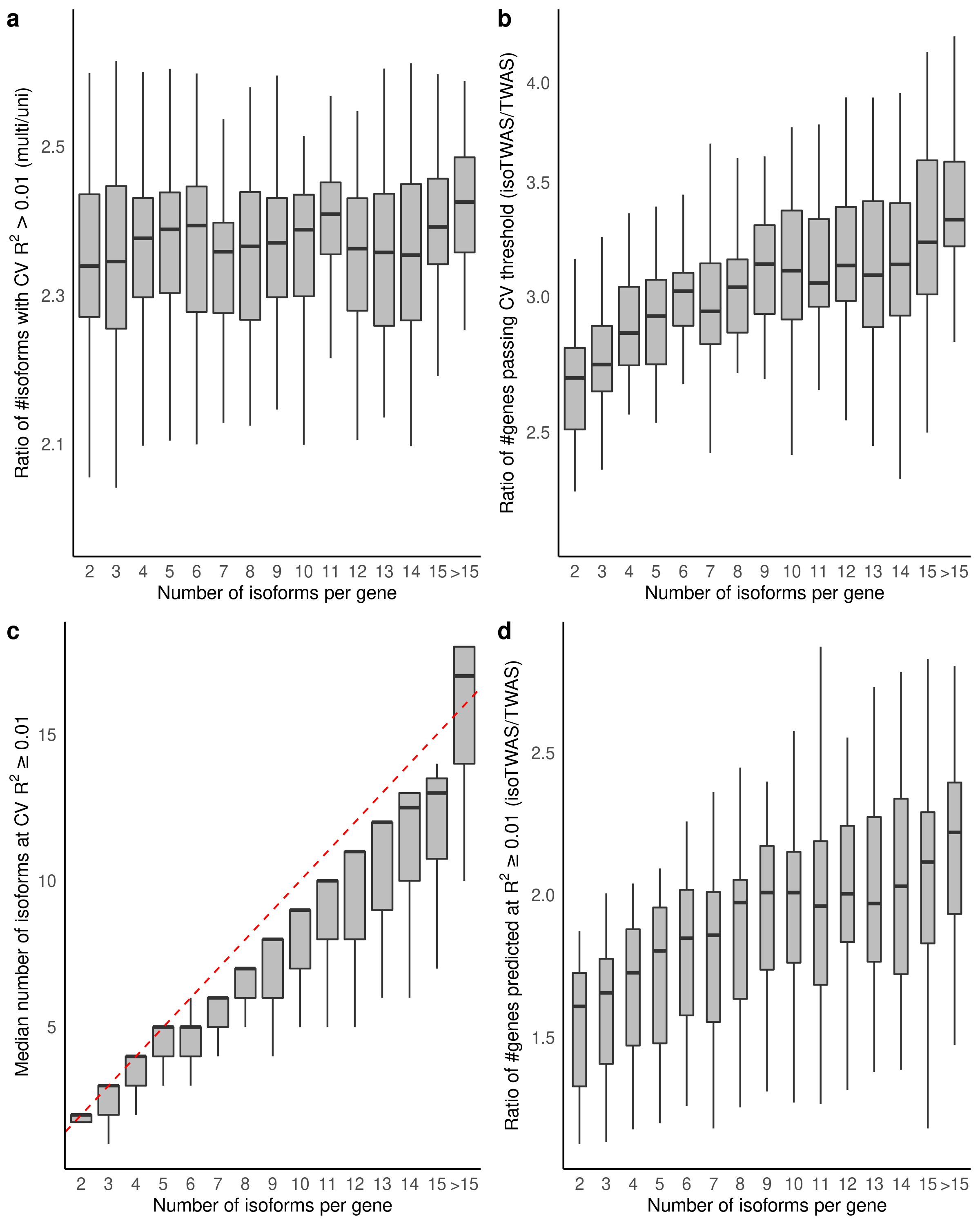

### **Figure S12**: Performance of isoTWAS across number of isoforms per gene across 48 GTEx tissues. **(a)** Ratio of number of isoforms predicted at R^2^ > 0.01 using multivariate versus univariate prediction. **(b)** Ratio of number of genes passing CV threshold using isoTWAS versus TWAS. **(c)** Median number of isoforms predicted at CV R^2^ > 0.01 in isoTWAS models across increasing number of isoforms per gene. The red line shows the line Y = X + 1. **(d)** Ratio of number of genes with CV R^2^ > 0.01 using isoTWAS versus TWAS.

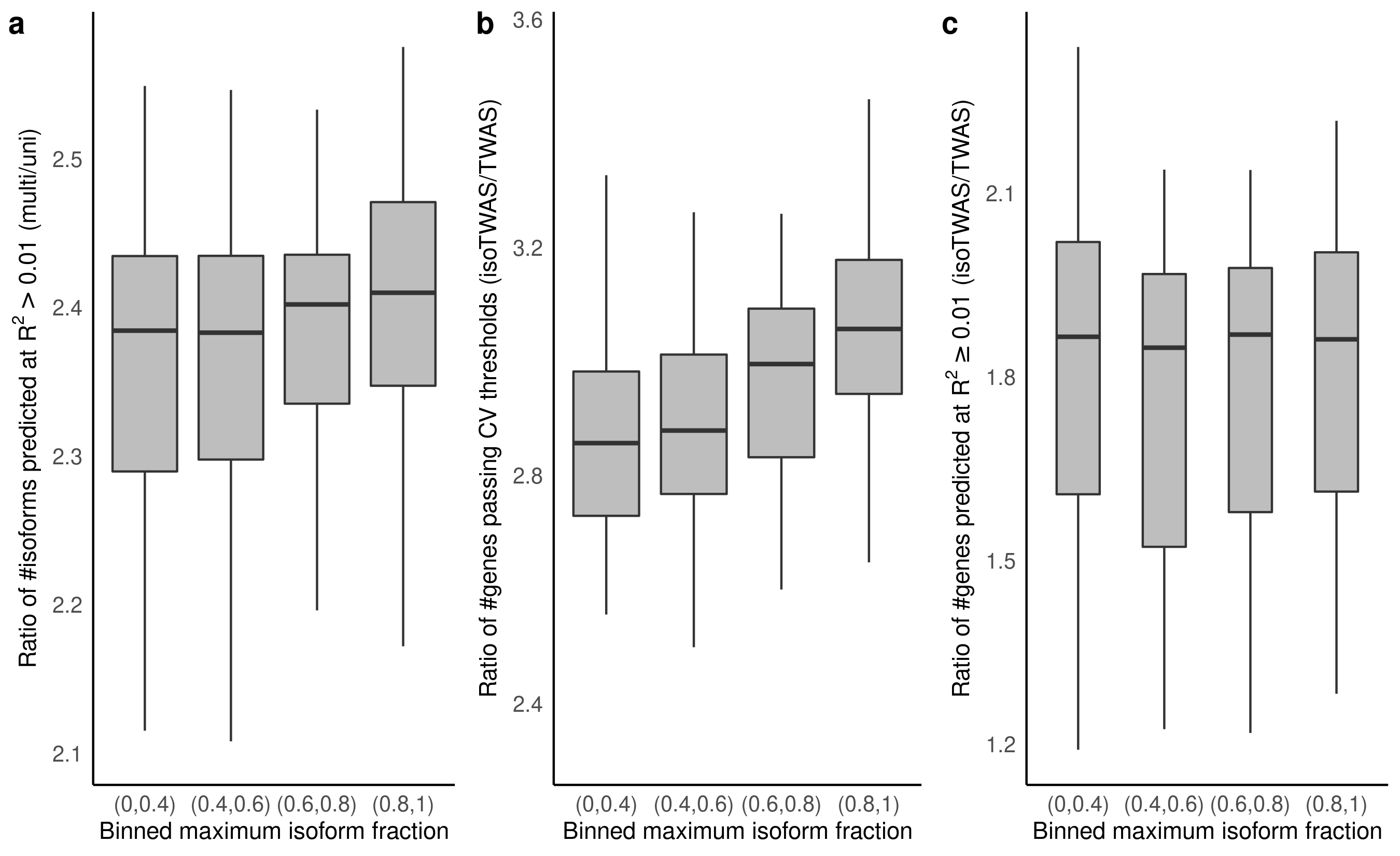

**Figure S13**: Performance of isoTWAS across increasing maximum isoform fraction across 48 GTEx tissues. **(a)** Ratio of number of isoforms predicted at R^2^ > 0.01 using multivariate versus univariate prediction. **(b)** Ratio of number of genes passing CV threshold using isoTWAS versus TWAS. **(c)** Ratio of number of genes with CV R^2^ > 0.01 using isoTWAS versus TWAS.

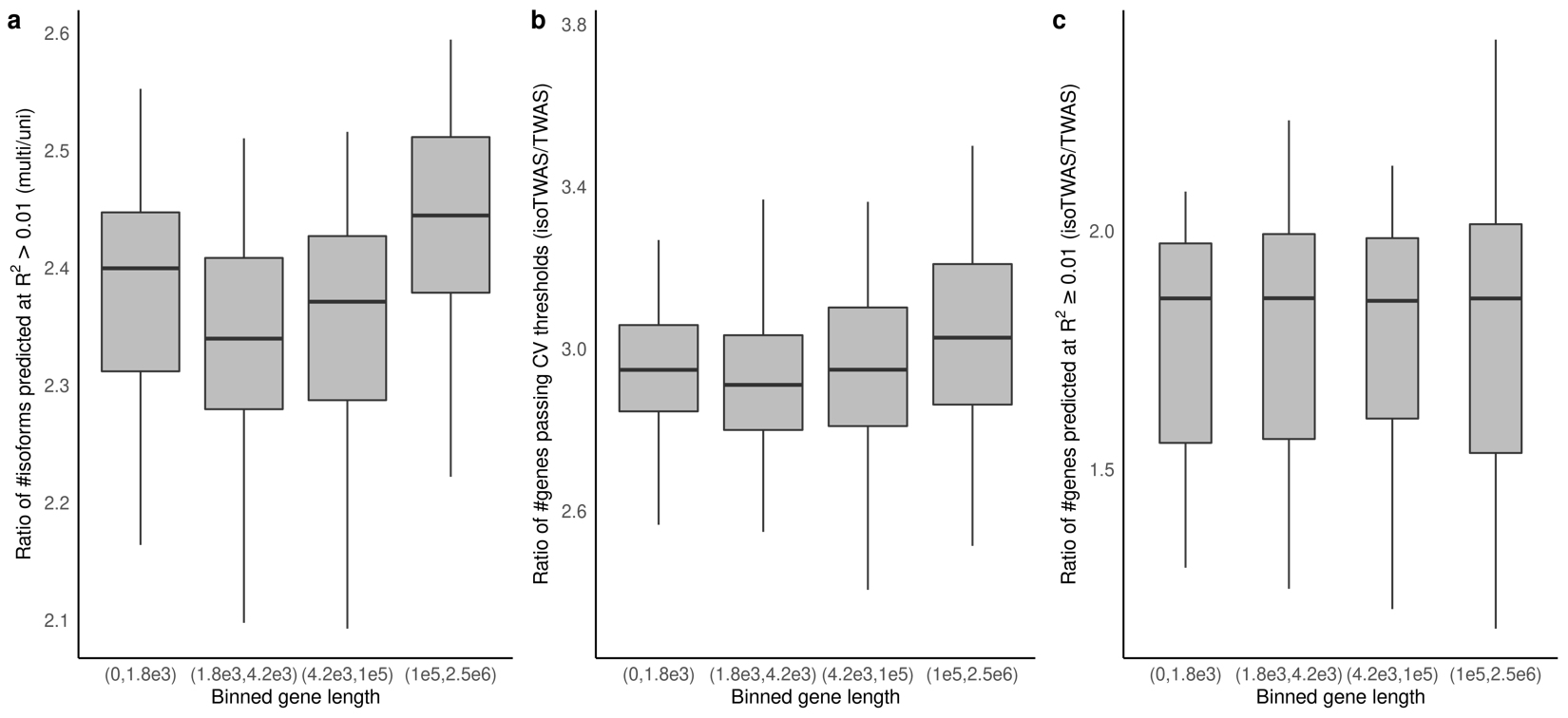

**Figure S14**: Performance of isoTWAS across increasing gene length across 48 GTEx tissues. **(a)** Ratio of number of isoforms predicted at R^2^ > 0.01 using multivariate versus univariate prediction. **(b)** Ratio of number of genes passing CV threshold using isoTWAS versus TWAS. **(c)** Ratio of number of genes with CV R^2^ > 0.01 using isoTWAS versus TWAS. R^2^ > 0.01 using isoTWAS versus TWAS.

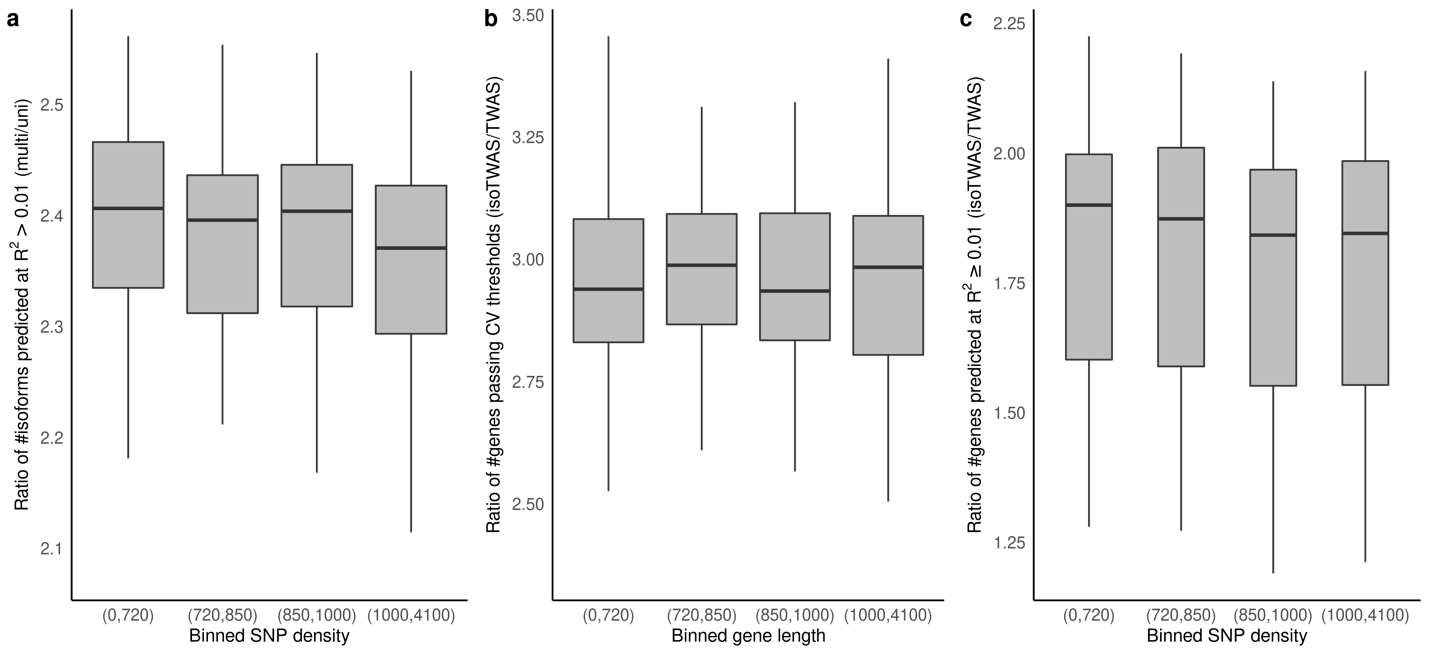

### **Figure S15**: Performance of isoTWAS across increasing SNP density across 48 GTEx tissues. **(a)** Ratio of number of isoforms predicted at R^2^ > 0.01 using multivariate versus univariate prediction. **(b)** Ratio of number of genes passing CV threshold using isoTWAS versus TWAS. **(c)** Ratio of number of genes with CV R^2^ > 0.01 using isoTWAS versus TWAS.

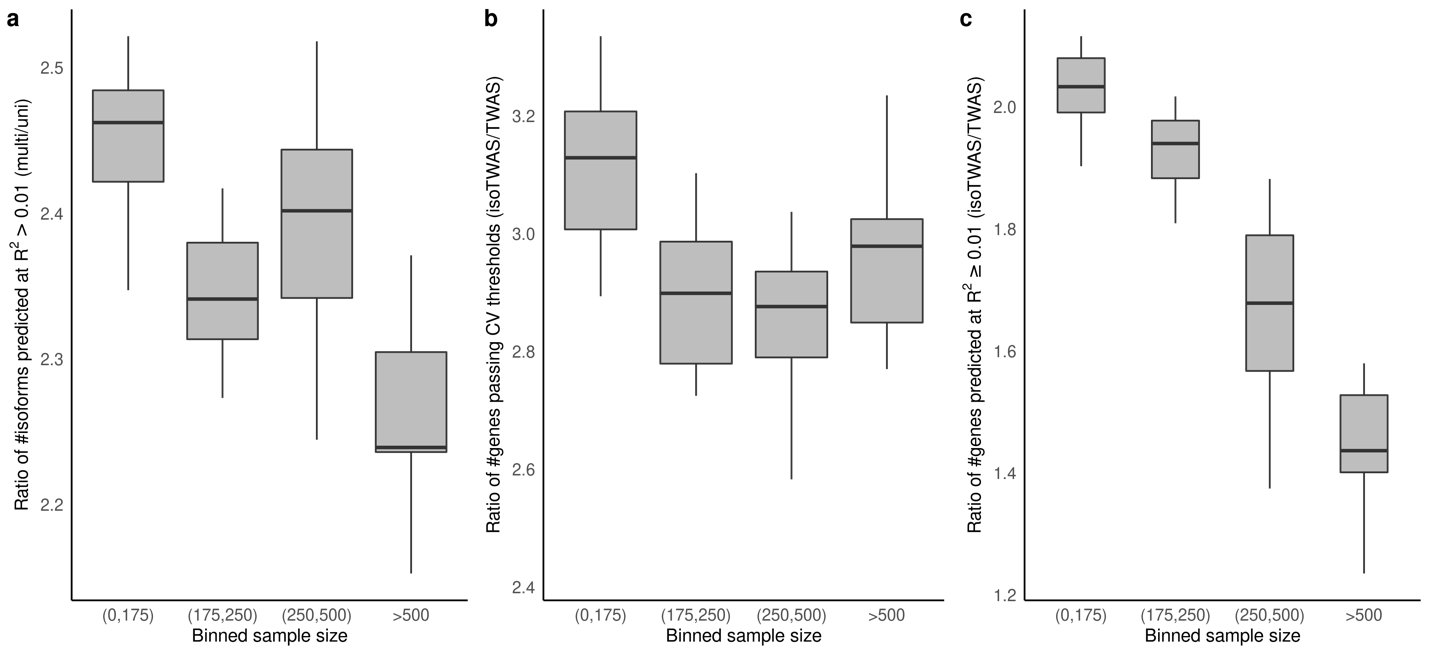
**Figure S16**: Performance of isoTWAS across increasing sample size across 48 GTEx tissues. **(a)** Ratio of number of isoforms predicted at R^2^ > 0.01 using multivariate versus univariate prediction. **(b)** Ratio of number of genes passing CV threshold using isoTWAS versus TWAS. **(c)** Ratio of number of genes with CV R^2^ > 0.01 using isoTWAS versus TWAS.

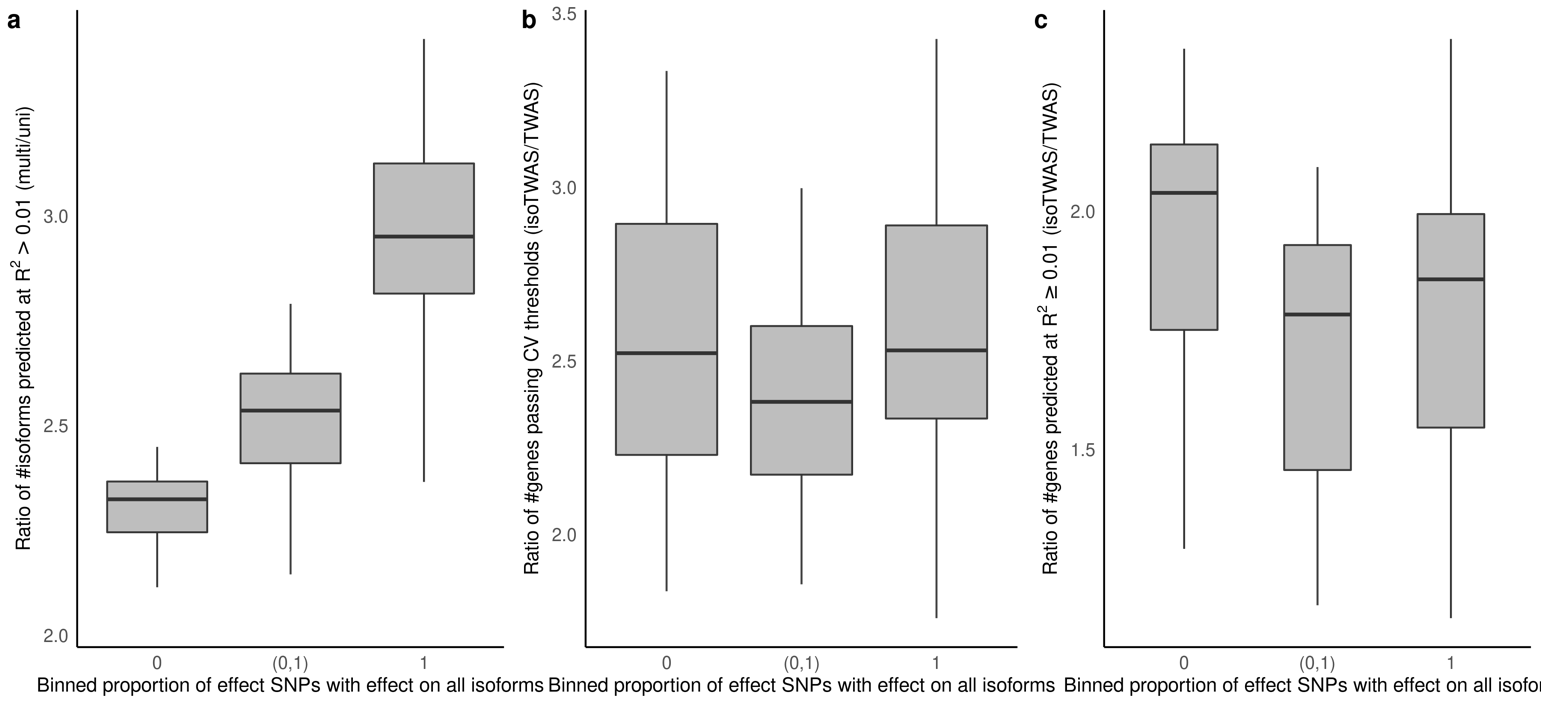

**Figure S17**: Performance of isoTWAS across proportion of SNPs with a non-zero effect on all isoforms of the gene, across 48 GTEx tissues. **(a)** Ratio of number of isoforms predicted at R^2^ > 0.01 using multivariate versus univariate prediction. **(b)** Ratio of number of genes passing CV threshold using isoTWAS versus TWAS. **(c)** Ratio of number of genes with CV R^2^ > 0.01 using isoTWAS versus TWAS.

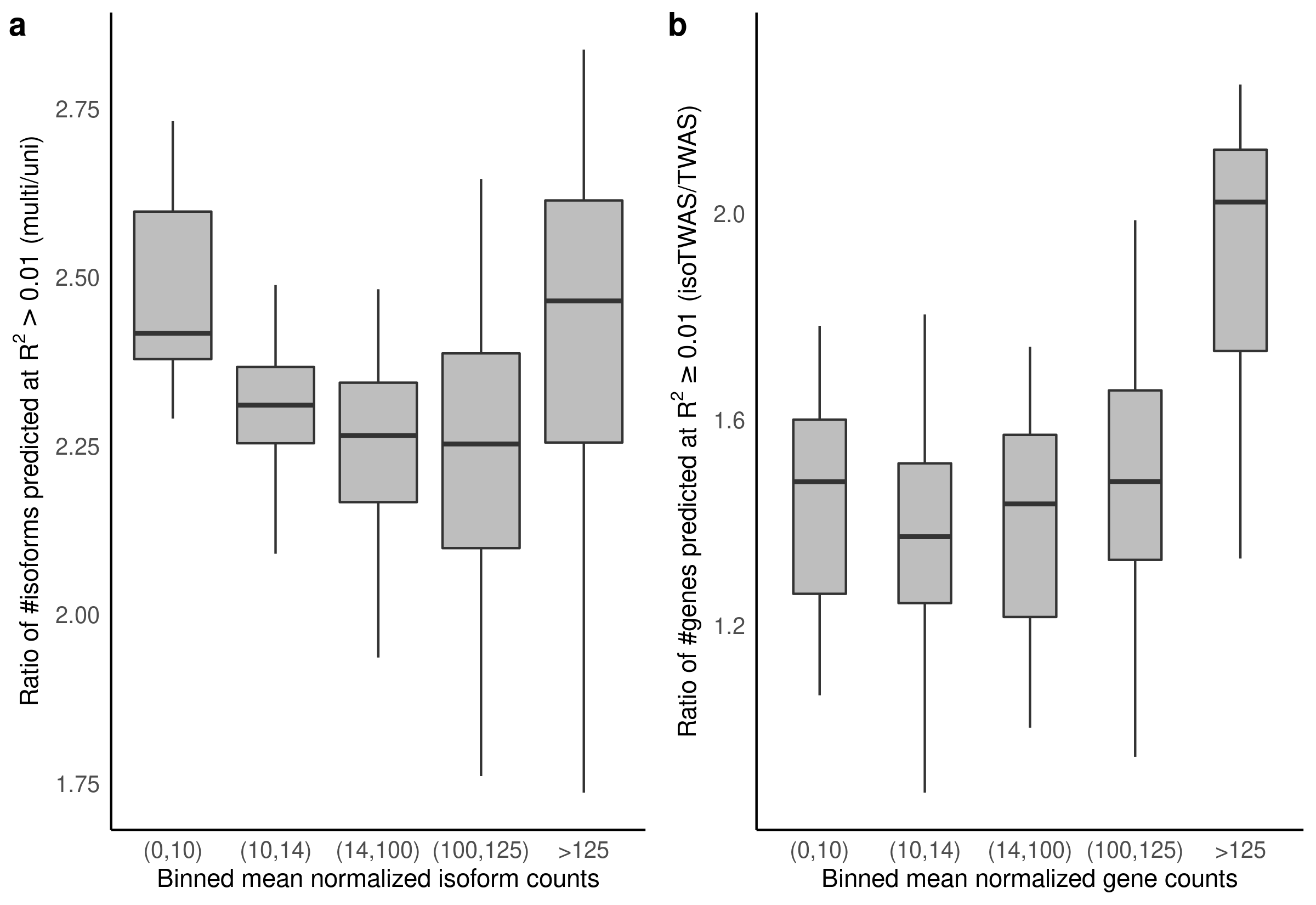

### **Figure S18**: Performance of isoTWAS across increasing mean counts of isoforms and genes across 48 GTEx tissues. **(a)** Ratio of number of isoforms predicted at R^2^ > 0.01 using multivariate versus univariate prediction across increasing mean counts of isoforms. **(b)** Ratio of number of genes with CV R^2^ > 0.01 using isoTWAS versus TWAS across increasing mean counts of genes.

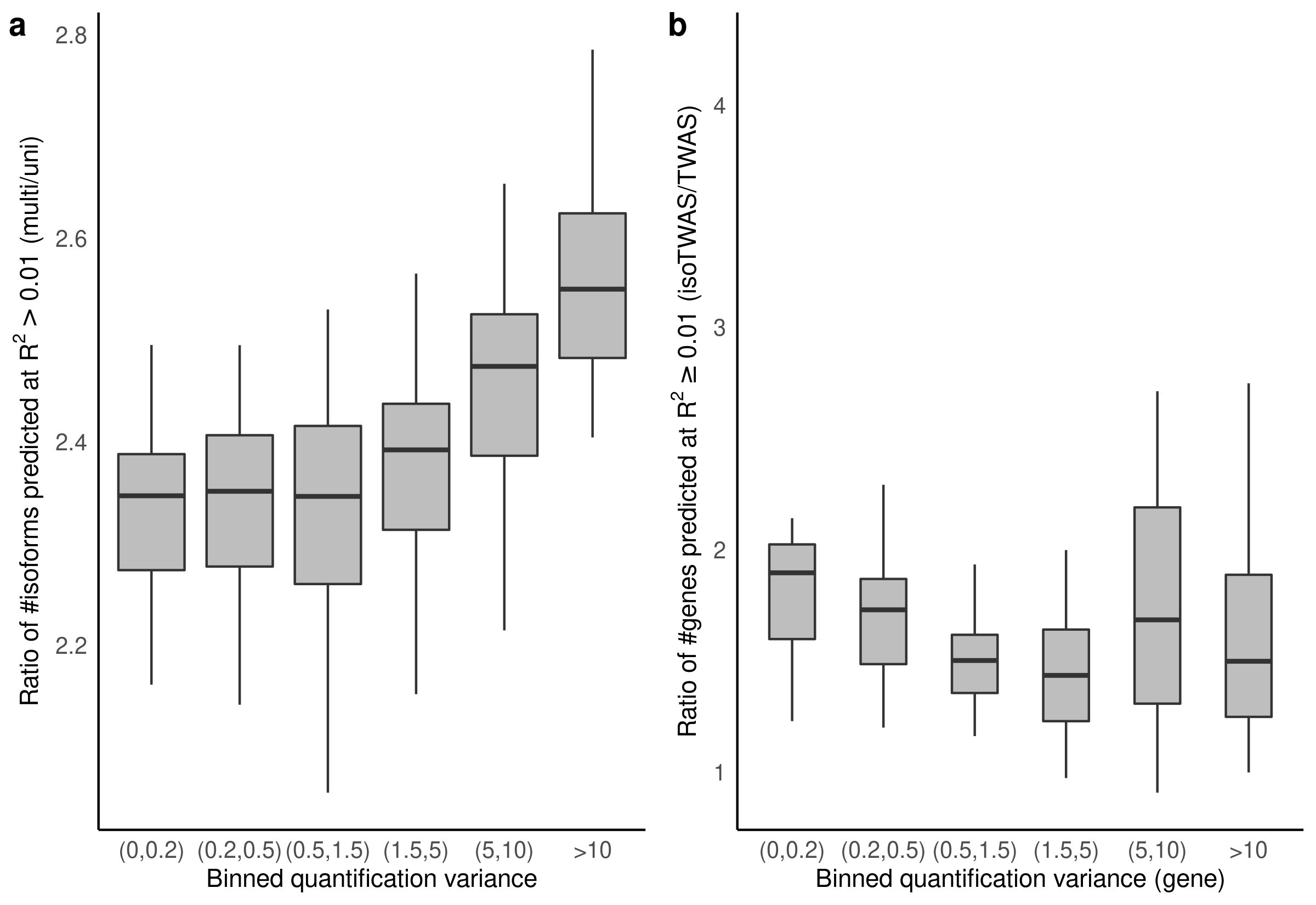

### **Figure S19**: Performance of isoTWAS across increasing quantification variance of isoforms and genes across 48 GTEx tissues. **(a)** Ratio of number of isoforms predicted at R^2^ > 0.01 using multivariate versus univariate prediction across increasing quantification variance of isoforms. **(b)** Ratio of number of genes with CV R^2^ > 0.01 using isoTWAS versus TWAS across increasing quantification variance of genes.

### **Figure S20**: **(a)** Median percent difference in R^2^ of predicting original isoform expression using multivariate versus univariate models across increasing number of isoforms per gene, colored by models trained in the original dataset (pink) and the leave-one-out dataset (teal) **(b)** Median percent difference in R^2^ of predicting original gene expression using isoTWAS versus TWAS models across increasing number of isoforms per gene, colored by models trained in the original dataset (pink) and the leave-one-out dataset (teal)

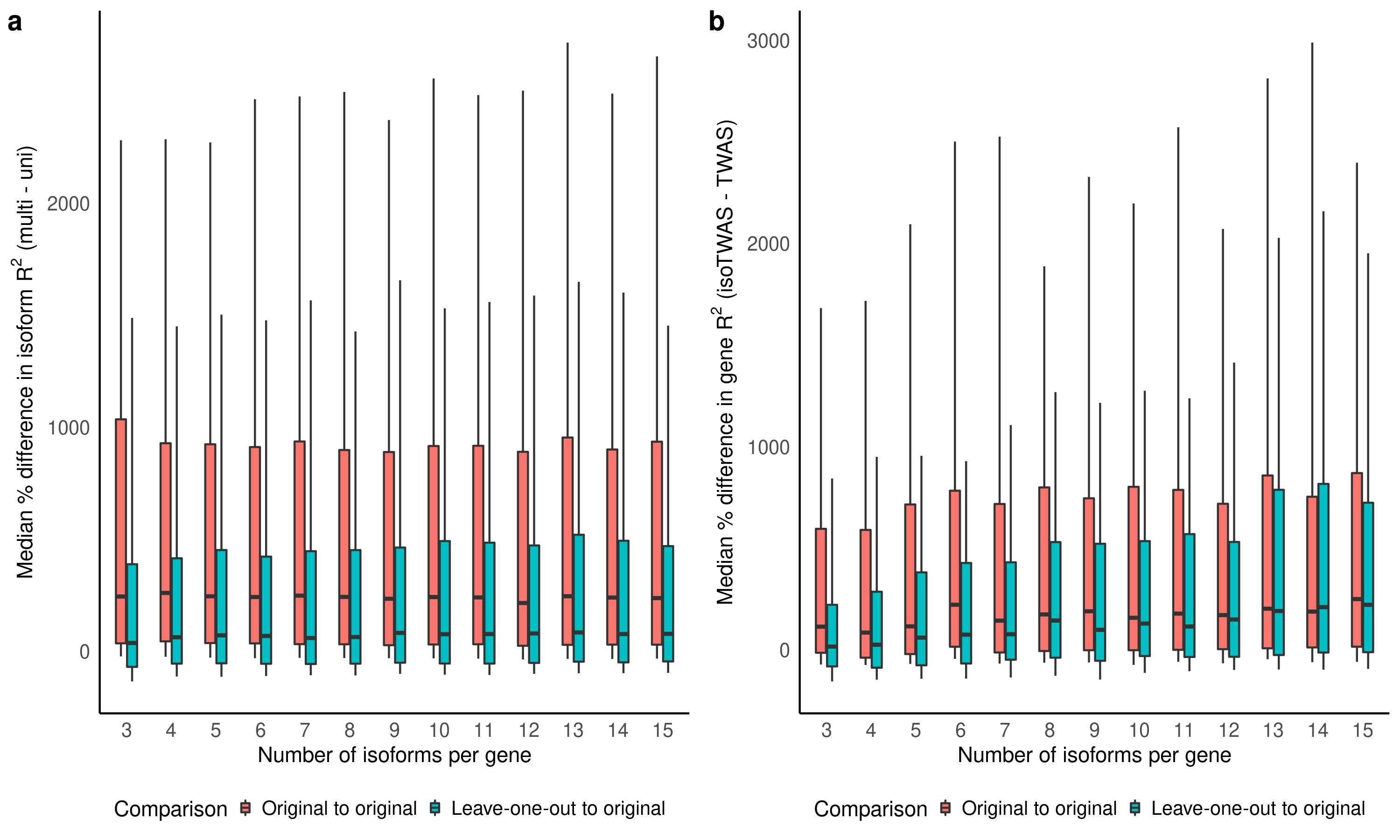

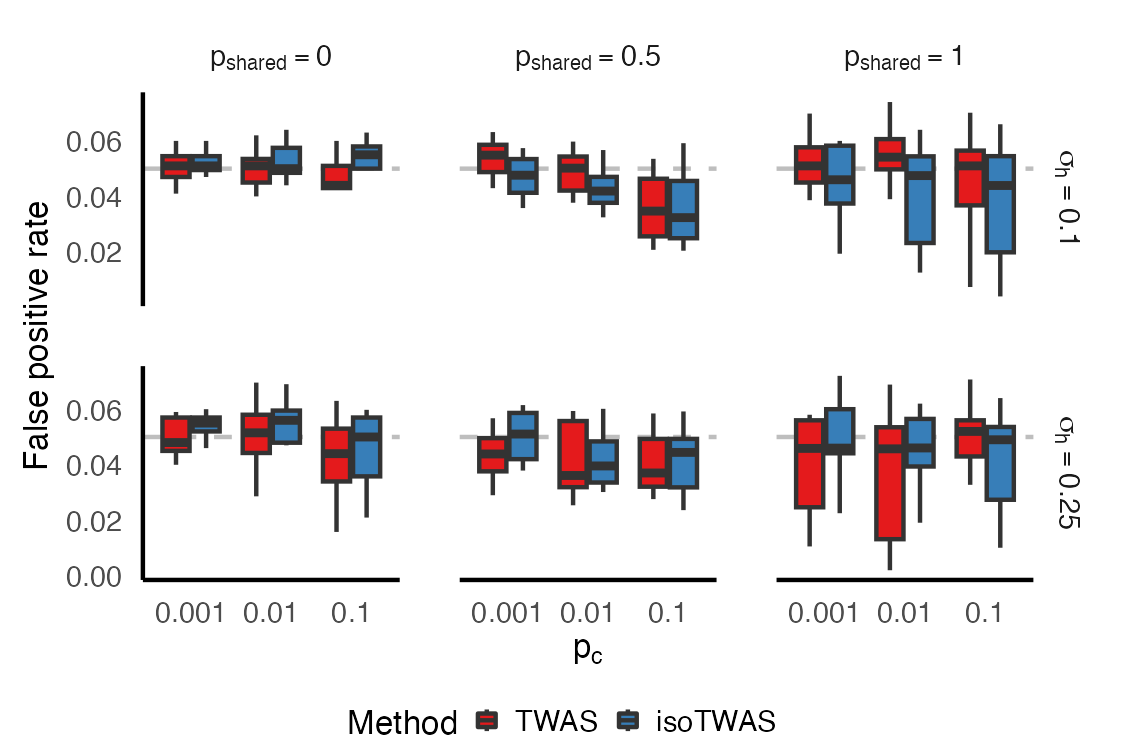

### **Figure S21**: *IsoTWAS and gene-level TWAS show similar false positive rates*. Across 20 iterations of 1,000 simulations, boxplots of false positive rate to detect a gene-trait association using Cauchy-aggregated P-values of isoform-trait associations (red) and gene-level TWAS (blue). In these simulations, we simulate 200 and 5,000 samples in the eQTL and GWAS panels, 5 isoforms, and isoform and gene expression heritability of 0.10. We vary the causal isoQTLs proportion (p_c_, shown on X-axis), the QTL architecture (right margin), and correlation between isoforms (top margin). For each iteration, we simulate one eQTL panel and 1,000 GWAS panels where genetically-regulated gene and isoform expression have no effect on the trait. We calculate the false positive rate as the proportion of the 1,000 tests that give P > 0.05.

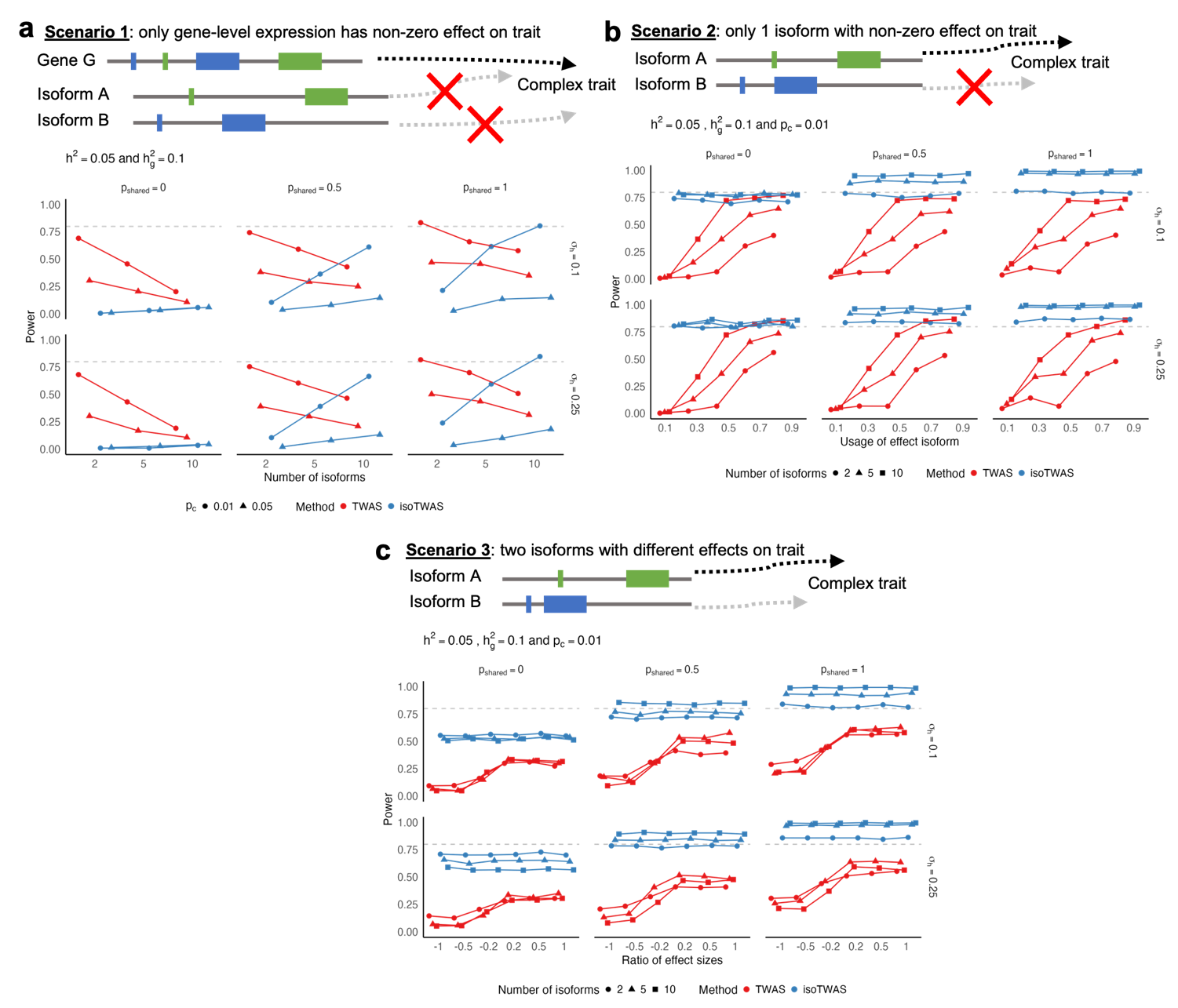
**Figure S22**: Power comparison between TWAS and isoTWAS in detecting gene-trait association across 3 scenarios **(Methods)**. **(a)** Power to detect gene-trait association (proportion of tests with P < 2.5 x 10^-6^, Y-axis) across number of total isoforms per gene (X-axis), facetted by proportion of shared isoQTLs (top margin) and proportion of expression heritability attributed to shared non-genetic effects across isoforms (right margin). Points are shaped by causal isoQTL proportion and colored by method. **(b)** Power to detect gene-trait association (proportion of tests with P < 2.5 x 10^-6^, Y-axis) across proportion of gene expression explained by effect isoform (X-axis), facetted by proportion of shared isoQTLs (top margin) and proportion of expression heritability attributed to shared non-genetic effects across isoforms (right margin). Points are shaped by number of isoforms per gene and colored by method. **(c)** Power to detect gene-trait association (proportion of tests with P < 2.5 x 10^-6^, Y-axis) across ratio of effect sizes of 2 effect isoforms (X-axis), facetted by proportion of shared isoQTLs (top margin) and proportion of expression heritability attributed to shared non-genetic effects across isoforms (right margin). Points are shaped by number of isoforms per gene and colored by method. Here, expression heritability is set of 0.05, trait heritability is set to 0.1, and causal proportion of **(b-c)** is set of 0.01.

**Figure S23**:
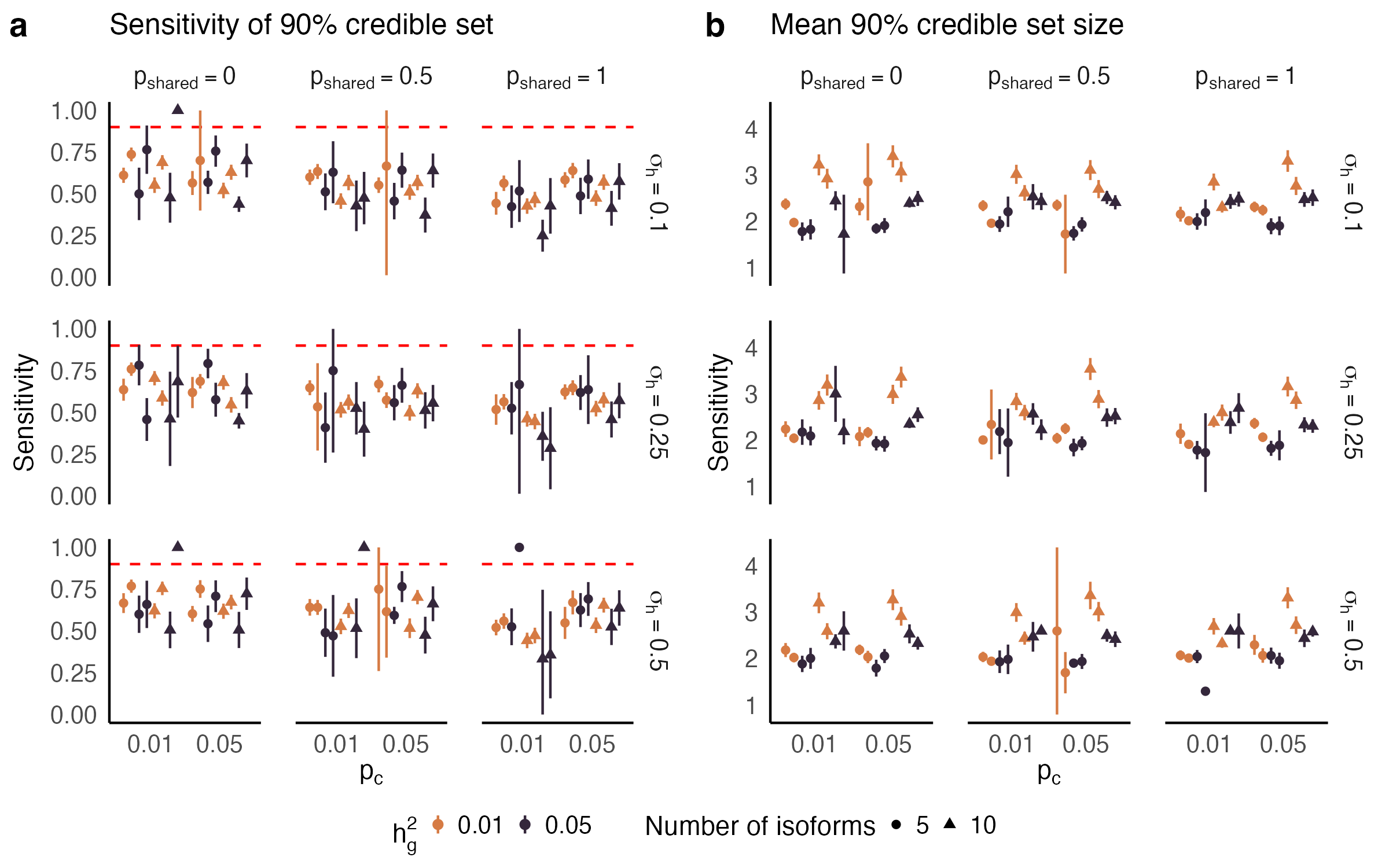
Sensitivity **(a)** and mean set size **(b)** of 90% credible set using FOCUS to finemap isoform-trait associations for a single gene, across causal isoQTL proportion (X-axis). Points are colored by trait heritability and shaped by the number of isoforms per gene. Plots are facetted by proportion of shared isoQTLs (top margin) and proportion of expression heritability attributed to shared non-genetic effects across isoforms (right margin). Line-range shows a 95% jackknife confidence interval.

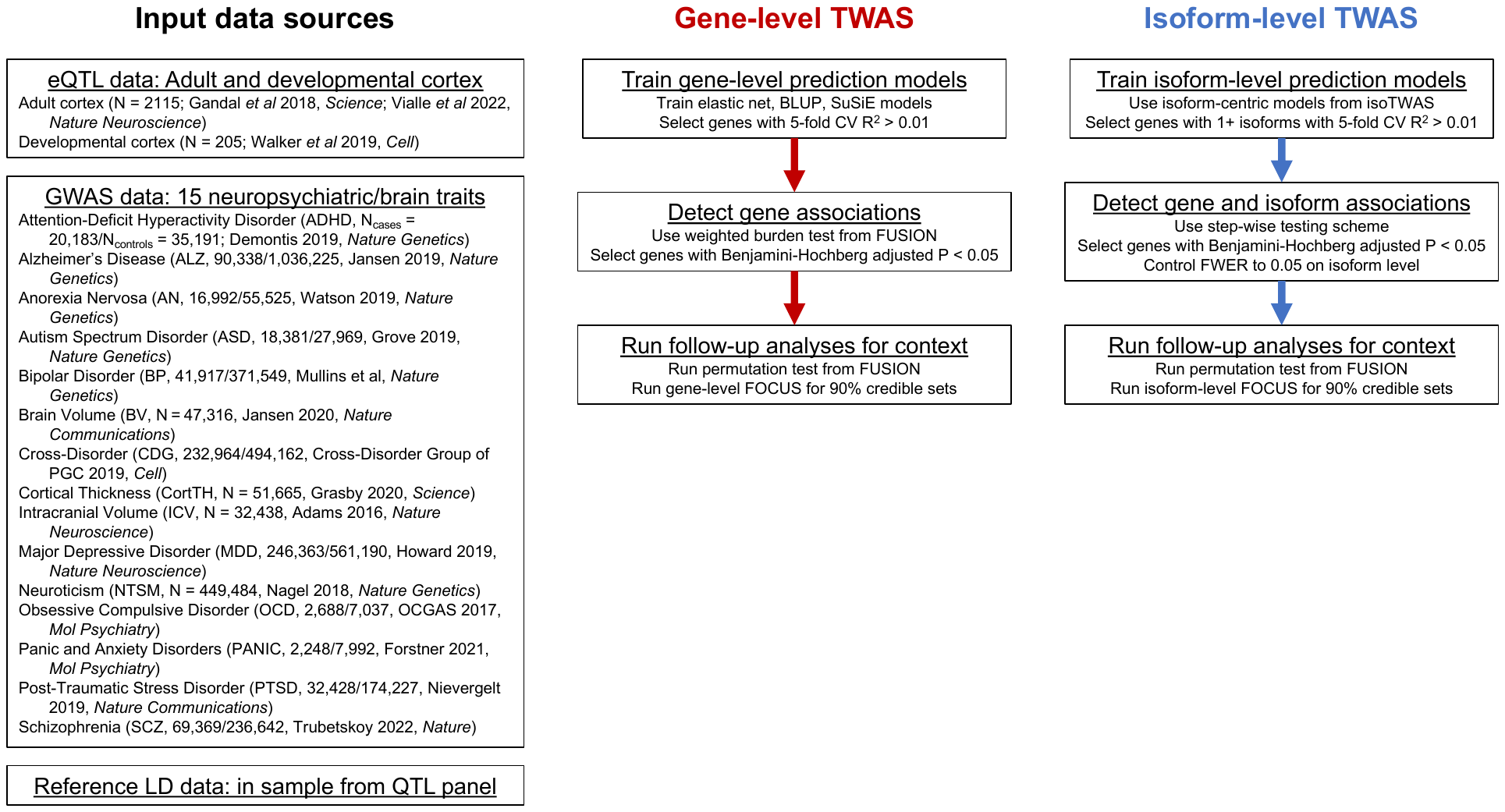

### **Figure S24**: *Schematic diagram for analysis using adult and developmental frontal cortex data from PsychENCODE and AMP-AD*. Data sources for eQTL reference data, GWAS cohorts, and reference LD data are provided on the left (black). The full gene-level TWAS (red) and isoTWAS (blue) are summarized on the right.

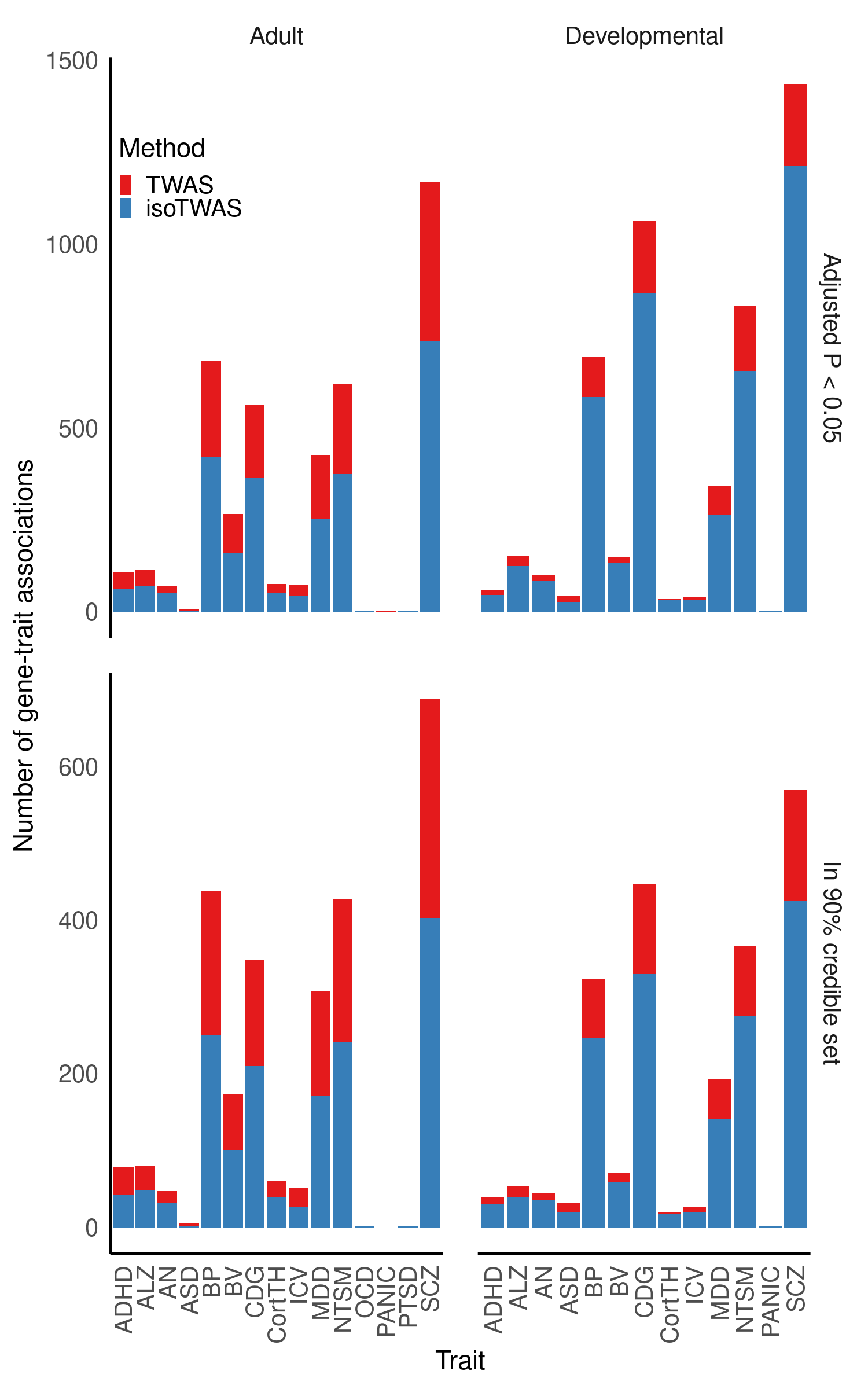
**Figure** **S25**: Number of gene-trait associations (Y-axis) using TWAS (red) and isoTWAS (blue) across trait (X-axis), faceting by tissue (top margin) and threshold (right margin: adjusted P < 0.05 and permutation P < 0.05, top; in 90% credible set using FOCUS fine-mapping, bottom).

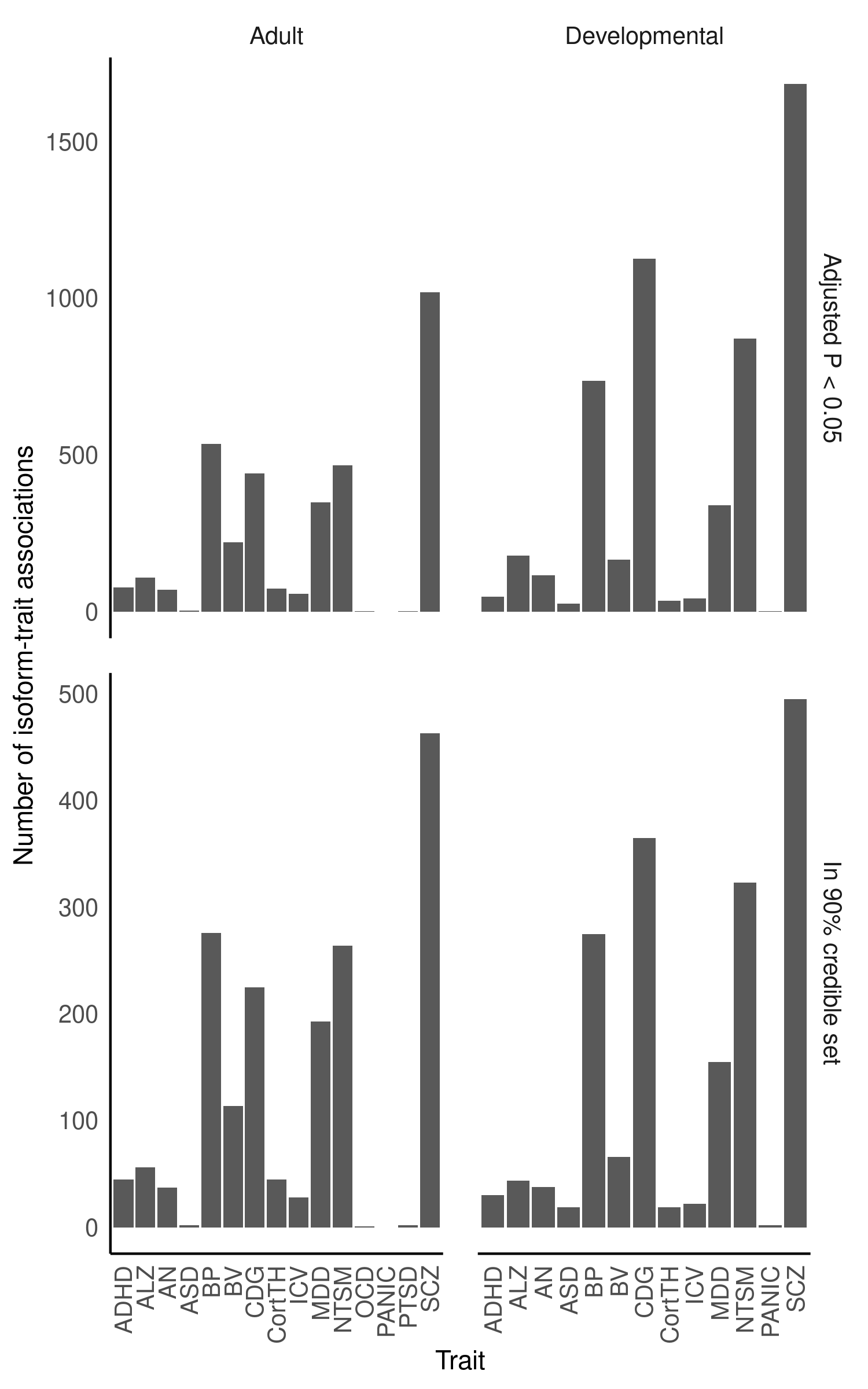
**Figure** **S26**: Number of isoform-trait associations (Y-axis) using isoTWAS across trait (X-axis), faceting by tissue (top margin) and threshold (right margin: adjusted P < 0.05 and permutation P < 0.05, top; in 90% credible set using FOCUS fine-mapping, bottom).

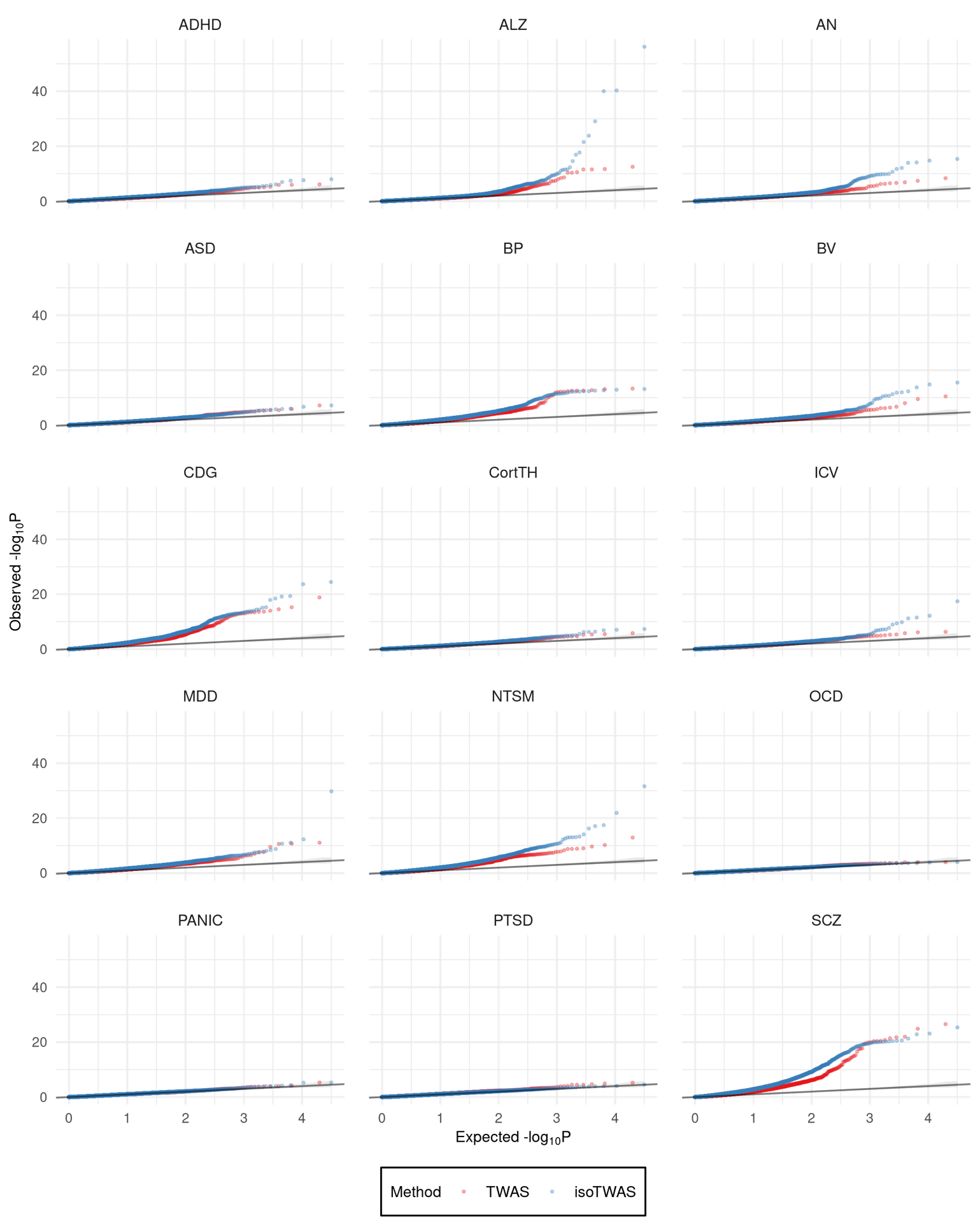
**Figure** **S27**: QQ-plots of gene-level P-values using TWAS (red) and isoTWAS (blue) across 15 traits.

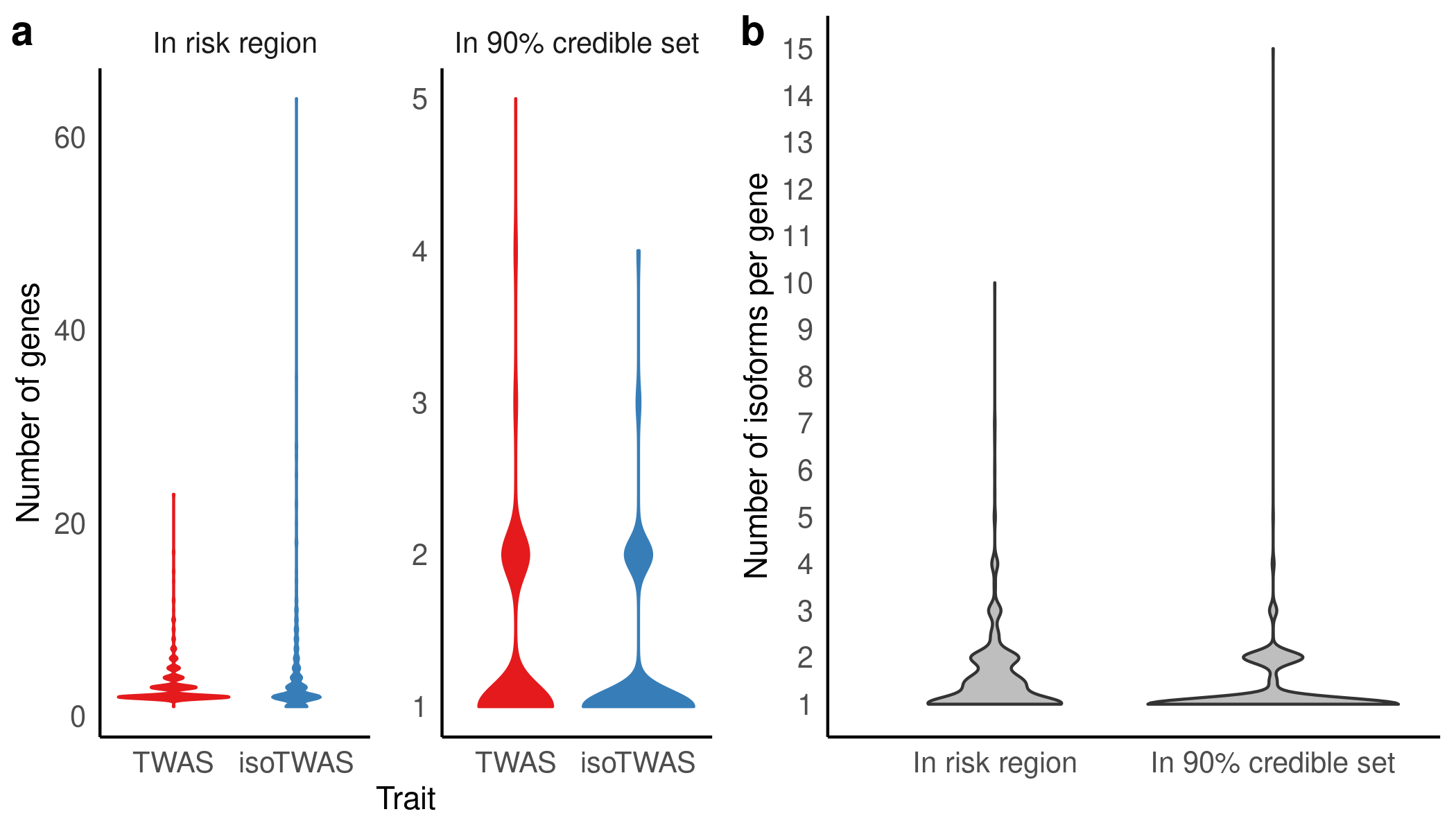

**Figure** **S28**: **(a)** Distribution of number of genes in risk region (left) and in 90% credible set (right) using TWAS and isoTWAS. **(b)** Distribution of number of isoforms per gene in risk region (left) and in 90% credible set (right) using isoTWAS.

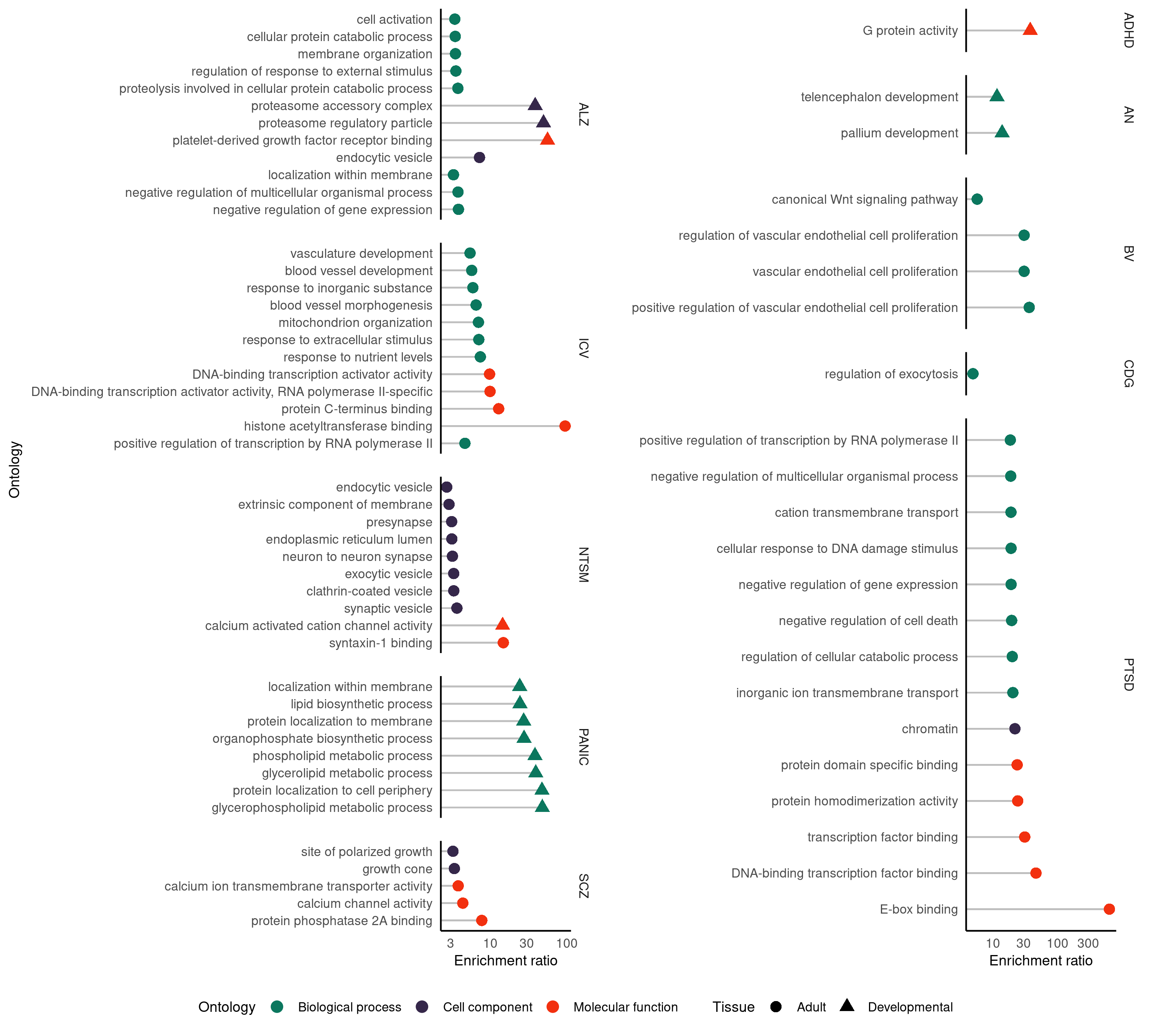

**Figure** **S29**: Lollipop plot of enrichment ratio (X-axis) of ontologies (Y-axis) for isoTWAS-prioritized genes associated at adjusted P < 0.05 and permutation P < 0.05. Points are shaped by tissue type (adult or developmental) and colored by ontology type (biological process, cell component, molecular function).

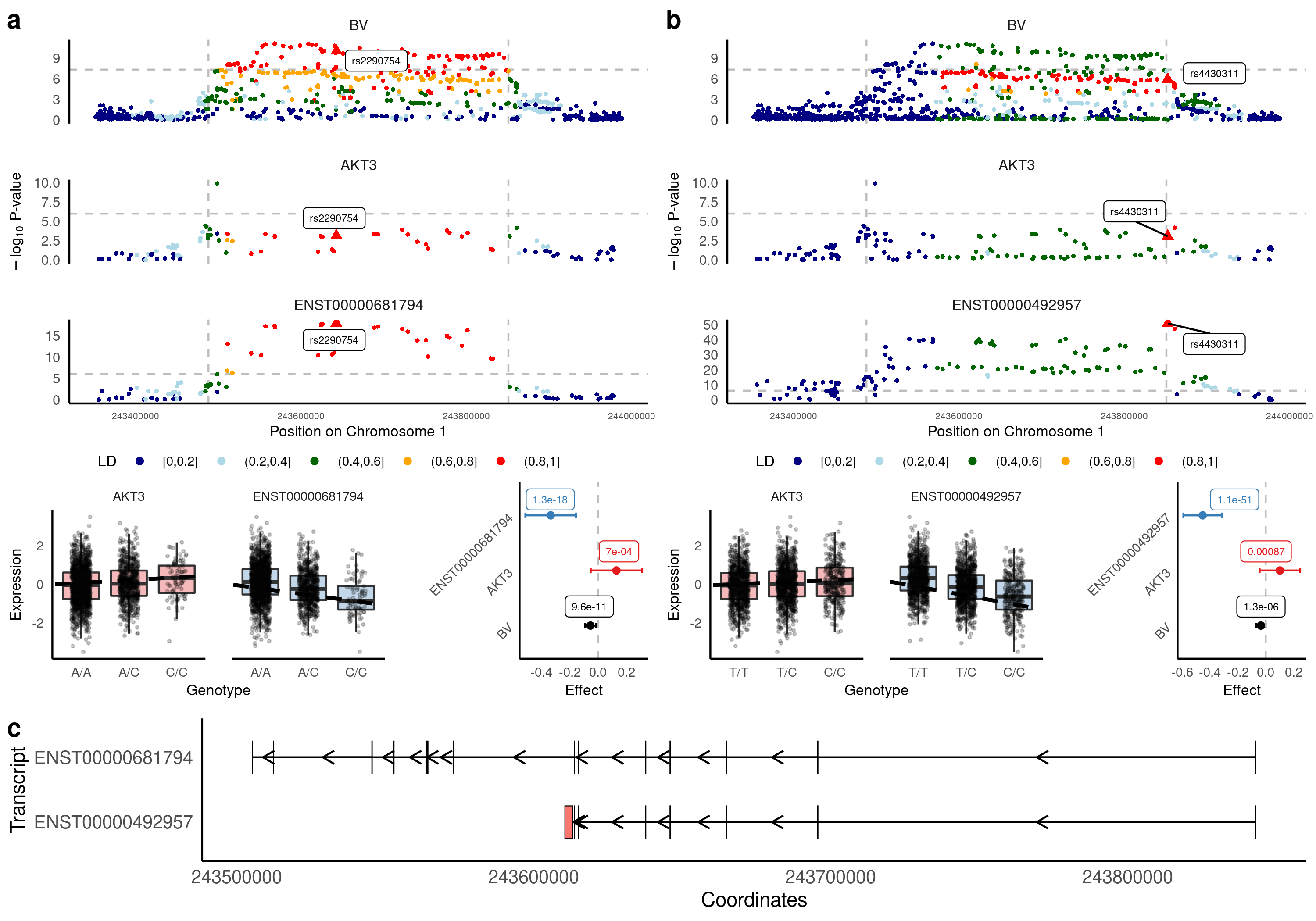

**Figure** **S30**: For ENST00000681794 association with BV **(a)** and ENST00000492957 with BV **(b)**, Manhattan plots of GWAS, eQTL, and isoQTL signal colored by LD (top), boxplots of gene (red) and isoform (blue) expression (Y-axis) by genotype (X-axis) (bottom left), and forest plot of lead isoQTL association with isoform (blue), gene (red), and trait (black) (bottom right). **(c)** Comparison of exon and intron structure of ENST00000681794 and ENST00000492957, based on Gencode v38 reference.
